# Mixed methods systematic review and metasummary about barriers and facilitators for the implementation of cotrimoxazole and isoniazid – preventive therapies for people living with HIV

**DOI:** 10.1101/2021.04.30.21256370

**Authors:** Pia Müller, Luís Velez Lapão

## Abstract

**Background:** Cotrimoxazole and isoniazid preventive therapy (CPT, IPT) have been shown to be efficacious therapies for the prevention of opportunistic infections and tuberculosis (TB) among people living with human immunodeficiency virus (HIV). Despite governments’ efforts to translate World Health Organization recommendations into practice, implementation remains challenging. This review aimed to explore and compare CPT and IPT with respect to similarities and differences of barriers identified across high TB/HIV burden countries. A secondary objective was to identify facilitators for implementing both preventive therapies.

**Methods:** We searched MEDLINE, Web of Science and SCOPUS databases for peer-reviewed literature published before September 2020. We extracted and synthesized our findings using Maxqda software. We applied framework synthesis in conjunction with metasummary to compare both therapies with respect to similarities and differences of barriers identified across seven health system components (in line with the modified WHO’s Framework for action). Protocol registration: PROSPERO (CRD42019137778).

**Findings:** We identified four hundred and eighty-two papers, of which we included forty for review. Although most barrier themes were identical for both preventive therapies, we identified seven intervention-specific themes. Like for CPT, barriers identified for IPT were most frequently classified as ‘service delivery-related barriers’ and ‘patient & community-related barriers’. ‘Health provider-related barriers’ played an additional important role for implementing of IPT. Most facilitators identified referred to health system strengthening activities.

**Conclusions:** For researchers with limited working experience in high TB/HIV burden countries, this review can provide valuable insights about barriers that may arise at different levels of the health system. For policymakers in high TB/HIV burden countries, this review offers strategies for improving the delivery of IPT (or any newer therapy regimen) for the prevention of TB. Based on our findings, we suggest initial and continuous stakeholder involvement, a focus on the efficient use and reinforcement of existing resources for health.

## Introduction

### Background

In 2019, 38 million people worldwide were living with the human immunodeficiency virus (HIV) [1]. An estimated quarter of the world’s population was latently infected with *Mycobacterium tuberculosis*; 10 million developed active tuberculosis (TB) [2, 3]. The fact that HIV-positive individuals have 20 times higher risk of developing active TB disease compared to HIV-negative individuals has been described as a lethal combination [4], which has transformed many low- and middle-income countries (LMIC) into countries with a high dual burden of TB and HIV [2]. The latest UNAIDS report continues to present TB as the leading cause of death among people living with HIV (PLHIV), accounting for around one in three AIDS-related deaths worldwide [1]. This review focuses on the implementation of two of the most important preventive therapies for PLHIV in countries with a high burden of TB/HIV: cotrimoxazole (CTZ) and isoniazid (INH).

Cotrimoxazole (CTZ) is a fixed-dose combination of trimethoprim/ sulfamethoxazole that combines broad antimicrobial activity with immunomodulatory properties. Besides preventing some AIDS-associated opportunistic diseases (*Pneumocystis jirovecci* pneumonia (PCP), toxoplasmosis), CTZ has shown to be successful in reducing malaria, severe bacterial infections, and mortality among PLHIV. As a result, World Health Organization (WHO) recommends cotrimoxazole preventive therapy (CPT) lifelong for PLHIV in resource-limited settings where malaria and, or severe bacterial infections are highly prevalent, irrespectively of their CD4 count. In settings were neither malaria nor severe bacterial infections are highly prevalent, CPT is recommended for PLHIV with severe or advanced HIV disease (WHO clinical stage 3 or 4, or CD4 count below 350 cells/mm^3^), and may be discontinued for those with evidence of immune recovery or viral suppression on antiretroviral treatment (ART). Adults, including pregnant women, children and adolescents with HIV, and HIV-exposed but uninfected infants, as well as PLHIV with active TB, are eligible for CPT, including those concurrently receiving ART [5].

Isoniazid (isonicotinylhydrazide or INH) is the most common type of TB preventive therapy. With its anti-mycobacterial activity, isoniazid monotherapy is prescribed to treat latent TB infection and prevent the progression from latent to active TB. WHO recommends at least six months of isoniazid preventive therapy (IPT) to people at risk of TB living in resource-constrained and high TB and HIV prevalence settings. PLHIV comprise a major risk group for TB, among which IPT has shown to reduce TB disease and mortality irrespective of receiving ART. Therefore, PLHIV unlikely to have active TB, are eligible for IPT. This includes HIV-positive positive adults including pregnant women, adolescents, infants aged under 12 months who are in contact with a TB case, and children older than 12 months who have no contact with a TB case. HIV-negative children aged under 5 years who are household contacts of people with bacteriologically confirmed pulmonary TB comprise another target group for IPT. Independent of the target group, only people with unknown or a positive tuberculin skin test (TST) unlikely to have active TB are eligible for IPT. In the absence of TST, patients should be screened for TB according to a clinical algorithm. Those who do not report any of the symptoms of current cough, fever, weight loss or night sweats are unlikely to have active TB and should be o□ered preventive therapy, irrespective of the degree of immunosuppression and including those who have previously been treated for TB [4].

### Rationale

Data from clinical trials and observational studies demonstrated that both preventive therapies are well-tolerated, highly efficacious, and cost-effective among PLHIV [6–10], resulting in WHO recommendations that were first adopted almost two decades ago [11, 12]. With increasing evidence supporting the benefits of the preventive therapies for PLHIV, WHO recommendations have been even expanded with regard to therapy duration and target populations indicated for CPT and IPT in today’s recommendations [5, 13]. Although efforts have been made by governments of high TB/HIV burden countries and their partners to translate these recommendations into national policy and practice, implementation of both preventive therapies has been challenging and coverage has been low [2, 14, 15].

### Objectives

The primary objectives of this review were to explore barriers to both preventive therapies reported across high TB/HIV burden countries (as per WHO [2]) and to generate explanatory knowledge of why their implementation has been so challenging. Additionally, this review aimed to compare both preventive therapies with respect to similarities and differences of barriers. A secondary objective was to identify strategies (facilitators) to improve the implementation of both preventive therapies. To identify relevant research, the broad question: “Which are the barriers to and facilitators for the implementation of preventive therapies (CPT, IPT) in countries with a high burden of HIV and TB?” was designed using FINER criteria [16] and the PICo framework: Population, Interest, Context (modified PICO) [17] (S1 Additional file). Barriers were defined as factors that limit, challenge or inhibit implementation, access, provision, delivery, or adherence to CPT or IPT. Facilitators were defined as factors that facilitate, support, encourage, or enable the implementation, access, provision, delivery, or adherence to CPT or IPT. In the published literature, the treatment of interest may be referred to as ‘preventive therapy’, ‘preventive treatment’, ‘prophylaxis’ or ‘prophylactic treatment’.

## Methods

### Protocol and registration

Following the Joanna Briggs Institute (JBI) methodology for mixed methods systematic review, we developed a protocol prior to undertaking the review [17, 18], which we registered and published in PROSPERO (CRD42019137778) (S2 Additional file). As outlined in section 8.5.1 of the JBI Reviewer’s Manual, we applied a convergent integrated approach to synthesis and integration [18]. We followed PRISMA guidance and reported our findings, according to the PRISMA checklist (S3 Additional file) [19].

### Eligibility criteria

Only peer-reviewed scientific papers meeting all of the following eligibility criteria were included: (1) papers reporting barriers and, or facilitators for either or both preventive therapies (CPT, IPT), sometimes in the literature also referred to as ‘preventive treatment’, ‘prophylaxis’, or ‘prophylactic treatment’. (2) Only studies conducted in high TB/HIV burden countries defined by WHO in the period 2016 to 2020 [2]; (3) published in English language; (4) until the 4th of September 2020 were eligible for this review. (5) Studied populations eligible were: (a) HIV patients or PLHIV. Although HIV-negative population groups may be eligible for IPT (e.g. household contacts of TB cases), HIV-negative population groups were not within the scope of this review. However, for CPT, HIV-exposed babies (HIV suspects) were eligible study populations, as well as patients co-infected with TB and HIV. Other studied populations eligible were (b) healthcare providers, also referred to as health professionals; (c) caregivers (i.e. parents or guardians of child patients involved in the preventive therapy collection or drug administration process); (d) any other stakeholder identified as influential in the overall implementation process of either IPT or CPT, and (e) countries defined by WHO as high TB/HIV burden countries [2]. Since this systematic review aims to analyse studies reporting primary data, we excluded editorial comments and systematic reviews. However, we screened reference lists of systematic reviews for original research eligible for this review. We included studies conducted in multi-sites or multiple countries only if barriers/ facilitators were separately analysed and reported per site or country. Because of the limited number of published qualitative studies on this topic, we included primary studies of any design (i.e. qualitative, quantitative, multimethod and mixed methods studies).

### Information sources and search

In February 2018, we systematically searched the electronic databases MEDLINE®, Web of Science® and Scopus® for original articles using the search terms presented in (S4 Additional file). We repeated the search in September 2020 to identify additional literature published after February 2018 and updated our review [20]. The full electronic search strategy is available for each database search (S4 Additional file). We searched references of systematic reviews only if the systematic review was relevant to the study question.

### Study selection

Following the search, PM collated and uploaded all identified citations into Endnote X9 reference management software, and removed all duplicates. We both independently screened and assessed all titles and abstracts against the predefined eligibility criteria for this review. Non-relevant studies and studies reporting from countries other than the thirty high TB/HIV burden countries were excluded during this initial title and abstract screening process. When the decision on exclusion was not clear, we included the study for full-text screening. PM retrieved the full-text of all potentially relevant studies. Eventually, we both independently assessed the full-text of the remaining studies against the eligibility criteria. Studies that did not meet all eligibility criteria were excluded, and reasons for exclusion were recorded. Studies that met all eligibility criteria were included. We compared our decision (i.e. inclusion/ exclusion and reason for exclusion) for each of the selected studies, discussed their full-text and resolved any disagreement concerning our decision through discussion.

### Data collection process and data items

We developed a data extraction table which we initially tested on three studies to ensure that all relevant data items could be extracted. PM extracted the following data items from each included study: first author, year of publication, geographic origin (one or multiple countries defined as high burden country for TB/ HIV), context (e.g. urban, rural), study type, study subject(s) of interest, sample size of study subjects for each data collection approach, study aim(s), data collection approach (e.g. interviews, record review), and findings related to the review question (i.e. barrier(s) and, or facilitator(s) for CPT, IPT, or both). LVL checked the extracted data items for accuracy and added or modified data items were necessary. We compared the data we individually extracted and resolved any disagreement through discussion.

### Data transformation

As outlined in Section 8.5.1 of the JBI Reviewer’s Manual, we extracted findings (barriers and facilitators) from quantitative, qualitative and mixed methods studies. Qualitative findings including the qualitative component of mixed methods studies were extracted as presented in the original research paper (e.g. themes, corresponding illustrations, paragraphs of textual description). We transformed quantitative findings, including the quantitative component of mixed methods studies, into textual description disregarding the effect size [18]. For example: “A quantitative study reported a twenty per cent decrease in cotrimoxazole stock-outs (p<0.05) among health facilities that had employed trained pharmacy personnel, compared to health facilities without any trained pharmacy personnel.” From this fictitious example, the following information would have been extracted: (1) Finding: Facilitator for CPT; (2) Description: employment of trained pharmacy personnel has shown to decrease stock-outs. Finally, we merged qualitative findings and transformed study findings together into one data set.

### Risk of bias in individual studies

Due to the methodological diversity of the studies included in MMSR’s, the selection of an appropriate tool that allows assessing the methodological quality of included studies is particularly challenging for this type of review. For the critical appraisal of methodological quality (internal validity) of the included studies, we selected the “Mixed Methods Appraisal tool” (MMAT), version 2018 [21]. The MMAT is a critical appraisal tool designed for the appraisal process of mixed methods reviews because it specifically addresses items related to the various study designs being evaluated in a single tool [21]. Methodological quality criteria to be rated in the MMAT are specific for (a) qualitative research, (b) randomised controlled trials, (c) non-randomised studies, (d) quantitative descriptive studies, and (e) mixed methods studies (S5 Additional file). As reported by the Cochrane Qualitative and Implementation Methods Group, this tool has been used widely in systematic reviews and has the advantage of being able to assess interdependent qualitative and quantitative elements of mixed-methods research [22]. We independently assessed methodological quality based on the details published in the studies included in our review using the MMAT [21]. We resolved any disagreement in rating through discussion. To preserve the transparency of the risk of bias assessment, we documented all individual ratings for each study included in this review. For studies that failed to meet more than one quality criteria, we discussed whether to exclude the study. However, we also considered that our study’s focus was on qualitative findings and on the generation of explanatory knowledge of why the implementation of both preventive therapies has been so challenging. Due to the risk of excluding insights relevant for a good understanding of the phenomenon under study, which may only become apparent at the point of synthesis, bias was toward inclusion [23, 24].

### Qualitative synthesis

We applied framework synthesis, a highly transparent and deductive approach recommended for the synthesis of evidence on complex interventions [25]. This approach allows combining elements of critical realistic and subjective idealistic epistemology. We analysed our data set using MAXQDA Analytics Pro software (Release 18.1.1.), iteratively coding and sub-coding the extracted results. Both researchers individually defined, cross-checked, discussed and refined the code system. We resolved disagreement through discussion.

Inspired by Getahun et al. (2010) [26], we applied a priori defined modified version of the WHO’s Framework for action [27] to analyse and present barriers to preventive therapies. First, we assigned each extracted result (i.e. barrier) to one of the seven health system components based on the level at which they hindered implementation. Health system components include ‘Patient & community’, ‘Health providers’, ‘Clinical information’, ‘Leadership & governance’, ‘Pharmaceutical management’, ‘Service delivery’ and ‘Financing’. We slightly modified three of the original component names and added the component “Patient & community” as the seventh component to the existing framework. Second, we thematically analysed each barrier considering its contextual description. Third, to summarise the comprehensive barrier descriptions, we applied metasummary. Metasummary is a quantitatively oriented aggregation of qualitative findings first proposed by Sandalowsky, Barroso and Voils (2007) [24, 28]. Mapping of key barriers across the seven health system components facilitated comparison of similarities and differences of barriers between both preventive therapies [25].

For facilitators, we applied ‘the preventive therapy cascade’ as framework for evidence synthesis. After familiarising ourselves with the extracted data set, reading and re-reading the identified facilitators and exploring underlying patterns, we found that facilitators generally aimed to enhance specific activities (e.g. providers’ prescribing practices, patients’ adherence) along a series of steps involved in the implementation of preventive therapies. Eventually, the preventive therapy cascade emerged as most relevant framework for the synthesis and presentation of our data, featuring each step along the preventive therapy cascade with all its bottlenecks. We assigned each extracted result to one or multiple steps along the cascade, followed by thematic analysis, inductively grouping facilitators into overarching themes [25].

### Risk of bias across studies

Published studies are more likely to report statistically significant results compared to unpublished studies. Hence, publication bias compromises reviewers’ ability to assemble a complete body of evidence that is not biased towards “positive findings”. Although publication bias is primarily a concern for systematic reviews evaluating an intervention’s effect, it is debatable whether the consequences of such bias is as potentially serious in qualitative evidence synthesis [29]. Dissemination bias (which encompasses publication bias) is gaining increasing attention in qualitative evidence synthesis. More research is required on how to assess dissemination bias in the context of qualitative evidence syntheses [30].

### Additional analysis

Due to the complexities associated with findings derived from both streams of evidence and the impact of data transformation and integration on the grading process, an assessment of the certainty of the evidence is currently not recommended for MMSR’s [18]. However, we analysed heterogeneity of the countries and the study subjects represented in this review and discussed potential limitations resulting from a heterogeneous representation of these variables.

## Results

### Study selection

The PRISMA Flow Diagram presents the number of papers included throughout the selection process, alongside with the reasons for exclusion (Fig 1).

**Fig 1.**
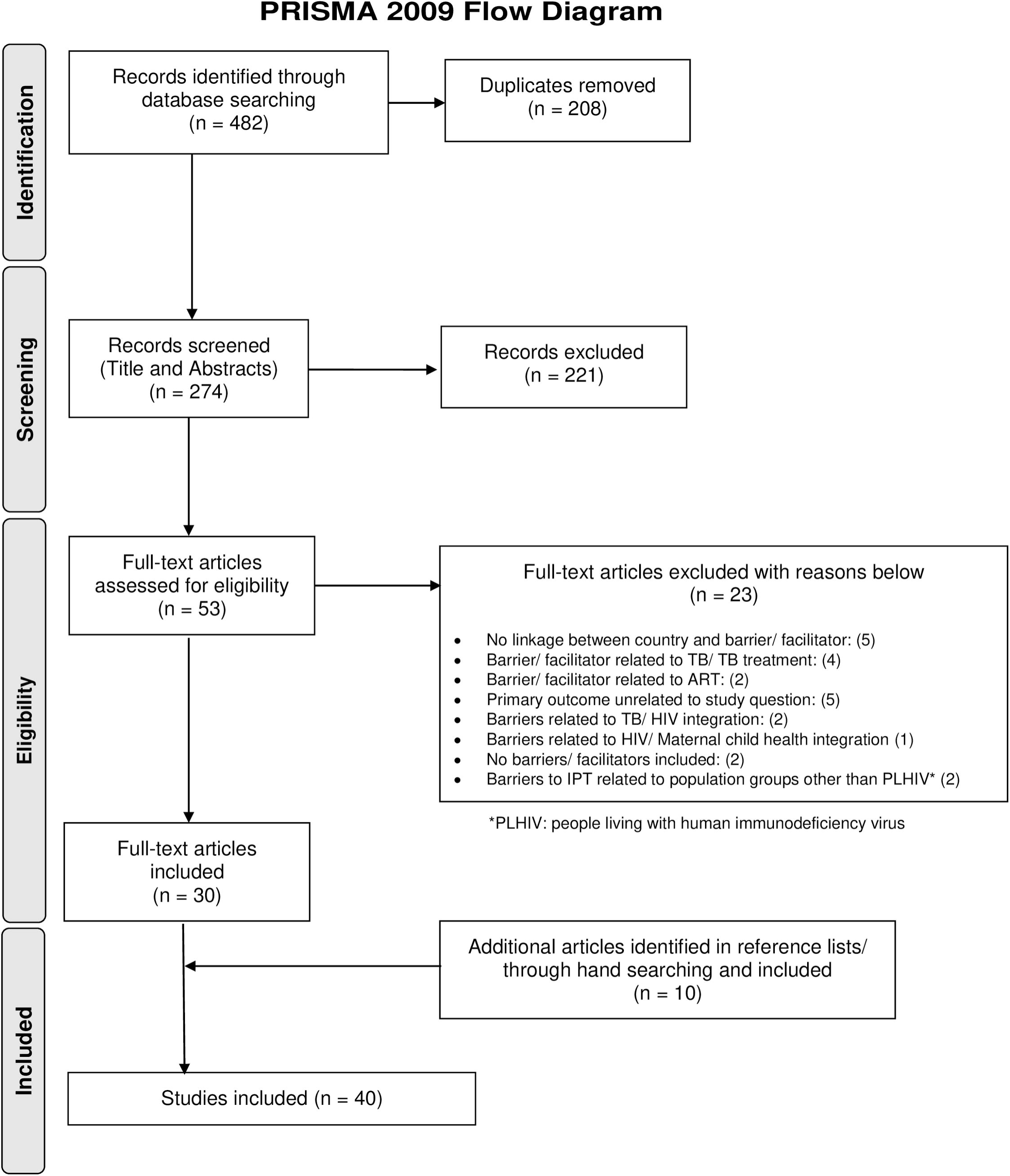
PRISMA Flow Diagram.

### Description of studies included in the systematic review

Forty studies were included for review, fourteen related to CPT [14, 15, 31–42]; twenty-eight related to IPT [14, 38, 43–68], including two studies that reported findings on both preventive therapies [14, 38]. While facilitators were reported in all studies, barriers were not presented in three studies [32, 40, 41], all related to CPT. Comparing the year of the first publication included for both preventive therapies, we found that the first study included on IPT has been published fourteen years before the first study on CPT (1995, 2009) [38, 44]. All studies related to CPT were carried out in WHO African Region [14, 15, 31–42], while studies related to IPT also included WHO South East Asia Region [47, 61, 62], WHO Western Pacific Region [47] and WHO Region of the Americas [46, 47] (Fig 2).

**Fig 2.**
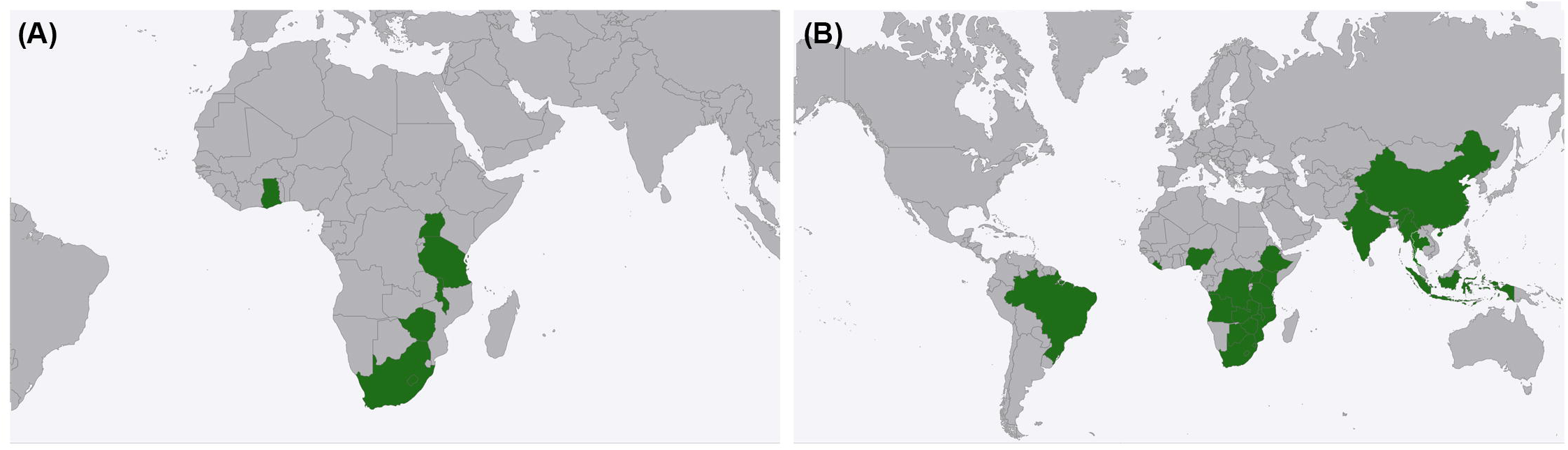
Regional representation of countries included in this review. (A) High TB/HIV burden countries represented in studies reporting barriers or facilitators for cotrimoxazole preventive therapy (n= 7): Ghana, Malawi, South Africa, Tanzania, Uganda, Zimbabwe and Lesotho (published between 2009 and 2018). (B) High TB/HIV burden countries represented in studies on IPT (n= 22): Angola, Botswana, Brazil, China, DRC, Ethiopia, India, Indonesia, Kenya, Lesotho, Liberia, Malawi, Mozambique, Myanmar, Nigeria, South Africa, Swaziland, Tanzania, Thailand, Uganda, Zambia and Zimbabwe (published between 1995 and 2020). Source: Own visualization of review data: https://www.amcharts.com/.

Among the thirty countries defined by WHO as countries with a high burden of TB and HIV [2], we identified research from twenty-three countries (77%) reporting barriers or facilitators to either or both preventive therapies. Countries were disproportionally represented. The majority of studies were conducted in South Africa (n = 11) [37, 42, 47, 50–52, 54, 63–65, 67], Uganda (n = 9) [31-33, 38-40, 44, 56, 66], Tanzania (n = 5) [15, 35, 36, 47, 60], Kenya (n = 4) [45, 47, 49, 68], Zimbabwe (n = 3) [33, 34, 47], Ethiopia (n = 3) [47, 53, 58], and Nigeria (n = 3) [43, 47, 57]. Another fifteen countries were represented in only one or two studies. All studies, except three [33, 47, 49] based their findings on data collected in one single country. Half of the studies missed to mention the context where the study was carried out. Among the twenty studies that reported the context, ten were conducted in urban areas [33, 35, 36, 41, 44, 45, 49, 58, 67, 68], eleven in rural areas [33, 38, 39, 42, 45, 49, 50, 55, 62, 63, 66] and six in intermediary areas [33, 38, 42, 44, 51, 65] (i.e. peri-urban, sub-urban, and semi-urban context). Some studies were conducted in multiple contexts.

Patients (n = 24) [31, 32, 35, 36, 38, 43, 44, 46, 48–52, 54, 55, 58–66], health providers (n = 14) [14, 15, 36, 38, 39, 45, 46, 51, 53, 54, 62, 63, 67, 68], and health facilities (n = 14) [15, 33, 36, 37, 39–42, 44, 45, 49, 56, 57, 67] were the study subjects of interest most frequently represented in this review, with at least one of the three study groups included in thirty-eight studies (95%). Few studies included other study subjects, explicitly caregivers [15, 34], community members [38], other stakeholders [14, 45, 46, 56], districts [38], and countries [47]. Table 1 shows the main characteristics of the studies included in this review. Additionally, we provided a more detailed presentation of study characteristics separately for studies on CPT and IPT (S6 Additional file).

**Table 1.**
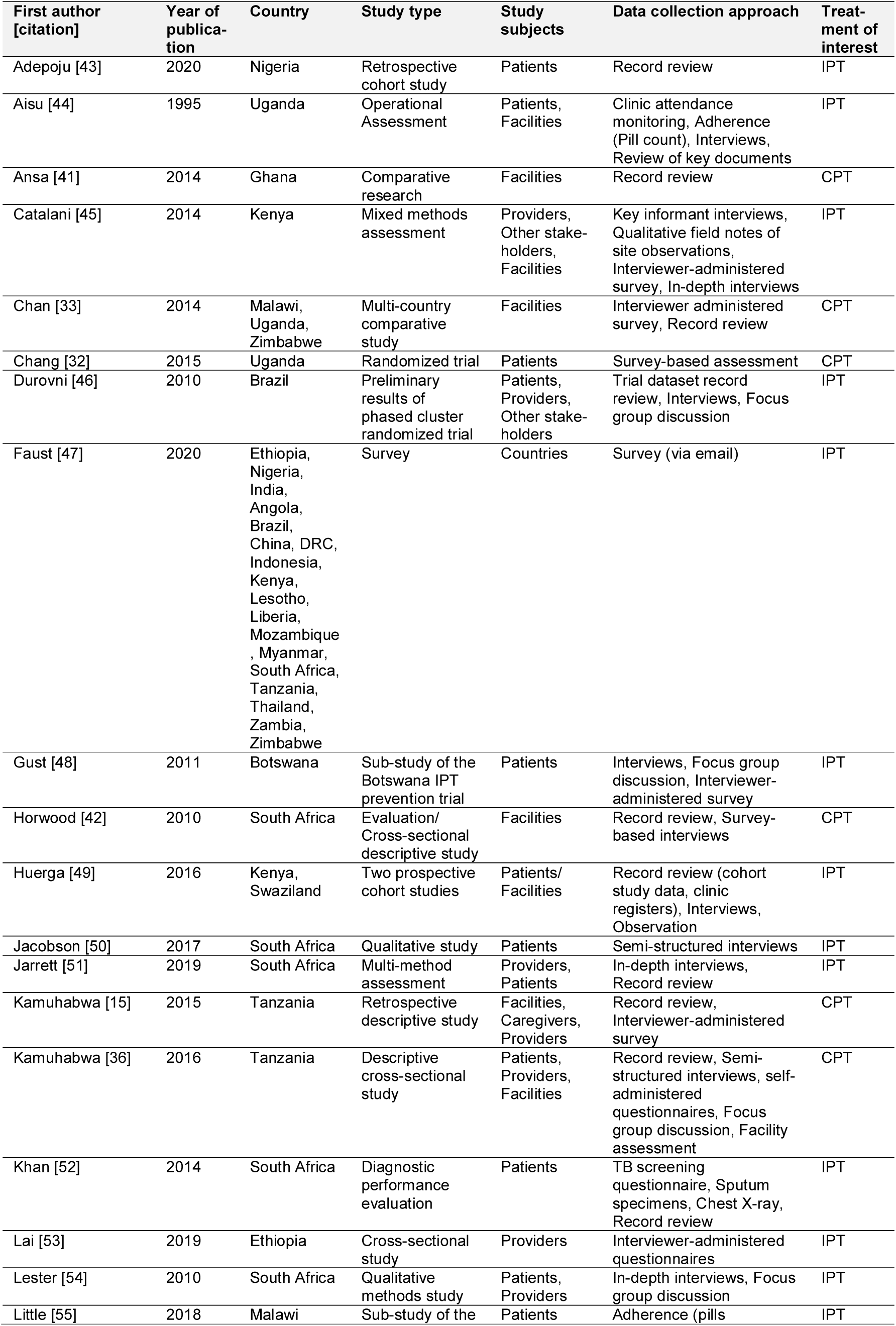
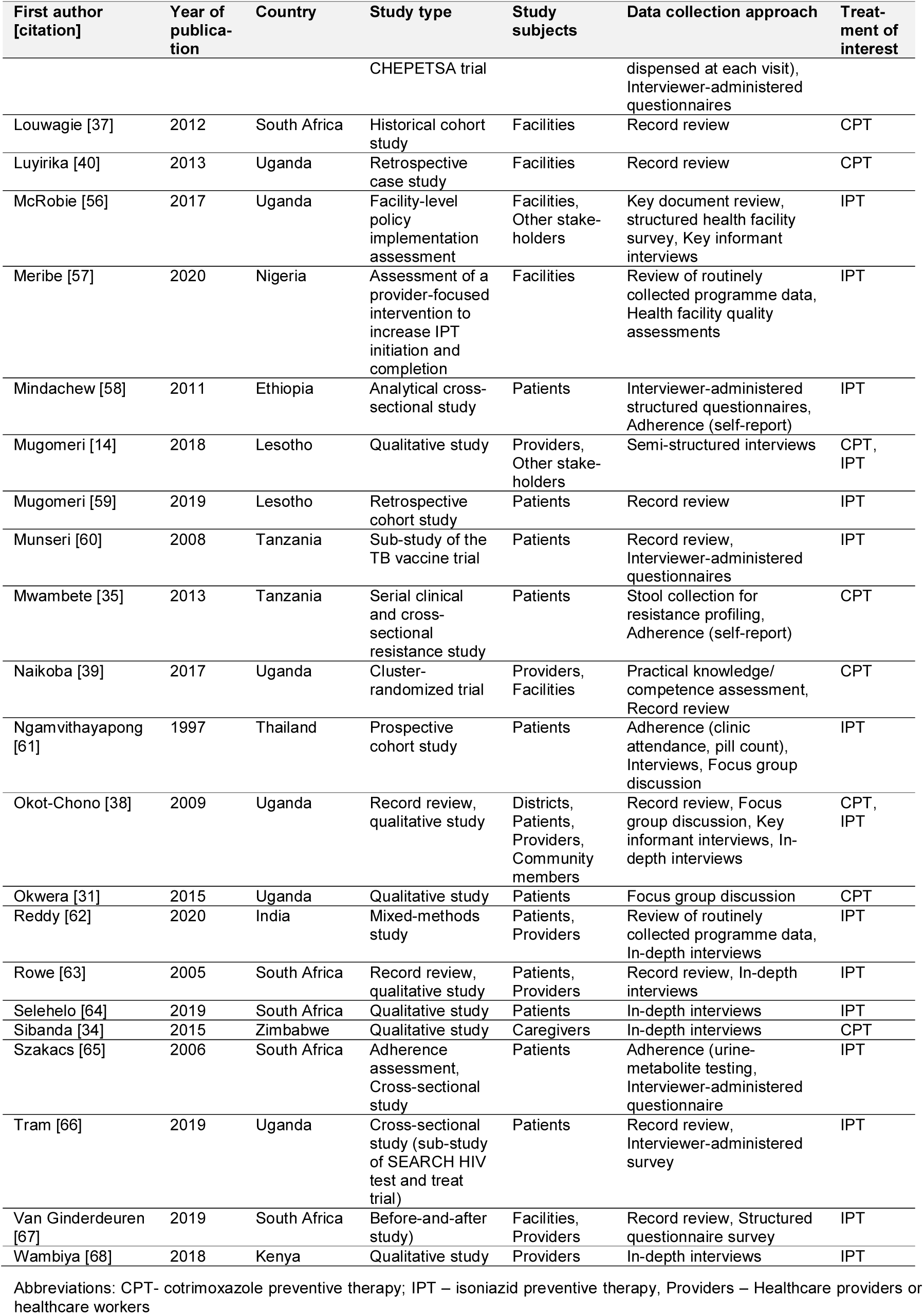
Main characteristics of the studies included in this review.

### Risk of bias

We assessed the methodological quality of each study individually (S7 Additional file). First, we identified the categories of study design using the MMAT algorithm and then appraised each study against the corresponding methodological quality criteria [21]. Among all studies reporting findings for CPT and IPT, more than half (57%, 50%) applied quantitative methods [15, 32, 33, 35, 37, 39, 41–44, 47, 52, 53, 55, 57–61, 65–67], followed by studies that applied a combination of methods (21%, 28%) [36, 38, 40, 45, 46, 48, 49, 51, 56, 62, 63] and those that applied qualitative methods (21%, 18%) [14, 31, 34, 50, 54, 64, 68], for CPT and IPT, respectively. Across all studies that combined quantitative and qualitative methods, most (80%) presented no methodological attempt to integrate phases, results, and data from both streams of evidence [36, 38, 40, 46, 48, 51, 56, 63]. Therefore, nine studies were categorised as multimethod studies [36, 38, 40, 46, 48, 49, 51, 56, 63] and two as mixed methods studies [45, 62]. According to the MMAT checklist, many included studies (n = 29) have methodological limitations in at least one of the quality criteria evaluated. Qualitative studies (including the qualitative component of mixed methods and multimethod studies) frequently failed to address and justify the qualitative approach applied (e.g. ethnography, phenomenology) [31, 34, 36, 38, 40, 46, 51, 54, 56, 62, 63]. Quantitative non-randomized studies frequently failed to address whether the intervention or exposure occurred during the study period as intended [41, 43, 58–60, 66]. Some studies also missed to provide reasons why certain eligible individuals chose not to participate [55, 60, 65]. Similarly, the risk of non-response bias was frequently not addressed across quantitative descriptive studies [15, 35, 42, 47, 57]. Overall, many studies’ research question(s) lacked clarity and focus [32, 35, 38, 45, 56, 59]. We identified seven studies with methodological limitations in more than two of the quality criteria evaluated [35, 38, 40, 45, 46, 56, 62]. However, to gain a broader understanding of the barriers and facilitators to PT’s, we did not exclude any study from our analysis regardless of the methodological limitations.

Selective reporting of studies and findings in primary research may have introduced a risk of bias across studies. Similarly, publication bias may have led to a systematic distortion of our understanding of the phenomenon of interest, solely because specific findings are less easily accessible or available. Bias introduced through studies or findings that are not available (e.g. time-lag bias, truncation bias) could not be controlled. To address potential biases that result from studies being less easily accessible or available, we highlighted the absence of specific and relevant evidence in the limitation section of this review.

### Barriers to preventive therapies

#### Patient and community related barriers

**Adverse reactions**, side effects or undesirable reactions were frequently assessed in the included studies [14, 15, 31, 34–36, 46, 48, 53, 58, 60–64, 66–68]. Many of these studies including two that based their findings on IPT trial data [46, 48] either found that both preventive therapies were generally well tolerated, that the majority of patients had not experienced any side effects, or both [15, 31, 35, 46, 48, 60, 63, 67]. However, we identified the following adverse reactions among a minority of patients.

Regarding CPT, few patients reported itches and headaches [35], nausea and vomiting [15, 35, 36], skin lesions or oral wounds [35], skin rashes [31, 35, 36], and hypersensitivity reactions [15]. Additionally, heartburn, throat dryness, dizziness, fatigue, appetite loss and numbness were reported by few HIV-positive adults [35]. In rare cases, severe cutaneous reactions or burning sensations (e.g. Stevens-Johnson syndrome) required therapy interruption [31, 36]. In this case, some received dapsone as an alternative therapy for the prevention of opportunistic infections [31].

Regarding IPT, few patients reported headaches [46], arthralgia [46, 48], pruritus [46, 64], dizziness or fatigue [48, 62, 64], gastrointestinal disturbances [46, 62, 64], nausea or appetite loss [48, 60, 62], vomiting [62, 64], weight loss [48] and darker skin complexion [64]. The most serious, very rarely reported adverse reactions to INH were seizures, psychiatric symptoms [48, 64], (peripheral) neuropathy [60, 62] and mild to moderate hepatoxicity [46, 60]. Very few patients experienced adverse reactions that required therapy interruption [48, 60, 63]. After stopping IPT, liver function abnormalities resolved [60].

Co-administration of other drugs was believed to have contributed to adverse reactions [35, 36, 48]. The simultaneous administration of ART, TB treatment and other drugs (e.g. to treat hypertension, diabetes, asthma) was common among PLHIV [35, 36, 48, 64]. One patient reacted to concurrent CTZ and INH, two weeks after initiating ART [14]. Concurrent use of CTZ and nevirapine may have contributed to rash and leukopenia [36]. Besides, pyridoxine shortages and patients’ poor nutritional status were believed to have contributed to INH-associated neuropathies [62].

Our review revealed that side effects influenced patients’ and health providers’ views, attitudes and practices regarding the use and prescription of preventive therapies. Some had heard about side effects and feared experiencing such reactions in the future [31, 34]. Three studies suggested that fear of potential side effects alone contributed to patients’ non-adherence to IPT [58, 60, 68]. However, seven papers also reported actual or perceived side effects as the reason for patient-initiated non-completion or lacking adherence to IPT [58, 60–62, 66, 68] and CPT [36]. Tram and colleagues (2019) found that patients who reported side effects, and those who believed INH was dangerous for their health had lower odds of completing IPT [66]. Eventually, patients who stopped taking IPT may have discouraged others from initiating the preventive therapy [68]. For IPT, side effects and contraindications were also acknowledged by health providers [53, 62, 67]. For example, health providers interviewed in rural India believed that side effects were an important reason for non-completion of IPT [62]. Although health providers believed that IPT was important for PLHIV [53], due to side effects and contraindications (e.g. active liver disease, history of adverse reactions), sometimes the therapy was not prescribed [53, 67].

Instead of discontinuing CPT, some patients independently reduced the prescribed doses as a measure to manage side effects [15, 35, 36]. Parents reported reducing their infants’ doses of CTZ if they were unwell or perceived side effects [15]. Similarly, HIV-positive pregnant women reported skipping doses of CTZ for the same reason or taking CTZ at night to cope with perceived side effects [36].

**Lacking financial and organisational feasibility** frequently emerged as barrier theme [15, 34, 38, 44, 46, 48, 50, 54, 58, 60–63, 65]. Evidence synthesis revealed that long distance to the clinic [44, 60], lacking transport [15, 44], and the with transportation associated costs [44, 46, 48, 50, 54, 63] routinely comprised an obstacle for patients to access therapy and care. Women and adolescents were reported to lack control over economic resources, leaving the decision to attend the clinic with their parents or husbands [63]. Jacobson et al. (2017) [50] argued that a major barrier to IPT had been patients entering HIV care. The researchers suggested that once linked to care (i.e. adherent to ART), the marginal cost for patients to take IPT was low in the South African study setting [50]. Our review found that frequent stock-outs at public health facilities caused therapy interruptions among patients who could not afford to buy INH or CTZ at the community pharmacy [15, 34, 38, 65]. Research from Uganda and Brazil suggested that TB screening incurred additional costs for patients, including fees for chest radiology, transport for three consecutive smear checks [46], as well as consultation fees [38]. HIV-positive clinic attendees in South Africa frequently reported hunger and lack of food security. Lacking food appeared to be a barrier to IPT because patients believed they could not take the therapy without food intake [63]. Health providers in Karnataka, India, listed ‘lack of social support and access to social protection schemes’ as a reason for patients’ non-completion of IPT [62]. In terms of organisational feasibility, studies from Botswana and South Africa showed that patients were facing competing priorities, with the need to choose between spending a full day waiting at the clinic, dedicating the time to their work [48, 50, 54] or addressing other family members’ needs [50, 63]. Studies from Ethiopia, Botswana, Uganda and Thailand showed that patients did not access treatment or care, when they relocated [48], moved far away [44] or were seeking work in other provinces [35, 50, 58, 61]. Parents and caregivers reported lack of a refrigerator at home as challenge for storing paediatric formulations of CPT [15].

**Knowledge gaps and misperceptions** regarding both preventive therapies were barriers for patients’ retention in care, therapy adherence and completion [15, 31, 38, 44, 48, 50, 51, 54, 61–64, 67, 68]. Patients in Uganda thought that CTZ is an analgesic drug, that concurrent use of CTZ and ART, or TB treatment is contraindicated, and that CTZ was a treatment for HIV-positive patients [31]. In Tanzania, parents and caregivers reported difficulties in understanding how to use CPT. They particularly struggled to prepare the CTZ suspension for their children using crushed tablets when the paediatric formulation was unavailable [15]. Patients in Thailand thought that INH was prescribed to reduce HIV blood concentration or prevent AIDS-related complications [61]. Patients in South Africa believed that INH would alleviate symptoms, leaving asymptomatic patients unconvinced [50]. Three studies reported that HIV-positive adults were not aware of the desired benefit of IPT in preventing TB [50, 54, 61], and some patients had never heard of IPT [61]. Not knowing the benefit, seemed acceptable for some patients, as shown in three studies, which revealed that patients took IPT without understanding its purpose or duration [50, 61, 64]. However, most studies suggested that low patient knowledge about IPT is rather harmful for its implementation [44, 48, 50, 61, 63]. Knowledge gaps can create wrong assumptions, mislead patients from the appropriate medical advice, or promote seeking information from other sources [61]. Five studies reported insufficient patient information was a reason for missing pills, therapy interruptions, or non-compliance to clinic appointments among PLHIV [44, 48, 50, 61, 63]. Qualitative research conducted in South Africa revealed that some patients interrupted therapy, thinking they no longer needed IPT when INH refills were accidentally omitted [50]. This patient belief illustrates that knowledge gaps among PLHIV also undermine patients’ potential to generate demand for IPT [62]. Patients in Uganda and South Africa felt better and did not understand the need to return to the clinic, or to take IPT when they did not have TB, suggesting they lacked understanding of the concept of prevention [44, 51, 63]. Health providers interviewed in Kenya, South Africa and India also acknowledged this problem [51, 62, 68]; some patients refused to take ‘TB medicine’ [INH] without having TB [51, 68]. Others did not value the preventive effect of IPT because of the low perceived risk of TB and lack of symptoms [62]. Health providers reported that information materials were lacking or inadequate to educate PLHIV, suggesting there was a lack of social campaigns, and consensus regarding patient education activities [62, 68].

**Patients’ lacking motivation** [15, 44] was identified for both preventive therapies alongside with a number of explanations [15, 31, 34, 38, 44, 48, 54, 58, 62–64, 68]. Patients reported they struggled with the need for too many clinic visits, on the one hand, due to a lack of same-day services [38], on the other hand, due to the long waiting time prior to receiving services [38], as well as the long preventive therapy duration [58, 68]. Patients reported they were too busy to take IPT [44, 63] and also struggled with the daily pill burden [31, 48, 54, 62, 64, 68], which was exacerbated through co-medication [54, 64]. Patients revealed they received ART, medications for hypertension and diabetes, or asthma in addition to IPT [64]. Pill burden affected patients adherence, providers believed [62, 68]. Some patients feared adding to the pill burden [62]; others completely declined to take IPT due to the high number of pills prescribed to PLHIV [68]. Studies from Uganda and South Africa reported that patients presumed side effects [44], found IPT unnecessary [63] or were just not interested in taking IPT [44]. Research from Zimbabwe revealed that one mother felt let down by pharmaceutical interventions and stopped giving CPT after her baby tested HIV-positive [34].

**Forgetfulness** has been frequently reported as a reason for patients’ non-adherence to preventive therapies [35, 48, 58, 65].

**Patients’ HIV denial, religion and competing medicinal approaches** impeded patients acceptability of preventive therapies [34, 36, 44, 48, 61, 63, 66]. Patients’ denial of HIV was a barrier to patients’ retention in care [61], and consequently, their preventive therapy uptake. Mothers who tested HIV-positive in Zimbabwe associated their diagnosis with impending death and reported that they preferred to ignore HIV and not give CPT to their babies when they felt depressed [34]. Non-compliant adult patients interviewed in Uganda and Thailand were not accepting their HIV status and thus, never returned to the clinic, or only months later [44, 61]. Research from Uganda suggested that after receiving their test result patients were still in the process of coping with their HIV diagnosis [44]. Religion and competing medicinal approaches were barriers identified on the sub-Saharan continent. Three studies suggested an influence of religion as an explanation for HIV denial [36, 48, 63]. Interviews with HIV-positive pregnant women in Tanzania revealed that people diagnosed with HIV were encouraged to pray that God will cure their infection, while born-again Christians were advised to stop taking CTZ [36]. Self-reported reasons for non-adherence and loss-to-follow up in the Botswana IPT trial also included religious beliefs [48]. In South Africa, a study participant who interrupted IPT reported that members of the church could not combine clinic medication with church tea. Similarly, people believed it was prohibited to take clinic medication with traditional medicines [63]. According to Rowe et al. (2005), HIV was perceived as incurable by western medicine, while traditional healers were perceived as able to cure HIV, suggesting two competing medicinal approaches [63]. Survey-based research conducted in Uganda found that among non-completers of IPT, some patients took traditional medicines ‘to prevent becoming sick with TB’ [66].

**Stigma and fear of rejection or discrimination** remains a barrier for HIV care and consequently for the implementation of both preventive therapies [34, 36, 38, 44, 48, 50, 60, 61, 63, 65, 66]. Cotrimoxazole and IPT were linked to HIV [36, 60, 63, 66], which although a common infection in the countries included in this review, was frequently associated with stigma and fear of rejection [34, 60, 63] or discrimination [48, 60, 63]. Studies found that women feared separation from their spouses and families because of their diagnosis [34, 60]. Although mostly reported by women, in-depth interviews with HIV-positive mothers in Zimbabwe, suggested that men also feared separation from their wives when testing HIV-positive [34]. HIV-positive people were laughed at or ridiculed, suggesting a lack of understanding of HIV within the community [34]. Among men, HIV was perceived as emasculating [34]. These HIV-positive patients’ experiences explain their frequently cited unwillingness to disclose their HIV status [34, 36, 50, 60, 61, 63]. One study reported that a woman disguised her HIV medication in a container for headache medication [50]. With the intention to keep their diagnosis secret, it has been difficult for women to take CPT at their crowded homes or administer it to their babies [34, 36]. Study participants in northern Thailand did not allow nurses to visit their homes because they wanted to keep their HIV status confidential [61].

The fact that patients feel the need to keep their HIV status confidential is a concern because it is likely to contribute to the non-completion of preventive therapy. According to Tram and colleagues (2019) [66], ‘not talking to friends or family about IPT’ was associated with non-completion of IPT’ while ‘feeling comfortable taking IPT in front of a friend’ was an independent predictor for IPT completion among HIV-positive study participants in rural Uganda. Lacking privacy and confidentiality at the health facility made it difficult for women to hide their diagnosis [44]. People attending the overcrowded health facility took the risk of being seen by their spouses and the community when collecting CTZ prescription refills [34] or sitting in the open waiting area [38]. Fear of stigma within the household, caused challenges to therapy adherence [58], whereas perceived at the health facility level, hindered patients’ retention in care [60].

**Influence of relatives and friends** may be an important factor for the implementation of both preventive therapies [31, 34, 36, 44, 48, 60, 62, 63]. Although social and family support seemed to positively influence some patients’ therapy adherence [63], compliance with the prescribed therapy regimen was at threat when patients lacked this support [34, 44, 60, 63, 66]. Sometimes, relatives and friends adversely influenced patients’ compliance with their clinic appointments, retention in care, therapy adherence or completion. Some patients felt discouraged by their relatives, who disapproved or sabotaged their treatment adherence [31, 44, 48, 60]. Interviews with HIV-positive adults in Uganda and Tanzania revealed that some had been discouraged by their spouse or relatives to take IPT [44, 60]. Similarly, HIV-positive adults in Uganda were advised by their relatives to discontinue CPT [31]. Interviews with HIV-positive mothers in Zimbabwe suggested that some husbands opposed CPT. They refused to accept their wife’s HIV status and treatment, as well as the administration of CPT for their baby [34]. Research from Botswana reported stolen pills, ‘the boyfriend threw out the pills’, or ‘the sister flushed them down the toilet’, as reasons for trial participants discontinuation of IPT [48]. Some wives reported their husbands stole their CTZ to take themselves [34]. Health providers in Tanzania reported that HIV-positive pregnant women sometimes discarded CTZ, or shared it with their family members [36].

**Socio-demographic, lifestyle and clinical factors** appeared to influence peoples’ use of IPT (i.e. their chances of initiating, receiving, completing or adhering to IPT) [14, 46, 48, 55, 58–60, 62, 66, 68]. Studies suggested that people’s age (being younger) [48, 55, 59, 62, 66], low literacy or educational level [60, 62], their lifestyles [46], including drinking alcohol [48, 62], negatively influenced patients’ use of IPT. With respect to sex, we identified contrasting findings; five studies suggested that being female was positively associated with the use of IPT [48, 55, 59, 61, 62], while one study from Tanzania suggested vice versa [60]. Although there was some consensus that younger age was positively associated with the use of IPT) [48, 55, 59, 62, 66], some subgroups lagged behind. In particular, children (and adolescents) had a lower probability of initiating (or receiving) IPT compared to their adult counterparts [14, 59, 62]. Pregnant women (unaware of their HIV status) attending antenatal care services as late as 36 or 38 weeks of pregnancy [14, 48], people aged over 65 [62] and spouses of PLHIV also lagged behind [14, 62]. A higher education level seemed to improve the chances of IPT initiation [62] and completion [60]. A mixed-methods study from India found that compared to those who were illiterate, patients with formal education had a higher chance of IPT initiation [62]. The authors argued higher IPT completion rates among participants with education above the secondary level could have been due to patients’ better understanding and comprehension of instructions [60]. Health providers interviewed in Kenyan cities thought non-adherence was more likely across patients with a poor immunological and clinical state and patients on second-line ART [68]. Similarly, results of a retrospective cohort study carried out in Lesotho suggests that patients with WHO clinical stage 4 were more likely to interrupt IPT than those with clinical stage 1, 2, or 3 [59]. Those bedridden and with a poor clinical state may be interrupting IPT due to difficulties in reaching the distant hospital [59].

**Concern about the efficacy of CPT** resulted from the steady evolution of antibiotic-resistant bacteria. Research from Dar es Salaam, Tanzania, attempted to investigate the incidence of faecal *E. coli* resistance to CTZ among HIV patients through in-vitro resistance testing and found considerably high antibacterial resistance. The authors concluded resistance concerns and suggested reconsidering the use of CPT in Tanzania [35]. Our review also identified factors that may have contributed to the evolution of CTZ-resistant bacteria. Factors presented by Okwera et al. include poor adherence to CPT and doctors’ loose prescription practices [31]. Besides, the availability of antibiotics without prescription (in pharmacies, parks, or bus stations) may have promoted resistance to CTZ through irrational use and self-medication [31, 35]. We did not identify resistance concerns or scepticism about the continuous efficacy of CPT among health providers.

#### Health provider related barriers

**Shortage of health providers** working at public health facilities emerged as a barrier for both preventive therapies [14, 36, 38, 43, 45, 46, 51, 56, 62, 64, 67, 68]. However, except from one study that referred to CPT [36], all other studies reported staff shortages as an obstacle for the implementation of IPT [14, 38, 43, 45, 46, 51, 56, 62, 64, 67, 68]. Shortage of health providers was found in a range of countries including South Africa [51, 64, 67], Uganda [38, 56], Kenya [45, 68], Tanzania [36], Nigeria [43], Lesotho [14], Brazil [46], and India [62]. Regarding CPT, Kamuhabwa et al. (2016) [36] reported a shortage of pharmaceutical personnel at the twenty-six antenatal clinics included in their study in Tanzania. Regarding IPT, three studies from Uganda and Kenya explicitly stated health provider shortages with few providers trained in IPT [68] or co-management of TB-HIV [38, 56], and altogether too few health professionals to implement HIV policies [56]. A disproportional provider-patient ratio was reported, suggesting the number of health providers did not allow the additional tasks required to deliver IPT [62, 64, 68]. Studies from Brazil and Kenya, aimed at exploring doctors attitudes to IPT, described doctors’ pressure to see as many patients as possible (i.e. twenty to forty patients per provider before taking lunch), thus indirectly reported provider shortages [45, 46]. Interviews with HIV and TB doctors regarding their work, motivation and capacity to implement IPT in HIV clinics in Brazil, showed that they were too busy, tense and solitary [46]. Although doctors interviewed in Kenya believed in the importance of IPT for PLHIV, they found it difficult to manage TB priorities among many other priorities [45]. Two studies from Uganda further advised that working conditions (i.e. low salaries, heavy workloads) and missing incentives (i.e. career development prospects) were demotivating for health providers, leading to high staff attrition and consequently loss of knowledge [38, 56]. Similarly, research from South Africa reported high staff turnover, which seemed to exacerbate the human resources challenges [67]. To assess IPT completion rates among HIV-patients and determine predictive factors for IPT completion, Adepoju and colleagues (2020) [43] carried out a retrospective cohort study that included HIV-patients from six governmental hospitals in Nigeria. The study reported human resources challenges in many hospitals and service delivery units. Similarly, a Ministry of Health key informant said that staff shortage was a major challenge for implementing IPT in Lesotho [14].

**Knowledge and training gaps** regarding both preventive therapies were identified among health providers, limiting their ability to make appropriate therapy decisions [14, 36, 38, 39, 42, 44–46, 51, 54, 62, 68].

Regarding CPT, knowledge gaps existed among low and mid-level health providers regarding the co-management of patients with HIV and TB [38, 39], CPT for malaria prevention among HIV-positive pregnant women [36, 42], and CPT for babies born to HIV-positive mothers [42]. Kamuhabwa et al. (2016) [36] applied multiple methods to assess CPT implementation among HIV-infected pregnant women in malaria-endemic Tanzania after the national policy had been changed from recommending sulfadoxine-pyrimethamine (SP) intermittent preventive therapy to recommending CPT for malaria prevention in this patient population. Although health providers perceived the new policy on CPT has been clearly written and was understandable, they still did not have sufficient knowledge about the inclusion criteria (HIV serostatus confirmed, time of initiation, no history of allergic reactions caused by sulfur-containing drugs), and exclusion criteria (skin rashes, Stevens-Johnson syndrome). The authors concluded that inadequate training was among the factors driving poor implementation of CPT [36]. Task shifting is an important strategy to strengthen health provider capacity; however, studies from Tanzania and South Africa have shown that lower cadre required training to pursue their roles [36, 42]. Lay counsellors in South Africa admitted they were undertaking tasks outside their scope when providing CTZ to babies [42], while nurses in Tanzania dispensed medicines, including CTZ, without pharmacy training [36].

Regarding IPT, knowledge gaps existed among HIV providers with various levels of education, including doctors. Our evidence synthesis suggested that providers were comfortable managing PLHIV [46], but lacked knowledge about TB [44, 46] and TB prevention [14, 45, 46, 51, 54, 62, 68]. Interviews carried out with doctors specialized in HIV in Brazil reported knowing the ART recommendations, but lacking knowledge about TB prevention and co-management of (HIV and) TB [46]. One of the ten patients interviewed as part of a multi-method assessment carried out in South Africa described being incorrectly denied an IPT prescription; according to the interviewee, the nurse refused to prescribe IPT because the patient did not have TB, suggesting knowledge gaps among nurses in the study setting [51]. Other knowledge gaps identified were related to the target groups for IPT [14, 62], IPT duration [51, 68] and the assessment of IPT eligibility [14, 45, 46, 51, 62]. Two studies revealed a lack of familiarity with IPT prescription requirements for children [14, 62] and pregnant women [62]. One study found that of the twenty-two providers involved in IPT prescription, not many knew that specific tablets were available for children (paediatric dosage) [62]. Lacking clarity was identified about the role of chest x-ray when determining IPT eligibility [45] and regarding the initiation of IPT without placing a TST [51]. Nurses pointed out that although the national guidelines instruct them to prescribe IPT even if the TST is out of stock, the guidelines state that a TST must be placed one month after IPT initiation. Not knowing what to do if TST remained out of stock after IPT initiation, sometimes prevented them from prescribing altogether [51]. Additionally, nurses were unsure how IPT duration varied by patient age, TST result, pregnancy and ART status [51] and about the period after which IPT should be repeated [14]. Research from South Africa revealed that doctors lacked knowledge about the IPT guideline and were concerned about the need to screen for and rule out active TB before initiating IPT [54]. Some health providers had theoretical knowledge about the IPT recommendations [45, 51], but lacked experience or familiarity in applying these guidelines in a clinical setting [45, 51, 54]. One nurse reported she had never used a TST on a patient in her clinic and was unsure whether she could read the result [51]. Lack of knowledge and familiarity with IPT among providers was explained by inadequate staff education [14, 38, 45, 62, 68], and minimal follow-up supervision after training [38]. Criticism was raised regarding the training modules provided to health providers because they lacked content on TB-HIV collaborative activities [38], or included limited or no specific training on IPT [68]. Consequently, providers felt inadequately equipped to handle emerging challenges associated with IPT implementation [68]. Research conducted in Lesotho by Mugomeri et al. (2018) [14], suggested that inadequate training has been the main reason behind delayed IPT initiation in the study setting.

**Negative attitudes, concerns and fears** among health providers impeded the delivery of IPT [14, 44, 45, 53, 54, 62, 64, 67, 68]. Our review revealed that health providers often had a negative or sceptical attitude toward IPT, for which we found several explanations [45, 51, 54, 62, 67, 68]. Providers perceived patients’ irregular clinic attendance, non-adherence to IPT and high loss to follow-up as demotivating and discouraging for IPT initiation. Counselling patients about IPT and encouraging them to adhere to preventive therapy was perceived as difficult and time-consuming [67]. During in-depth interviews, nurses working in HIV care in South Africa expressed that patients misunderstood or were unwilling to cooperate with an IPT prescription. In the absence of external oversight (e.g. clinical mentorship), nurses would lose their motivation to prescribe IPT interviews revealed [51]. Other factors that have discouraged providers from prescribing IPT include discomfort with the long duration of IPT [68], the increasing workload [45, 68], managing IPT among other priorities, and the influence of fellow health providers [68]. Two studies reported that negative perceptions or doubts about IPT among some health providers affected the perception and delivery of IPT by their fellow providers [54, 68].

Although evidence strongly recommends concomitant use of IPT with ART, research from Lesotho suggests a general perception that HIV patients who are on ART for a long time have a low-risk of developing TB. For this reason, providers mostly prescribed IPT to patients newly diagnosed with HIV [14]. We also identified contrasting perceptions about the effectiveness of IPT. Research from Lesotho stated that health providers perceived IPT was effectively reducing TB disease among patients with HIV [14]. Meanwhile, a study from South Africa reported health providers were unconvinced about the benefits of IPT [54]. Concerning was that consequently, health providers omitted to prescribe IPT in the South African setting. Qualitative research carried out by Lester and colleagues (2010) [54] aimed to explore patients’ and providers’ attitudes toward IPT. The study found that none of the twenty HIV-patients interviewed had heard of IPT and doctors were either unaware of the efficacy of IPT in preventing TB or believed the evidence was equivocal. Clinic staff and doctors admitted that IPT was not part of routine practice (such as CPT); the authors concluded that in this setting barriers to IPT were predominantly derived from health providers [54].

Health providers’ concerns and fears were related to active TB [14, 44, 64, 67, 68], antibiotic resistance [45, 54, 64, 68] and side effects [14, 53, 68]. In the first place, health providers were concerned about their ability to exclude active TB based on TB symptom screening. Providers who worked in a setting where no TST was available believed that applying the screening algorithm alone was insufficient to rule out TB [67, 68]. Atypical clinical presentations complicated TB screening of patients coinfected with TB-HIV [54], and occasionally, HIV-patients developed TB disease during [44, 64] or after the course of IPT [14, 44, 64]. Drug resistance was another concern of health providers which led to scepticism about IPT [45, 54, 68]. Bacterial resistance to INH was believed to result from providers’ failure to rule out TB [64], periodic and long-term drug stock-outs [45], patients’ non-adherence to IPT [68] and the use of IPT in settings with a high prevalence of multi-drug and extremely drug-resistant TB [54]. First, failure to rule out TB was revealed by Selehelo, Makhado and Madiba (2020) who reported that health providers sometimes prescribed IPT without symptom screening [64]. Their qualitative study applied in-depth interviews with HIV-patients to explore their experiences regarding IPT provision and derive strategies to improve IPT uptake in community health centres in South Africa. Patients interviewed reported that symptom screening for active TB was not done before the provision of IPT and that they developed TB while on IPT, regardless of reporting symptoms to the nurses [64]. Second, health providers working in selected clinics in Kenya reported they were concerned about contributing to drug resistance by exposing patients to IPT when INH stock-outs did not permit therapy completion. If they were not confident all their eligible patients had 9-months IPT supply providers preferred not to initiate IPT [45]. Third, health providers interviewed in urban HIV clinics in Kenya expressed their concern that patients non-adherence would lead to the development of resistant, multi-drug resistant or extensively drug-resistant TB [68]. Fourth, health providers interviewed in South Africa expressed such fear of inducing multi-drug resistance through INH monotherapy that some suggested starting empirical courses of full treatment for TB. Providers believed a combination of drugs was safer and offered greater benefit than INH monotherapy [54]. Eventually, some providers feared that INH could induce side effects [14, 53, 68]. Overall, the fears and concerns described made health providers feel uncomfortable or unwilling to prescribe IPT.

**Provider-patient communication** emerged as a barrier to the implementation of IPT. Our review revealed that patients were often afraid to speak to health providers about IPT, suggesting ineffective provider communication and a lack of trust in the health provider-patient relationship [50, 51, 58]. Jacobson and colleagues (2017) [50] explored patients’ reasons for default from IPT through individual interviews. IPT defaulters stated that they avoided reporting side effects, questioning missing medications, or requesting to re-start IPT after treatment lapses, due to their fear of nurses scolding [50]. Research from Ethiopia found that patients lacked advice from doctors, feared side effects and stigma [58]. The authors concluded lack of provider-patient communication contributed to patients non-adherence to IPT [58]. Similarly, HIV-patients interviewed in South Africa revealed that they felt intimidated during their conversation with nurses; nearly half of them reported that nurses were sometimes rude and that this discouraged them to ask about IPT [51].

#### Clinical information related barriers

**Inaccurate recording and lacking integration of health records emerged as a** barrier for both preventive therapies [14, 36–38, 42, 45, 51, 63, 67]. Although poor recording of clinical information on patients’ records was frequently reported [14, 37, 38, 42, 45, 67], our review revealed that incomplete records did not necessarily imply that services have not been provided [14, 37, 42, 45]. Horwood et al. (2010) interviewed HIV-positive mothers in South Africa, regarding the health services and interventions their babies had received, and compared this information with data recorded on the post-natal ward- and child health records [42]. According to the mothers, significantly more babies had received CPT, compared to what has been recorded on their health records [42]. Similarly, Lougwagie et al. (2012) [37] reviewed altogether over two-thousand patient records from separate and semi-integrated health facilities. Their study showed that CPT was less well recorded than counselling and HIV testing, raising the question whether this was due to poor recording or poor provision of the preventive therapy [37]. Van Ginderdeuren and colleagues (2019) [67] obtained a similar pattern of results for IPT. Their record review showed a discrepancy between what has been recorded by health providers in IPT registers and what patients had received according to pharmacy records. The authors concluded underreporting of IPT in routine IPT registers [67]. In line with this finding, Mugomeri et al. (2018) [14] suggested underreporting of monthly statistics through the paper-based record system.

However, we identified weaknesses related to the health information systems available to record patient data, contributing to inaccurate recording practices. First, health information systems in many TB/ HIV high burden countries were paper-based or relied on a mix of paper-based and electronic records [14, 45]. Research from Uganda described recording tools were poorly designed and unpractical; HIV registers lacked data entry sections regarding TB-HIV collaborative services, and TB information were only recorded on patients’ chronic care cards [38]. Similarly, Jarrett et al. (2019) [51] reported that patient charts were disorganised and lacked a specific log for IPT registration, making it challenging to track each patient’s progress toward IPT completion. Most nurses discontinued IPT to avoid the possibility of prescribing IPT past the intended duration, a study from South Africa revealed [51]. Providers relied on a patchwork of documents completed by various team members in a Kenyan study setting where patients did not see the same provider during each visit [45]. The study reported that health providers frequently suffered from missing patient data. In case patient data were missing, patient self-report was used to determine the steps of TB diagnosis and IPT eligibility assessment previously completed. Doctors reported they trusted patient recall over electronic medical record (EMR) data when patient information conflicted [45].

Our review revealed that lacking integration of patient data recorded from different service units bears two risks. First, documentation of patient data in separate patient files (e.g. HIV registers, patients’ chronic care card, laboratory-, pharmacy records) affected service delivery efficiency by multiplying the time spent for document retrieval, data entry, and in some cases the provision of services that had already been provided [38, 45]. Second, in the context of TB-HIV co-infection, poor recording, communication and lacking service integration [36, 63] may lead to preventable drug interactions or adverse reactions. A study from Tanzania reported that eight HIV-positive pregnant women were co-administered both SP and CPT for malaria prevention, which is contraindicated because of increased risk of severe cutaneous reactions [36]. Research from South Africa reported that four patients whose clinical records suggested they were not eligible for IPT, had started the course [63].

**Ineffective monitoring, evaluation and surveillance** emerged as a second barrier related to clinical information [14, 38, 56, 67]. Both concepts, monitoring and evaluation (M&E) and surveillance require collecting data, that are analysed, and finally transformed into strategic information that governments and leaders use to make informed decisions. We identified weaknesses of the health information system and found that patient data recorded at the health facility level were commonly inaccurate or lost [14, 45]. The detection and notification of health events (surveillance) and reporting of routine programme data (M&E) consequently leaked in its foundation.

In 2009, research from Uganda reported that no tools existed for routine recording and reporting of TB-HIV surveillance data that permitted informing planning processes [38]. Eight years later, McRobbie et al. (2017) [56] reported that there were still no regional or national information systems in place, that allowed the integration and submission of health facility data in Uganda. Different views about obstacles for M&E in Lesotho were particularly well described by Mugomeri et al. (2018) [14]. Through interviews with key informants, the qualitative study found that major barriers to the implementation of IPT included ineffective health information systems [14]. The representative of the WHO country office in Lesotho warned that the IPT indicator system was paper-based and that data reported by health facilities were inaccurate. One health provider acknowledged that the total number of patients enrolled in IPT was unknown due to a shortage of paper registers [14]. The provider added that monthly reports were sent to the MoH for congregate analysis, but feedback was rarely received at the health facility level. The investigators confronted the responsible ministry of health (MoH) representative who explained that it was not feasible to frequently assess the effectiveness of their programmes and that their primary goal was to increase IPT uptake. Providers were concerned that INH could induce side effects; some believed that poor M&E resulted in the late detection of these undesirable reactions [14]. A partner organization representative acknowledged the need for more resources, including stationary and patient registers, and noted that failure to monitor patient referral, additionally confounded monthly statistics [14]. A study from South Africa showed that IPT indicator data were not comparable due to inconsistent denominators across health facilities. Some facilities routinely collected data to measure IPT uptake among all PLHIV and others used HIV-patients newly enrolled in HIV care as denominator [67]. Weaknesses identified by Okot-Chono et al. (2009) [38] included that the existing supervisory visits carried out by national and district officers for monitoring of the TB-HIV collaborative activities were infrequent and had not been performed jointly by the TB and HIV programmes.

#### Pharmaceutical management related barriers

**Drug stock-outs or shortages** were common for both preventive therapies [14, 15, 33, 34, 36, 38, 45, 50, 51, 56–58, 62, 64, 65, 67, 68]. Except from one study that reported stock-outs in India [62], all other studies reported stock issues in countries located in sub-Saharan Africa. African countries included Kenya [45, 68], South Africa [50, 51, 64, 65, 67], Uganda [38, 56], Tanzania [15, 36], Zimbabwe [34], Nigeria [57], and Lesotho [14].

Inconsistent drug supply, periodic and long-term stock-outs, or shortages of CTZ [14, 15, 33, 34, 36, 38] and INH [14, 45, 50, 51, 56, 57, 62, 64, 65, 67, 68] were obtained from record reviews, reported by patients, health providers, representatives of MoH or partner organizations. It may be noteworthy, that stock-outs were also reported for dapsone, an alternative drug to CTZ [14] and pyridoxine, a drug prescribed to prevent peripheral neuropathy caused by INH [14, 62]. Our review revealed consequences of and reasons for the inconsistent availability of CTZ and INH.

Stock-outs of INH hindered therapy initiation [45, 56, 62], therapy adherence [64, 65] and completion [62]. Interviews with doctors in Kenya showed that providers remained hesitant to initiate IPT due to frequent pharmacy shortages [45]. Providers in Kenya perceived stock-outs of INH and other supplies related to IPT as a major impediment to effective implementation and acceptance of the therapy. Some providers felt the poor supply of INH showed a lack of support for the IPT programme among policy-makers and management. This, in turn, negatively affected their perception, morale and delivery of IPT [68]. HIV patients interviewed in South Africa, reported medication stock-outs as the reason for defaulting or not adhering to IPT [50, 65]. In-depth interviews with HIV-patients in South Africa revealed that patients perceived the occasional unavailability of INH as inconvenient and frustrating because it increased clinic visits and may lead to non-adherence [64]. One study suggested that the location from which PLHIV collected their drug supply may have predicted adherence to IPT, as peripheral pharmacies ran out of INH more frequently than hospital pharmacies [65].

For CTZ, the included evidence suggested that drug shortages led to irregular uptake of CPT [38], dose alterations among infants [15, 34], and patients skipping scheduled clinic visits [36]. Stock-outs of CTZ at health facilities and HIV clinics were reported as the reason why many children had missed one or more doses of CPT [14] and why only a few TB/HIV co-infected patients received CPT regularly [38]. Studies from Tanzania and Zimbabwe found that frequent shortages of the paediatric formulation at health facilities caused challenges to caregivers and providers [15, 34]. In Tanzania, parents had difficulties understanding how to crush tablets and prepare suspensions for their infants [15]. Lack of CTZ at the health facilities was suggested as a reason for pregnant women skipping visits scheduled at the antenatal clinics [36].

Among explanations to why health facilities and pharmacies frequently ran out of CTZ, we found an increasing number of HIV patients [38], inaccurate recording [36], challenges with quantification and forecasting of the prescribed preventive therapy [36, 38]. At the health facility level, we identified the late submission of health facility requisitions [38] or ordering of drug quantities that did not cover the facility demand [36]. Outside the health facility level, we found district-wide stock-outs of INH [51] and scarcity of CTZ at the medical stores department or government supplier side [15]. A lack of support from policymakers, programme management [14] or district health departments to adequately supply the lower level units with medicines was also believed to facilitate stock-outs [38, 68]. A district health officer interviewed in Uganda reported that it was not surprising that CTZ was not available at the health facilities. Health offices failed to support lower-level health providers with correctly quantifying drugs in stock and calculate drug consumption [38]. Ineffective supply chain management [13, 35, 63, 65] and logistical challenges [58] provided alternative explanations for frequent stock-outs. Finally, an uncoordinated implementation of IPT shown by Meribe et al. (2020) [57] also led to long-term INH stock-outs. Their study reported a strong focus on IPT at the beginning of the implementation process in Nigeria, which led to the utilization of all stock at most sites, followed by national level stock-outs that lasted from December 2018 to April 2019 [57].

**Lack of pharmaceutical personnel** emerged for both preventive therapies [36, 56]. Studies reported that sites in Uganda lacked a dedicated logistics manager [56] and that health facilities lacked pharmacy personnel in Tanzania, where nurses without pharmaceutical training dispensed medicines [36]. **Lack of written instructions** was reported by Kamuhabwa et al. [36] referring to missing documented strategies for ensuring the availability of CTZ at the health facilities on one hand, and missing written instructions clarifying how medicines should be administered by patients at home on the other hand. The authors suggested the need to provide patients with a list of known potential adverse reactions, including those that require to stop taking the therapy [36].

#### Service delivery related barriers

**Sub-optimal service delivery** emerged as a critical barrier for the implementation of both preventive therapies. We identified examples of sub-optimal service delivery in fourteen studies that reported from a large range of countries with a high burden of TB and HIV [15, 34, 37, 38, 42–44, 46, 50, 51, 56, 62, 64, 68]. These examples primarily showed that service delivery was time-consuming, the patient was not put at the centre of care and that health services delivery was inefficient.

Service lapses, limited working days, and doctors’ limited working hours have been reported at public health facilities [44, 50]. A study from Uganda found that the AIDS information Centre was closed when patients returned the first time after testing HIV-positive; they never returned thereafter [44]. Studies reported that patients faced long queues, spending hours waiting before receiving services or their medications [34, 38, 50, 64]. Jacobson et al. (2017) [50] reported that HIV patients endured long queues to collect their medicines, often skipping meals and taking medications on an empty stomach [50]. Dispense of IPT and ART in separate lines [50] and the lacking synchronization of IPT with other HIV clinic appointments [43] added to patients’ waiting time and clinic visits. A qualitative study explored women’s ability to maintain their babies on CPT. The study found that mothers typically had to dedicate the entire day to one clinic visit, which they found difficult, considering they also had to attend the clinic for their own HIV care [34]. Due to lack of same-day services, one family had to spend multiple days at the clinic for HIV care [34, 38], providing a possible explanation why only a few HIV-positive mothers in South Africa reported attending regular follow-up for their own care [34], and why patients felt discouraged. Overall, services delivery did not appear patient-friendly and routinely tested patients’ willingness and ability to receive HIV care [44]. Other examples of sub-optimal service delivery showed weaknesses in the quality of healthcare provided.

Patient information, including tolerance to preventive therapy, side effects, drug interactions, and therapy adherence, were frequently missing on patient records [14, 64, 66]. Some HIV-patients reported that their test results got lost at the health facility [50]. Interviews revealed that HIV-positive mothers had received insufficient quantities of CTZ, lasting for forty days, while their next clinic appointment was two months away [34]. Often, there was no follow-up of HIV-patients who missed clinic appointments [15].

Among the potential underlying causes of sub-optimal service delivery, the disproportional provider-patient ratio was the most prominent. Human resource constraints presented a critical operational challenge to HIV services delivery [56, 62, 64]. As a result of MoH funding constraints (and differing donor priorities), there were too few providers available to provide necessary services [56]. It may be noteworthy that since the scale-up of HIV testing and ART followed by the universal treatment policy released in 2015, there has been an ever-increasing number of PLHIV requiring continuous treatment and care. So far, the majority of HIV services have been delivered at the health facility level, resulting in organisational and operational challenges to handle the heavy patient load [15, 38, 40, 56, 62, 64]. For IPT, the high patient load was believed to prevent nurses from screening every patient for IPT eligibility [51]. Among other factors, IPT service delivery was believed to be greatly affected by lack of integration between HIV and TB services [46], the inconsistent INH drug supply [51, 62, 68] and inadequate records for IPT registration [51].

**Inadequate facility infrastructure** emerged as barrier theme for both preventive therapies [15, 38, 56]. Health facilities in Tanzania lacked weighing scales to determine infants’ weight for CTZ doses calculation. Thus, providers based the doses on age rather than weight, unable to ensure infants received the correct amount of preventive therapy [15]. Facility infrastructure has been reported as inadequate to ensure patients’ privacy and confidentiality; lacking counselling rooms and inadequate space to implement the HIV policies was reported [38, 56]. Research from Uganda suggested that an outside waiting area for HIV services attendees may raise suspicion among husbands seeing their wives there seated, suspecting that she may have contracted HIV [38].

**Poor integration of preventive therapy-related services** was another important cross-cutting barrier theme [14, 33, 36, 38, 42, 46, 50, 54, 56, 68] that arose from the segregated delivery of health services in many high TB/HIV burden countries. Studies showed that preventive therapies were frequently not part of the services provided at the health facility [33, 42, 56]. Not all primary care facilities in Malawi and Uganda offered CPT for patients with HIV on-site [33]. Only half of the immunization clinics in a South African study setting provided CPT for HIV-exposed babies [42]. None of the twenty-four health facilities, included in a study carried out in Uganda, offered IPT as service for patients with HIV [56]. Likewise, research showed that additional testing services required before initiating IPT were also frequently not available on-site [54, 68], which meant that for receiving preventive therapies, patients had to attend other health facilities as well.

Overall, preventive therapy was poorly integrated with other speciality services [36, 38, 46, 50, 54, 68], often requiring patient referral between health facilities and incurring additional patient costs [38, 68]. However, inter-clinic referral was reported to be weak [14, 38], added to patients’ costs [68] and challenged service quality [54]. In Kenya’s capital HIV clinics, most clinical examinations required before IPT initiation were carried out in separate departments [68]. According to a study that was carried out in South Africa, only one of ten clinics had onsite chest radiography, while none could test sputum on-site. Investigations for TB performed off-site lacked coordination between TB and HIV shown in lost specimen and laboratories not communicating TB test results with HIV sites. Lacking coordination between TB and HIV sites was believed to hinder patients’ initiation of IPT [54]. Poor inter-clinic referral and separate clinic days for TB and HIV services were reported in Uganda, demonstrating poor integration of IPT with HIV care [38]. Interviews with key informants in Lesotho suggested that confirmation of patients’ arrival at the referred centre was often lacking [14]. Wambiya and colleagues (2018) [68] interviewed eighteen health providers in urban Kenya to explore factors that influenced providers’ acceptability of IPT. The study reported that most clinical examinations were conducted in separate departments at additional costs. Health providers urged that all testing services should be provided at the same site. Costs incurred by additional exams should be subsidised to increase IPT uptake, providers argued [68]. The need to refer patients to another facility for preventive therapy, not only added to patients’ waiting time and transport costs, it has shown to carry service delivery related risks, such as dual administration of intermittent treatment for malaria prevention in HIV-positive pregnant women. A study identified patients in Tanzania who received both, SP at the antenatal care unit, and CPT at the Prevention of Mother To Child Transmission (PMTCT) unit [36]. Research from Uganda reported that few TB patients were tested for HIV, not identifying co-infected patients eligible for CPT [38]. In South Africa, efforts have been made to integrate IPT-related services with ART services at the same location. However, interviews with HIV-patients from four primary care health facilities and one district hospital revealed that IPT and ART were dispensed in different queues and that most facilities had not fully integrated TB and HIV services on-site [50].

**Patient loss to follow-up** was identified for both preventive therapies [42, 48, 55, 56, 62, 63, 67], referring to patients not linked to care (after HIV diagnosis) and those who stopped attending clinic appointments. Mc Robie and colleagues (2017) [56] argued that recent efforts in Uganda had focused on HIV testing and immediate ART provision without strengthening health facility infrastructure and human resources accordingly to allow the implementation of the complete set of HIV care policies. Consequently, systems failed to maintain patients in care downstream in the care cascade, the authors concluded [56]. Besides difficulties to retain patients in care, engaging those reluctant to present for HIV services (i.e. testing, care and support) from the beginning, also remained a challenge for HIV service delivery [63].

A sub-study of the Botswana IPT trial explored self-reported reasons for trial participants’ loss to follow-up (LTFU) following IPT initiation. Reasons included patients’ financial and organisational feasibility to attend the clinic appointments, lacking motivation or social support. Stigma, relocation, religious beliefs, and pregnancy also comprised reasons for patients to stop preventive therapy, while some believed they had completed the study [48]. We identified another example showing patients’ loss to follow-up due to lack of linkage to care, which can turn into a lost opportunity to provide CPT, as shown in a study from South Africa. Coverage of PMTCT interventions was high during pregnancy and delivery, but mothers and HIV-exposed infants were poorly linked to follow-up care after the baby was born. The provision of CPT was also poorly recorded on the child health records. Follow-up care (e.g. CPT and HIV testing of the new-born) therefore depended on the mother reminding the health provider that their baby has been HIV-exposed. Less than half of the eligible babies had a PCR test and initiated CPT at their first visit to the immunization clinic [42]. Research suggests that patients’ non-completion of IPT was frequently associated with failure to return to the clinic for follow-up of IPT [55, 62]. Recent efforts have focused on assigning health providers responsible for contacting these patients via telephone or in-person. However, providers perceived that following-up of these patients was difficult and time-consuming [67].

**Ruling out TB disease** has been reported as a major barrier to implementing [14, 38, 45–47, 49–54, 62, 64, 67, 68]. Due to the need to rule out TB disease before therapy initiation and during the course of IPT, delivery of IPT has been perceived as difficult [53]. Since 2011, WHO recommends using a simple algorithm to identify PLHIV unlikely to have active TB, no longer requiring other tests in countries where other tests are not available [13]. However, some countries continued using other recommended tools to determine patients’ eligibility for IPT. According to a recent email survey [47], three different TB screening tools were used for PLHIV in high TB/HIV burden countries. Besides symptom-based TB screening (Kenya, Lesotho, Liberia, Myanmar, Nigeria, South Africa, Tanzania), tuberculin skin tests (Brazil, Ethiopia, Indonesia and Thailand) and interferon-gamma release assays (Ethiopia) have been used. Several countries reported combining screening tools (Brazil, Ethiopia, Indonesia, Kenya and Thailand). For example, Brazil reported using a combination of tuberculin skin tests (TST) and chest radiography, while Kenya reported a combination of symptom-based screening and radiography [47]. Evidence synthesis revealed several limitations for each screening tool.

First, weaknesses of TST were frequently associated with the tuberculin (purified protein derivate) required to carry out the test. We identified shortage of purified protein derivate (PPD), delayed or insufficient supply from the manufacturers [47], challenges of ensuring the quality of PPD [47, 49], as well as the limited utility of TST in settings where BCG vaccination continued [47]. One study aimed to assess the acceptability and operational feasibility of TST in urban Kenya and rural Swaziland [49]. The study revealed that some health providers initially confused tuberculin with BCG vaccine. In both settings, some patients received an inappropriate amount (half-dose) of tuberculin injection. In rural Swaziland, cold chain monitoring issues were experienced [49]. Besides, the need for a second visit for TST reading was a major concern among patients and providers, implying additional time and transport costs [49, 67]. A questionnaire survey designed to identify barriers to IPT was administered to health providers in South Africa [67]. Survey findings showed that health providers had no time to perform TST; they found the TST reading time consuming and struggled to motivate patients to return for the reading [67]. Research from Brazil reported an alarmingly long time between the first clinic visit to IPT initiation. The time delay was explained by the fact that TST was not routinely offered in all clinics on all days and the chain of processes involved in TST. Once an asymptomatic patient received a positive TST result, chest radiography was required to rule out TB, for which the patient received a prescription. The patient then needed to schedule an imaging appointment before returning to the clinic to present the result to the doctor. Adding to the delays involved in the process of TST testing, patients may not have felt the urgency to quickly return to the clinic because they did not feel sick, the authors argued [46]. A review of patient records from five primary care health facilities revealed that only the facility that opted against TST reported a significant increase in IPT uptake [67]. Overall, the reported findings suggest that TST may have contributed to the low uptake of IPT [46, 47, 67]. At the time of our review, TST had been removed from most high burden countries’ IPT policies. However, some countries, including Brazil, continued to apply TST as a necessary screening component [47]. Proponents of TST argued that providing IPT only for those with latent TB infection (after careful exclusion of active TB) would avoid unnecessarily treating a significant number of patients that do not stand to benefit [49].

Second, more recently developed interferon-gamma release assays (IGRAs) address important limitations of TST. Patients are only required to visit the clinic once for blood collection and the result is available within one day. Additionally, prior BCG vaccination does not cause a false-positive IGRA test result. However, due to a lack of budget, lab infrastructure and trained personnel, and insufficient capacity for specimen transport, these relatively expensive tests have hardly been used in TB/HIV high burden countries [47].

Third, symptom-based TB screening refers to a simple algorithm applied prior to initiating IPT and at every follow-up visit, thus allowing health providers to interrupt preventive therapy in case a patient develops symptoms of TB. Those who do not report any symptoms of current cough, fever, weight loss, or night sweats are unlikely to have active TB. If no contraindications to IPT exist, these patients should be o□ered preventive therapy [13]. Although symptom-based screening has significantly advanced IPT uptake in low-resource settings [53], we identified challenges associated with this approach.

Challenges included doubts about the reliability of TB symptom screening among patients with HIV and TB-HIV, lacking provider capacity to routinely screen TB symptoms, and service delivery inefficiencies. Solely based on TB symptom screening, health providers found it difficult to rule out active TB [53] or decide whether a patient had active or latent TB [68]. Some providers believed that using the screening algorithm alone was insufficient to rule out TB [54, 67, 68]. Opponents of symptom-based screening argued that atypical clinical presentations complicated TB screening of patients coinfected with TB-HIV who frequently suffered from extrapulmonary TB [54]. Khan et al. (2014) carried out a diagnostic performance evaluation among HIV patients in South Africa [52]. Study findings suggested that symptom-based screening was very sensitive and diagnostically useful among patients not on ART, however, less reliable among patients on ART [52]. The high patient load was believed to prevent providers from screening every HIV-patient for IPT eligibility [38, 51]. During interviews conducted in South Africa, nurses repeatedly stated that staff shortages and the large patient burden prevented them from screening every HIV patient [51]. Research revealed possible consequences of providers’ lacking capacity to screen patients for TB. Health providers either lose their motivation to prescribe IPT [51] or continue prescribing IPT without TB symptom screening [64, 67]. The latter was shown by Selehelo et al. (2019) [64]. The authors reported providers were prescribing IPT without screening for TB symptoms, which meant risking a delay of TB treatment and favouring the emergence of drug resistance [64]. We identified service delivery issues with symptom-based screening that delayed IPT initiation. Research from Lesotho [14] suggested that ineffective TB screening was due to an inefficient service delivery system. Delays in IPT initiation have been reported for patients with presumptive TB, indicating it took much time until TB was ruled out. The study also found that IPT was mainly initiated among patients newly diagnosed with HIV, while IPT uptake among patients enrolled on ART before IPT was launched in 2011 lagged behind [14]. Health providers interviewed in India reported that proxy attendance delayed IPT initiation. In the mainly rural study setting, patients often send their caregivers or other attenders to collect their ART refill, making it impossible for providers to assess patients’ eligibility for IPT [62]. In Kenya, a combination of TB symptom screening and obligatory chest radiography was applied to determine HIV-patients’ IPT eligibility [45, 47]. Doctors reported they were unsure about the steps to take to determine IPT eligibility. However, doubts were mostly related to the role of chest x-rays, when and if it should be ordered and how to read the radiographs [45]. Irrespective of the screening tool applied in each country, suspects of TB required additional investigation.

**Investigation of TB suspects** is a crucial requirement for all patients with a positive TB screening result [13]. For those who developed TB during the course of IPT, additionally, drug susceptibility testing should be performed [52]. However, we identified several challenges associated with evaluating patients with presumptive and confirmed TB [14, 38, 45, 49, 52, 54, 64, 67]. Diagnostic equipment needed to investigate TB included chest radiography [54], laboratory equipped for the analysis of sputum samples [45] and the identification of resistant TB strains [52]. However, the capacity to diagnose TB was limited in high TB/HIV burden countries, where most health facilities could not diagnose TB on-site [52, 54]. Although the more accurate GeneXpert® test allows rapid diagnosis and identification of patients with drug-resistant TB, this expensive molecular test technology was often only available at centralised sites, reserved for a subset of complex cases [14, 45]. Besides, our review revealed knowledge gaps among health providers related to co-management of TB-HIV [38] and their diagnostic competences [38, 45]. In Kenya, doctors specialised in HIV reported they were unsure how to read a chest radiograph [45]. Providers in Uganda reported struggling with the assessment of smear-negative and extra-pulmonary TB [38]. Most concerning was the study finding that only a minority of PLHIV with self-reported symptoms were investigated for TB [49, 67], causing TB treatment delays [64] or resulting in too many patients denied for IPT [49].

#### Health system financing related barriers

**Health system funding issues** were identified for both preventive therapies [14, 15, 47, 56]. Many high TB/HIV burden countries historically depended on funding for HIV and TB from international donors and governments. Many will continue to require donor backing to improve the provision of a comprehensive care package to PLHIV [56].

Lacking funds to purchase essential medicines (as which CTZ and INH are classified), has been reported in a Tanzanian study setting [15]. The quantitative descriptive study involved questionnaire-based interviews with health providers, who suggested lack of funds to purchase essential medicines at the health facility level had been an attributing factor for drug scarcity at public hospitals [15]. Similarly, qualitative research from Lesotho reported that among the main challenges for implementing IPT were interrupted supply chains caused by insufficient health system funding [14]. Through interviews with MoH key informants, health providers and representatives of partner organisations, the study found that additional resources were needed to sustain a seamless supply of drugs and reagents. A health provider reported that health facilities frequently ran out of CTZ, pyridoxine, and dapsone, among other drugs. Ministry of health officials expressed the need for health system funding to be increased to improve the sustainability of HIV/ TB programmes, including IPT [14]. A recent email survey carried out by Faust et al. (2020) [47] aimed to identify challenges experienced in countries with a high burden of TB regarding the implementation of latent TB infection policies. Respondents from six high TB/HIV burden countries (i.e. Ethiopia, Liberia, Nigeria, South Africa, Tanzania, Zimbabwe) reported that guidelines existed, but due to financial barriers guidelines were not fully implemented [47].

**Vertical funding** emerged as barrier theme for both preventive therapies [14, 38, 56], referring to a stand-alone programme approach (e.g. HIV programme, TB programme) used in many resource-constrained countries for health system funding, typically associated with vertical governance and health service delivery. However, health system strengthening and the integrated provision of interventions that go beyond a single disease programme are not characteristic for vertical funding. Both preventive therapies are good examples for interventions that require some degree of collaboration between disease programmes (i.e. HIV, TB, PMTCT, child health) to ensure their successful implementation. Lacking funds for implementing TB/HIV collaborative activities in Uganda was reported on a national TB/HIV programme level and district level [38, 56]. Policymakers reported international donor dependency of the HIV programme [56]. Implementers and researchers interviewed about HIV policy implementation in Uganda reported misalignment of goals between the government and donors in determining priorities, suggesting lacking donor support for health systems strengthening and prevention activities [56]. Donors were considered to focus more on the provision of ART [56], while human resources constraints presented a critical operational challenge for providing HIV services, including IPT [14, 56]. However, funding of human resources primarily rested with the individual Ministry of Health [14, 56].

#### Leadership & governance related barriers

**Issues regarding policies and guidelines** emerged for both preventive therapies [14, 15, 38, 47, 51, 54, 67, 68]; however, were most commonly reported for IPT. A recent survey [47] suggested that guidelines on latent TB infection (LTBI) did not exist in several high TB/HIV high burden countries (Angola, China, DRC, India, Indonesia, Kenya, Myanmar) [47]. Barriers to guideline implementation reported among these countries included prioritization of active TB over LTBI management (Angola, China, DRC, India, Kenya), financial barriers to programme implementation (Angola, DRC, Kenya), lack of programme staff (DRC) as well as guideline development still in progress (India, Indonesia, Kenya, Myanmar). The study suggested that in China, PLHIV were not targeted for LTBI treatment. However, the evidence relied not only on National TB programme staff but also on researchers and health officials. Findings, therefore, reflect the respondents’ current knowledge and not necessarily the official situation in their respective countries [47].

Preventive therapy guidelines were not always accessible [15, 38] and understandable for health providers [14, 15, 38, 51, 68]. Health providers in Tanzania reported no up-to-date guidelines were available at the health facility, hindering effective implementation of CPT for children born to HIV-positive mothers [15]. In Uganda, district officers had received policy guidelines on TB/HIV collaborative services but had not disseminated the documents to providers and communities [38]. In Kenya, South Africa and Lesotho, national guidelines for IPT were described as unclear [14, 68], ambiguous, confusing, and incomplete [51]. Lesotho’s guideline was reported to lack clarity about the duration after which IPT should be repeated. Health providers noted that they were left with too much flexibility regarding the timing of IPT initiation. Some providers initiated ART before IPT to allow unmasking side effects of ART and ensure patients are stable. Others prescribed IPT only two weeks after ART initiation. This flexibility in the guidelines was believed to have contributed to the poor implementation of IPT [14]. Research from Kenya suggested that lacking clarity of IPT guidelines meant that providers struggled to implement IPT effectively. According to health providers from three HIV clinics in Nairobi, Kenya, particularly unclear were eligibility criteria and the duration of IPT, as well as how to decide whether a patient had active or latent TB [68]. Lacking clarity might have contributed to health providers’ low fidelity to the IPT guideline in South Africa [54, 67]. Van Ginderdeuren et al. (2019) [67] reported that across selected primary care health facilities, TST was not implemented as intended, symptomatic patients were not investigated for TB, and IPT initiation rate was generally low. Others have shown that, only in one out of ten sites included in their study, health providers prescribed according to the existing policy guideline. Providers seemed to lack clarity regarding the eligibility criteria for IPT. For example, some thought that IPT was indicated for patients with a CD4 count below 200 cells per microlitre, or reserved for patients on ART [54].

**Lacking leadership and coordination** emerged for both preventive therapies [14, 38, 42, 46, 50, 51, 68]. Both interventions require some degree of collaboration between disease programmes (i.e. HIV, TB, PMTCT, maternal and child health) to ensure their successful implementation. However, weak leadership and coordination at the strategic and operational level have been reported as a major challenge for TB/HIV collaborative activities [38, 46, 68]. Studies showed that TB and HIV services’ responsibilities remained fragmented [46, 50] with lacking clarity concerning roles and responsibilities for collaborative service delivery at the health facility level [38, 42]. Research from Uganda found that it was not clear in the Ugandan study setting whether the responsibility for coordination belonged to TB or HIV health providers. With the exception of two health facilities, no coordination meetings had been held between the two programmes [38]. Mugomeri et al. (2018) [14] suggested that lack of leadership was among the major barriers to scaling-up IPT for PLHIV in Lesotho. Research carried out in two KwaZulu-Natal districts showed that PMTCT services were poorly integrated with maternal and child health services. Interviews with nurses and lay counsellors revealed a lack of clarity regarding their roles and responsibilities in the provision of CPT for HIV exposed babies [42].

**Lacking top-down policy support and management issues** emerged as barriers for both preventive therapies [38, 50, 51, 68]. On an institutional and policy level, we identified a lack of commitment, support and oversight as barriers to implementing IPT policies and collaborative TB/HIV activities. Providers interviewed in Kenya felt demotivated by the limited commitment at the policy level in ensuring effective implementation and streamlining of the IPT programme [68]. After investigating patients’ reasons for defaulting IPT, Jacobson et al. (2017) [50] concluded that long queues, drug stock-outs, and a lack of service integration require attention on an institutional and policy level. At the health facility level, ‘no-one was overseeing’ the implementation of TB/HIV collaborative services, participants of a study conducted in Uganda reported. Inadequate joint supervision may lead to providers’ poor performance, the authors suggested [38]. Similarly, in South Africa, nurses admitted that in the absence of external oversight, they would lose their motivation to prescribe IPT [51]. The multimethod study carried out by Jarett et al. (2019) [51] involved interviews with health providers after they had received a nurse-centred intervention. The intervention was designed to promote two target behaviours: checking IPT-eligibility for all PLHIV and prescribing IPT to this target population. The authors concluded that without adequate oversight, complicated guidelines would continue to hinder the implementation of IPT.

**Inadequate planning** was identified as a barrier to the implementation of IPT and TB-HIV collaborative activities [14, 38]. Inadequate national planning was reported as an important barrier to IPT implementation in Lesotho, key informants said. Representatives of partner organizations reported that IPT was a□ected by lack of foresight at the planning stage and poor capacity to solve problems that arise [14]. Multimethod research from Uganda revealed that out of five districts implementing TB-HIV collaborative activities, only one had incorporated collaborative activities into its district work plan. The remaining four districts had separate HIV and TB plans that excluded TB-HIV joint activities. Another barrier to TB-HIV collaborative activities reported by the authors was that patients and communities were not involved in the planning process [38].

**Lacking stakeholder engagement** emerged as a barrier theme for both preventive therapies, suggesting that community consultation and engagement of implementers and the population targeted for preventive therapies were insufficient, which may have hindered the scale-up of preventive therapies [14, 38, 68]. According to research from Uganda, community-related activities were planned without community involvement. Additionally, the study reported a lack of information flow from health facilities to the communities concerning the TB and TB-HIV services available [38]. The limited engagement of health providers in the development of the IPT guideline provided one explanation for their low acceptability of IPT in Kenya [68]. Health providers interviewed in Nairobi’s selected HIV clinics reported they were told: “Here are the guidelines to be followed!”. Providers felt they should have been involved in the process of guideline development [68]. Similarly, lacking engagement of the people responsible for implementing IPT (i.e. health providers) was reported to inhibit IPT uptake in Lesotho [14].

#### Metasummary

We summarised the identified barrier themes in tables and compared both preventive therapies with respect to similarities and differences of barriers (S8 Additional file). Overall, thirty-two barrier themes emerged from the comprehensive barrier description of which the majority were cross-cutting barrier themes (n = 25). We also identified seven intervention-specific themes.

Barrier themes specific to CPT (n = 2) were ‘concerns about the efficacy of CPT’ in areas of high bacterial resistance [31, 35] and a ‘lack of written instructions’ (i.e. procedures to ensure the availability of CTZ and patient information) [36]. Barrier themes specifically identified for IPT (n = 5) include ‘patients’ socio-demographic, lifestyle and clinical factors’ [14, 46, 48, 55, 59–62, 66, 68], ‘providers’ attitudes, beliefs & fears to induce INH resistance’ [14, 44, 45, 51, 53, 54, 62, 64, 67, 68] and challenges related to ‘provider-patient communication’ [50, 51, 58]. Additionally, we identified issues with ‘ruling out TB disease’ [14, 38, 45–47, 49–54, 62, 64, 67, 68] and ‘the investigation of TB suspects’ as intervention-specific barriers for IPT.

After carefully grouping each barrier to the health system component from which it arose, we compared both preventive therapies with respect to similarities and differences of health system components most frequently associated with barriers. We found that barriers to both preventive treatments were most frequently related to ‘health service delivery’ and ‘patients & their community’. For IPT, health provider related barriers seemed to play an additional important role, third most frequently referred to. In contrast, barriers to CPT, were third most frequently related to ‘pharmaceutical management’. ‘Financing’ was among the health system components least frequently associated with barriers to either of the preventive therapies. We applied an innovative mapping technique to summarise the comprehensive barrier information. The resulting metasummary graphics illustrate all identified weaknesses (i.e. barrier themes) within each of the seven health system components, together with the number of papers that contributed information to each barrier theme. Therefore, mapping allows visual comparison of barrier themes identified for CPT (Fig 3) and IPT (Fig 4).

**Fig 3.**
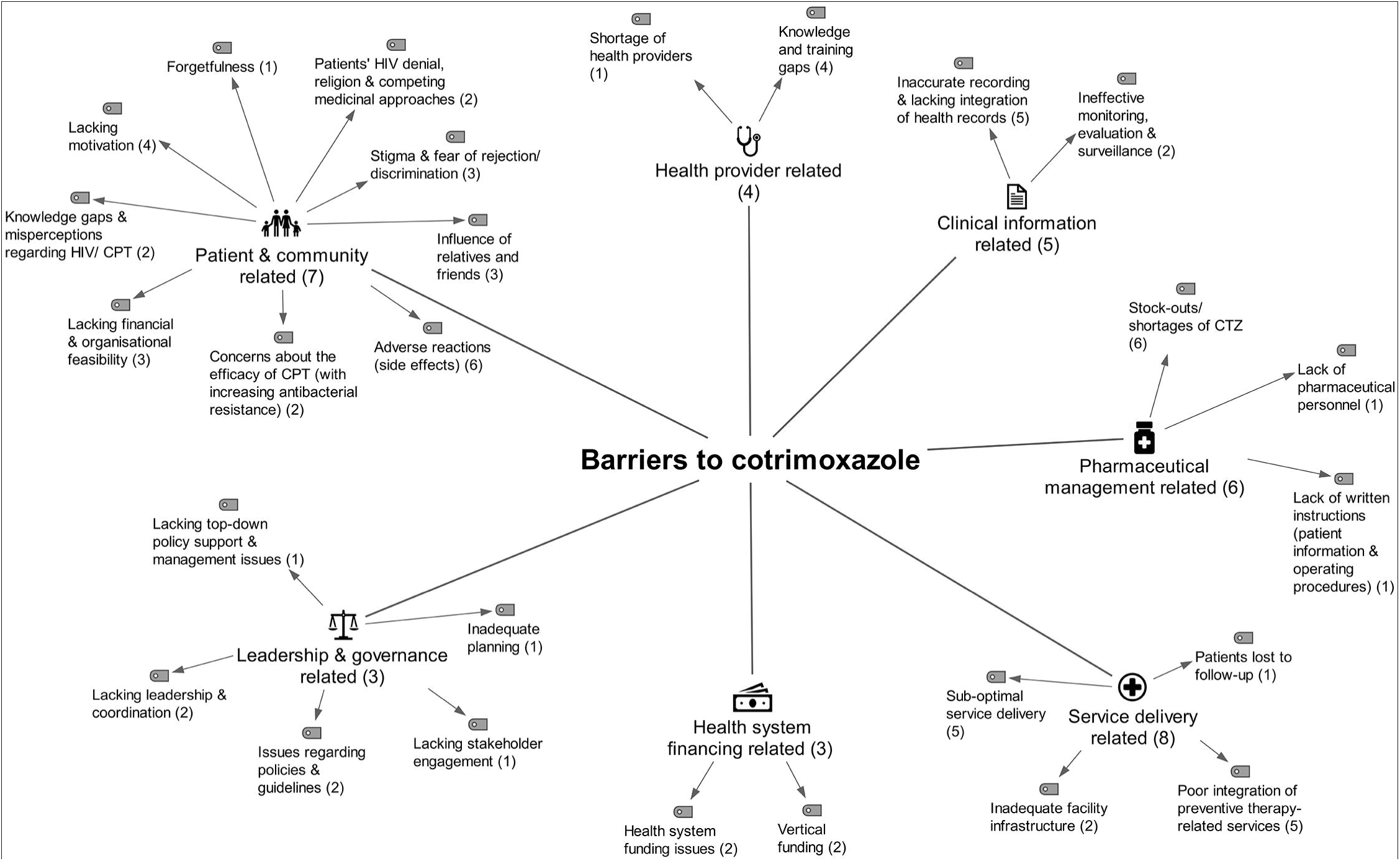
Metasummary: Barriers to cotrimoxazole preventive therapy for people living with HIV. The graphic presents all main barrier themes identified in countries with a high burden of tuberculosis and HIV, assigned to the health system component from which the barrier theme arose, considering a seven component health system framework (modification of the WHO Framework for Action, 2010). Barrier themes identified in more than or equal to ten papers are presented with an exclamation mark. Numbers in brackets display the number of supporting papers. Total number of peer-reviewed papers included with barriers to cotrimoxazole in this review (N= 11).

**Fig 4.**
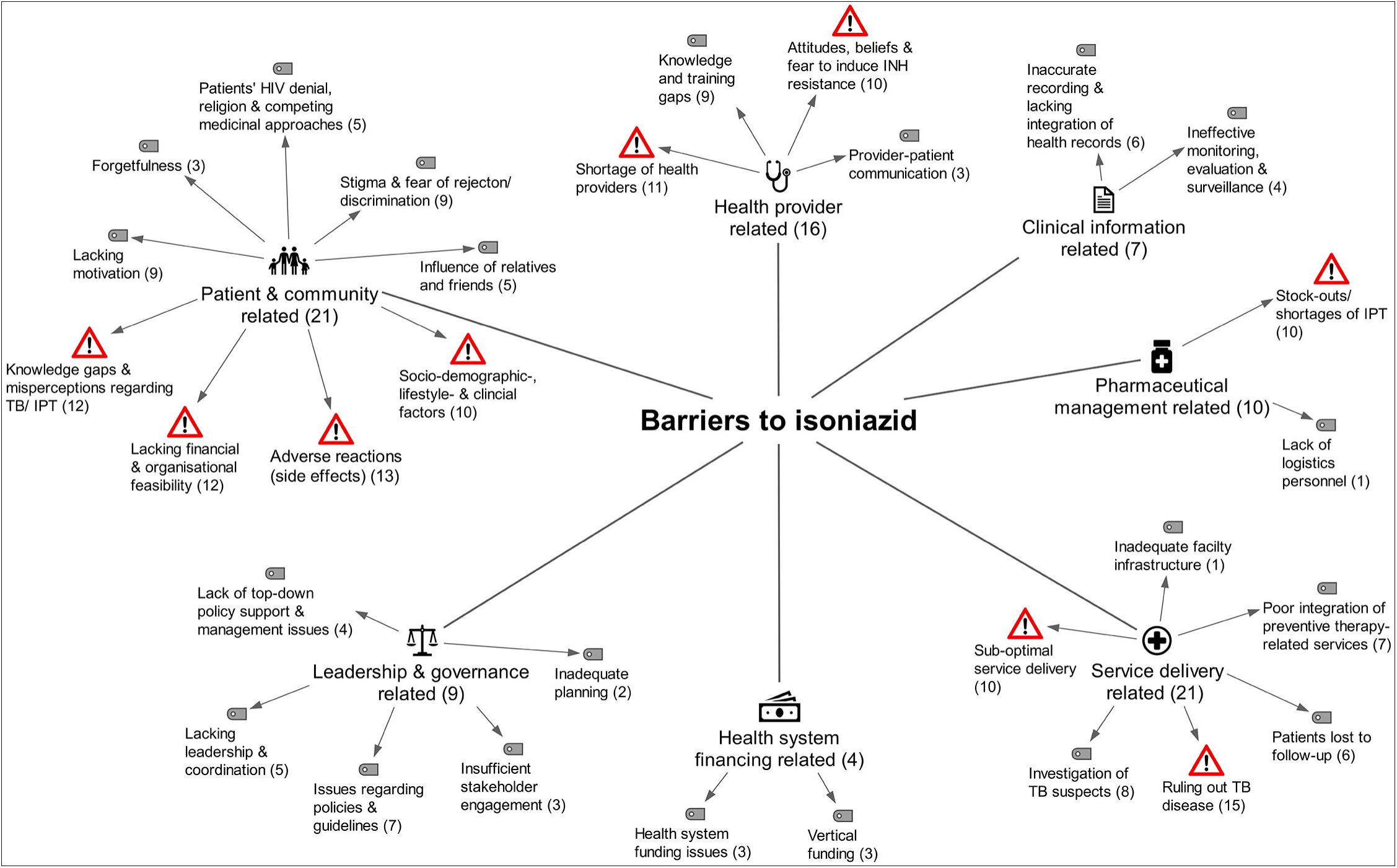
Metasummary: Barriers to isoniazid preventive therapy for people living with HIV. The graphic presents all main barrier themes identified in countries with a high burden of tuberculosis and HIV, assigned to the health system component from which the barrier theme arose, considering a seven component health system framework (modification of the WHO Framework for Action, 2010). Barrier themes identified in more than or equal to ten papers are presented with an exclamation mark. Numbers in brackets display the number of supporting papers. Total number of peer-reviewed papers included with barriers to isoniazid in this review (N= 28).

### Facilitators for the implementation of preventive therapies

We identified a set of strategies for enhancing the health facility-based implementation of preventive therapies for PLHIV. We presented our findings, according to the preventive therapy cascade - a coding framework that we developed a priori (Fig 5).

**Fig 5.**
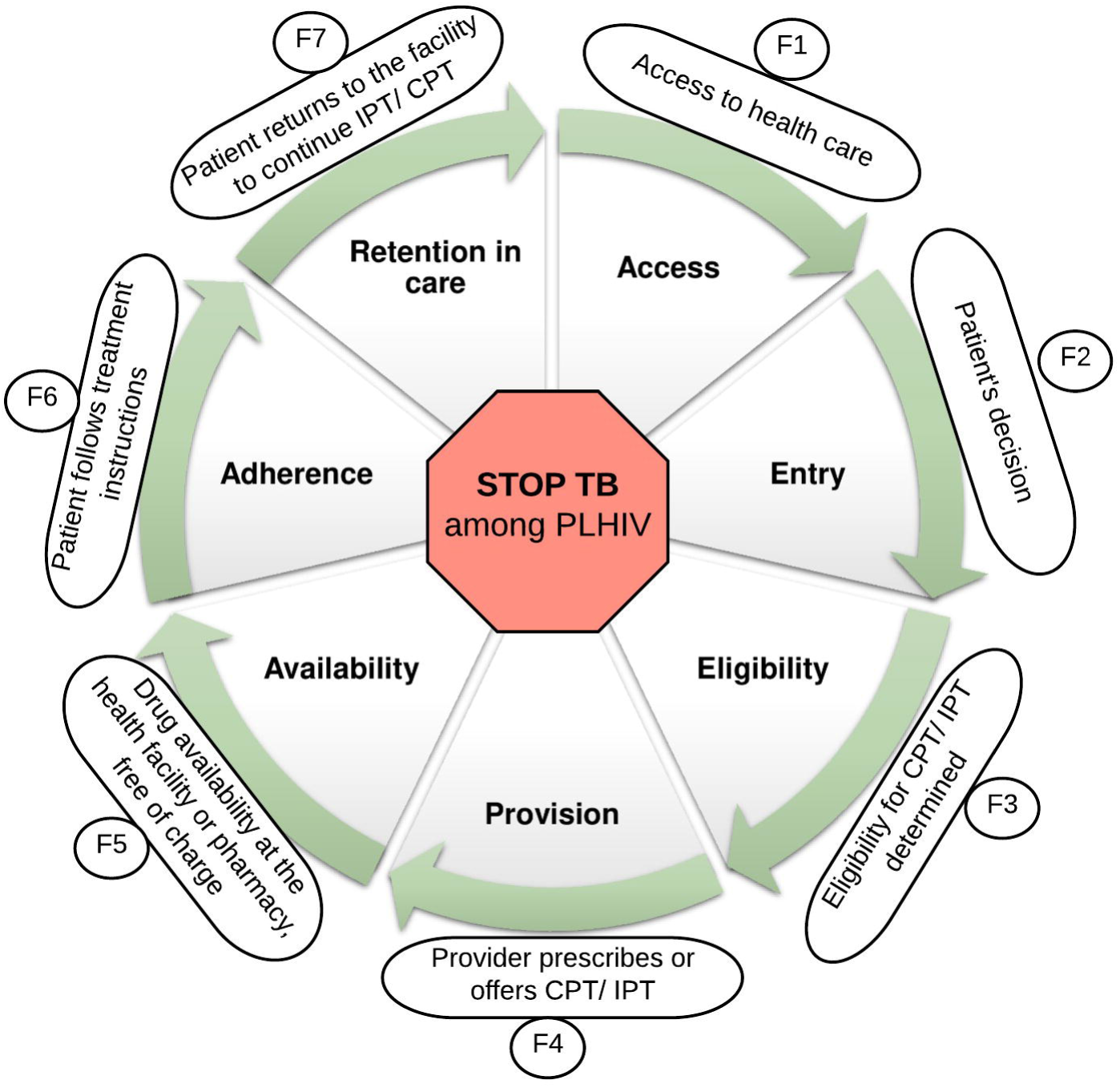
The preventive therapy cascade. TB - tuberculosis; HIV - human immunodeficiency virus; PLHIV- people living with HIV; IPT- isoniazid preventive therapy; CPT - cotrimoxazole preventive therapy.

The preventive therapy cascade outlines the steps that PLHIV repeatedly go through from having access to appropriate healthcare to completing preventive therapy. We developed this conceptual framework to promote stakeholder discussion about the ‘weak spots’ within the cascade in their specific setting. We listed the identified facilitators for the implementation of both preventive therapies accordingly. Therefore, this approach allows stakeholders (1) to select steps within the cascade that require attention, (2) to rapidly screen potential strategies that address these ‘weak spots’ within the cascade and (3) collaboratively choose a combination of strategies that are most appropriate in their specific context. For example, strategies to enhance patients’ adherence to PT’s are presented in section F6, shown in Fig 5. and Table 2.

**Table 2.**
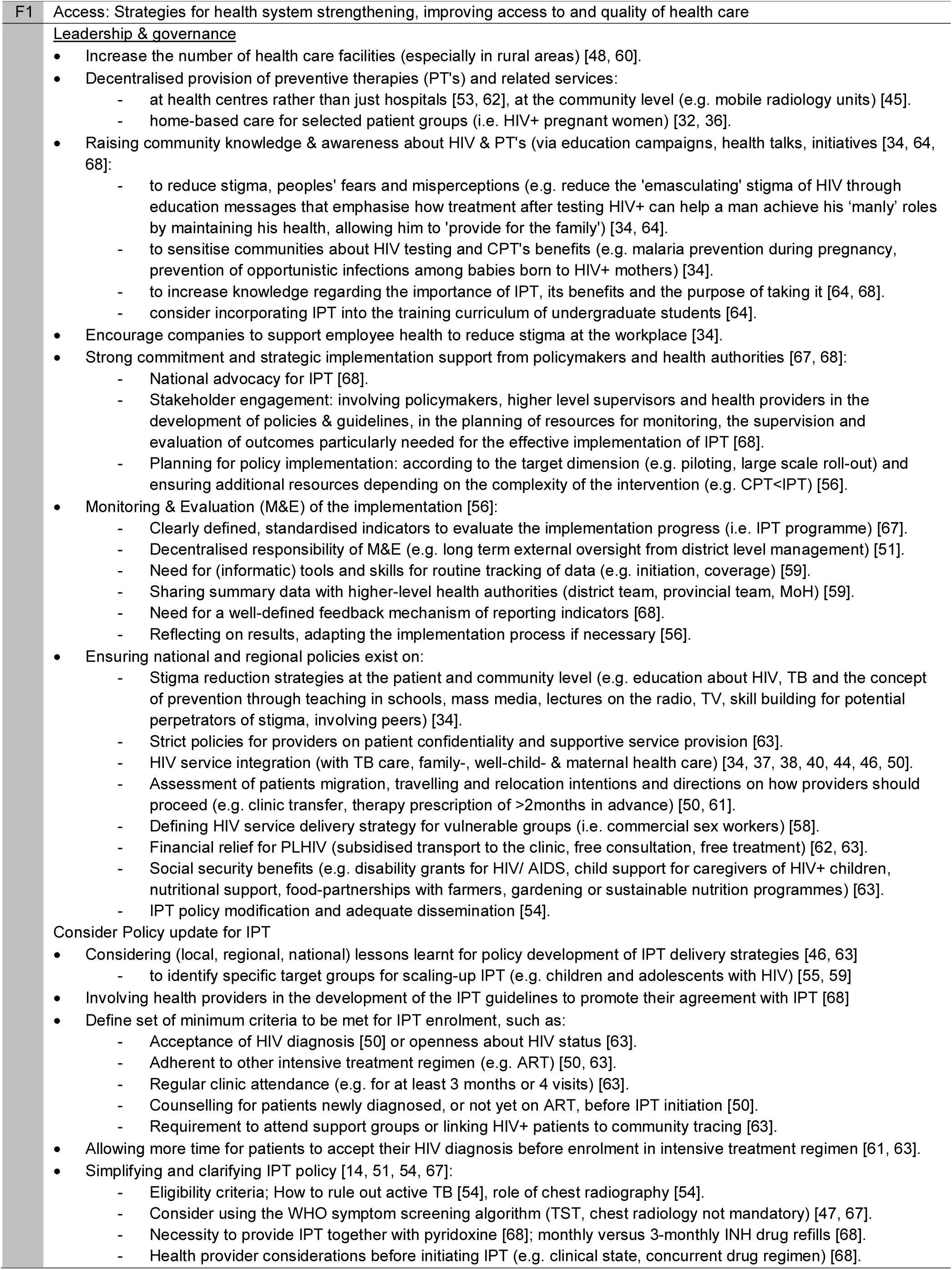
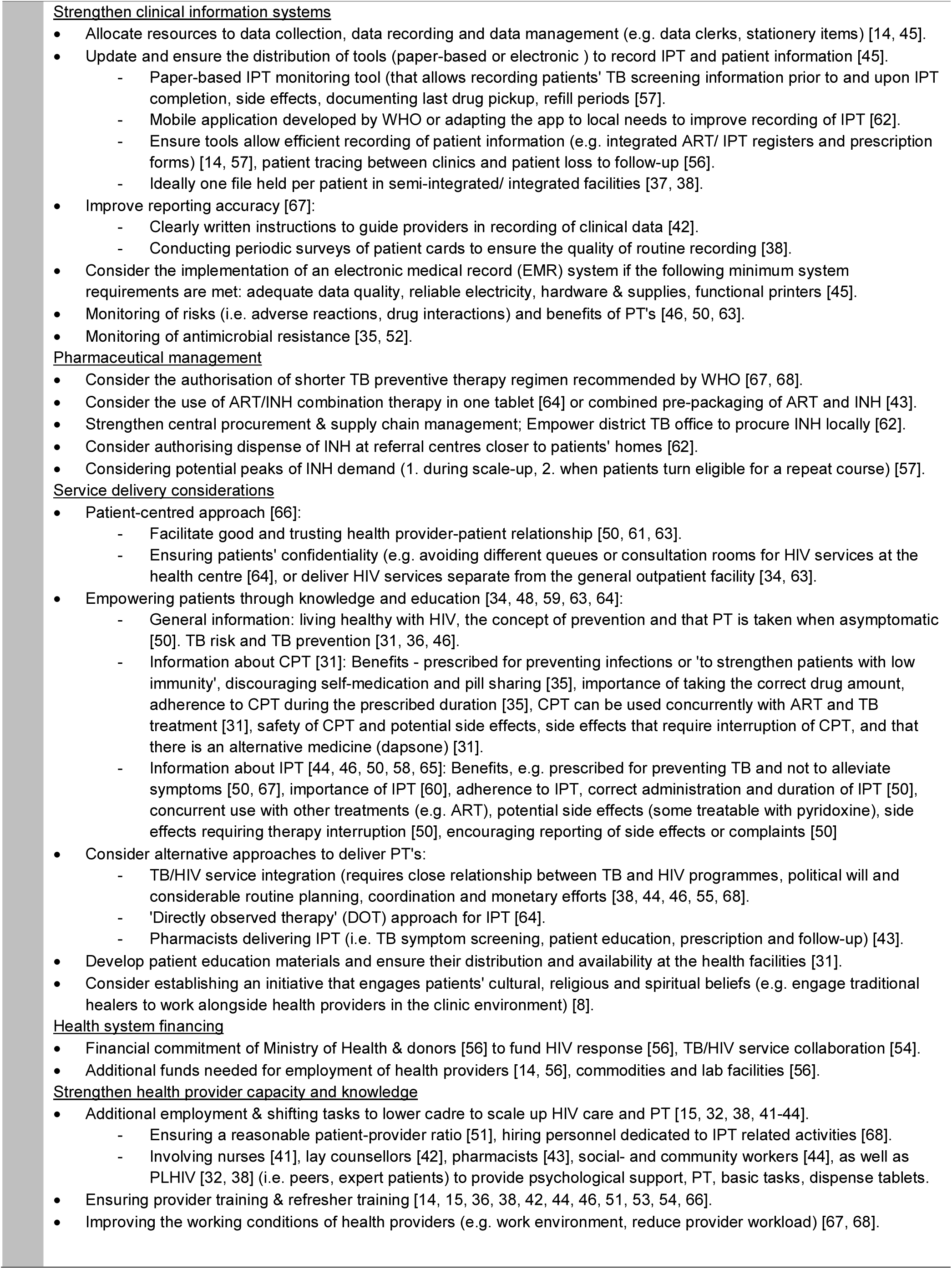
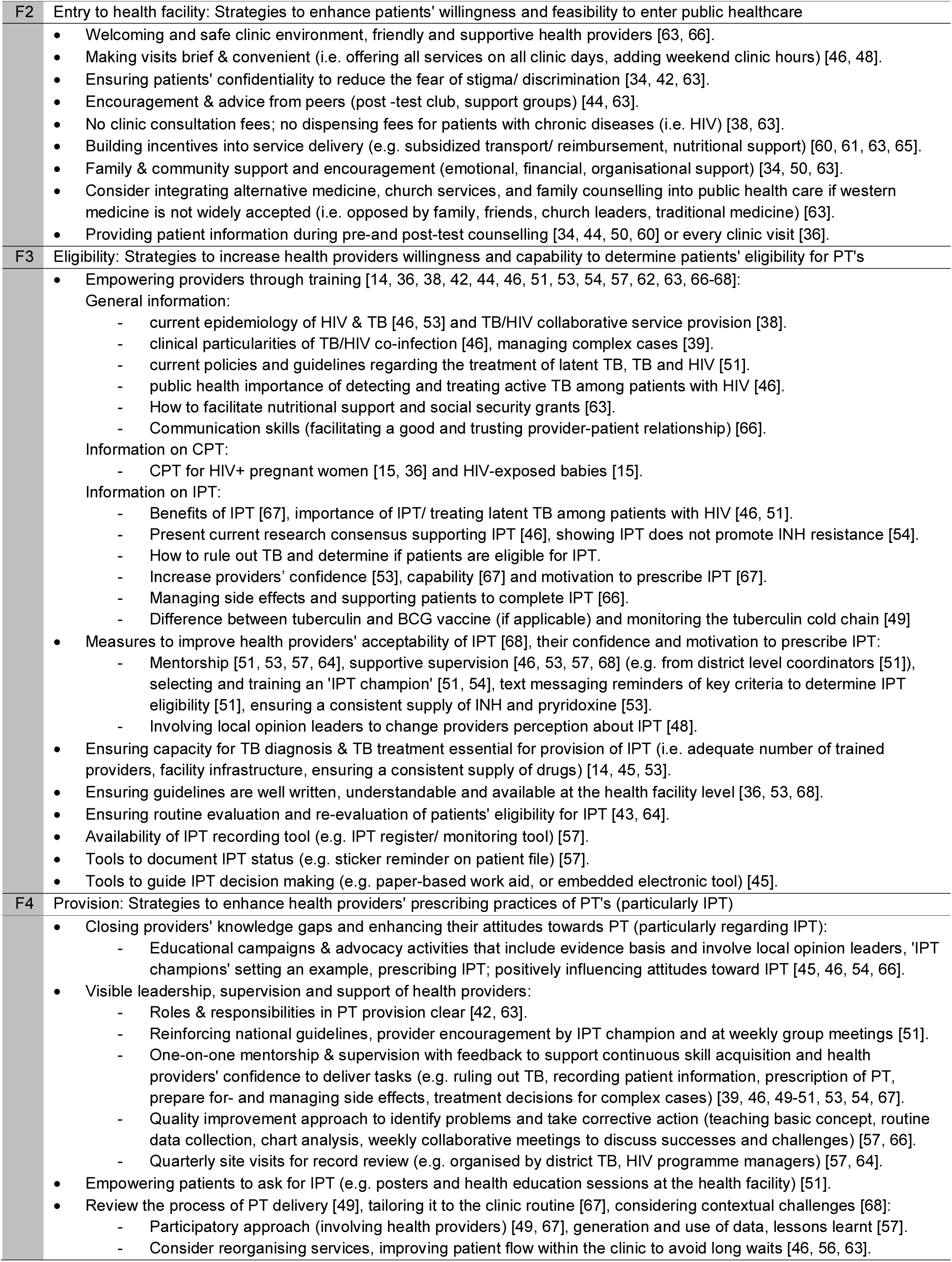
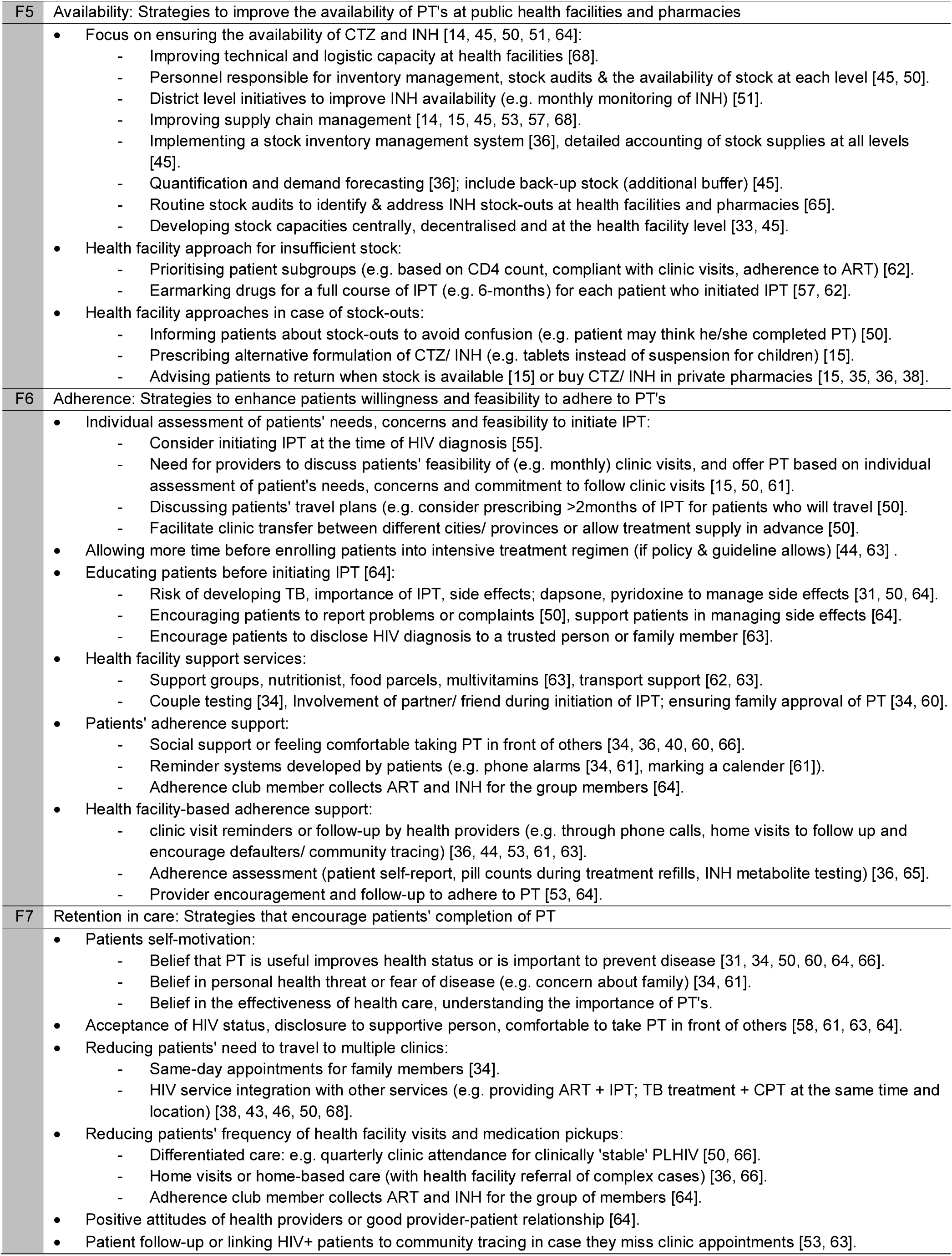
List of facilitators for the implementation of CPT and IPT.

## Discussion

In high TB/HIV burden countries, both preventive therapies in the centre of this review (i.e. CPT, IPT) are key interventions for PLHIV that can save lives if administered together with ART, or even on their own [6, 9]. However, governments of high TB/HIV burden countries face major implementation challenges that limit the effectiveness of both preventive therapies [16–18]. We explored and compared both preventive therapies with respect to similarities and differences of barriers identified across high TB/HIV burden countries published in peer-reviewed literature. Additionally, we explored and summarised strategies (facilitators) with the potential to tackle the identified barriers.

### Similarities and differences in barriers

Detailed barrier description presented in this review offered various explanations to why the implementation of both preventive therapies has been so challenging in the concerned countries. However, careful analysis of the extracted barrier information showed that many barriers were very similar for both preventive therapies. Our metasummary findings showed that the majority of barrier themes (25/32) that emerged from this review were identical for CPT and IPT and thus not intervention-specific (Figs 3 and 4). This leads to the question: “What are the underlying patterns of these cross-cutting barrier themes?” Taking into consideration that cross-cutting barrier themes were scattered across all building blocks of the health system and rather generic in nature indicates that systematic weaknesses within the health system may be an important underlying reason for implementation struggles. All barrier reporting studies (N=37), except one [52], reported such cross-cutting barriers, which at least hints that many high TB/HIV burden countries’ health systems were weak and unable to meet the basic requirements of a well-functioning health system [27]. Fragile public health systems could be explained by the fact that more than three-quarters of the countries with a high burden of TB and HIV are low- and lower-middle-income countries (as per 2021 World Bank’s classification) [69]. Many of these countries have severely constrained health budgets and historically depend on foreign assistance [70]. Following two decades of increasing domestic and generous international funding that has been mainly channelled into vertical health programs, impressive gains have been achieved in reducing AIDS-related deaths [27]. However, health systems strengthening is not characteristic for vertical funding and healthcare organisation. A major concern is that impressive gains of the past have been neither universal, nor sustainable [27]. We speculate that the sustainability of the HIV response and health facility-based delivery of both preventive therapies will become more challenging over the next years. The dilemma is that funding from international donors is flatlining [70], while an increasing number of HIV patients require life-long medical care [71] to be delivered by vertical programs that have long reached the limit of their effectiveness. Others have claimed that strengthening weak public health systems in the developing world is the next ‘emergency’ and an inevitable step to sustain the progress achieved in the last two decades [72]. At this stage of understanding, we believe it is quite possible that health system constraints identified for CPT and IPT, similarly hinder the implementation of other health facility-based interventions in high TB/HIV burden countries. Therefore, in addition to intervention-specific barriers, context-specific health system constraints should be considered to move towards a more feasible implementation strategy.

Like for CPT, barriers identified for IPT were most frequently classified as ‘service delivery-’ or ‘patient & community related’. Typically, ‘service delivery related barriers’ were interrelated with constraints identified across other health system components. For instance, with inaccurate clinical information, insufficient health providers and frequent drug stock-outs, efficient and reliable delivery of preventive therapy is unrealistic. These interrelations highlight that service delivery strongly depends on a well-functioning interplay of the health system components represented in the original WHO’s Framework for action [27]. We observed that barriers identified across the six original health system components (i.e. health workforce, information systems, essential medicines, financing, leadership/ governance, service delivery) represented constraints on the supply side. Meanwhile, ‘patient and community-related barriers’ comprised limitations that limited the demand of preventive therapies, hindering patients from entering healthcare, leading to inconsistent clinic visits or therapy interruptions. Among all ‘patient and community-related barriers’, we would like to draw special attention to ‘lacking financial and organisational feasibility’. This barrier theme was supported by more than one third of all studies included in this review [12, 15, 16, 22, 23, 25, 28, 31, 40–45], suggesting that in resource-constrained countries and in the context of poverty, free provision of health facility-based services may not be sufficient to ensure equitable access to healthcare. The economic impact of COVID-19 is expected to further decrease patients’ average household income in the coming years, posing an increasing threat to their ability to commit to frequent health facility visits and medication pickups. Summarising the above, barriers to both, CPT and IPT were most frequently classified as ‘service delivery-’ or ‘patient & community related’, representing supply- and demand-side constraints that introduce complex, interrelated dynamics [73]. Therefore, we emphasise the value of a ‘seven component health system framework’ for the purpose of barrier exploration and the formulation of an implementation strategy that is tailored to address potential barriers on the supply- and demand-side.

We also revealed some differences between barriers to both preventive therapies. Overall, our review suggests that implementing IPT has been more troublesome than CPT, supported by overall more barrier themes and evidence reporting barriers to IPT. This is consistent with the cross-sectional study carried out by WHO HIV/AIDS programme officers in 2007 [74], which showed that less progress had been made in implementing IPT when compared to CPT at the time. Thus, more research focus may have been on IPT since. Nevertheless, we argue that there is also more complexity involved in the eligibility assessment and provision of IPT compared to the more straightforward provision of CPT. Implementing IPT requires a higher degree of coordination; its delivery involves multiple steps, requires particular skills and seems to depend on additional capacity (particularly human resources) to follow-up patients who initiated IPT and those with a positive TB screening result.

Considering challenges specific to implementing IPT, ‘health provider related barriers’ appeared to play an important role. Our review revealed that having to rule out active TB disease was an important issue for the implementation of IPT. First, local policies and guidelines were described as unclear, so that health providers lacked knowledge and confidence in determining who is eligible for IPT [14, 68]. Second, health providers appeared to be overwhelmed with this additional task of routinely assessing and documenting patients’ eligibility [38, 51]. Third, while health providers attitudes towards CPT seemed neutral or positive, providers’ attitudes and beliefs toward IPT were partially negative [14, 44, 45, 53, 54, 62, 64, 67, 68]. One health provider concern was that extrapulmonary TB, common among PLHIV, may go undetected when screening for pulmonary symptoms [54]. Some health providers appeared to be unconvinced whether the benefits of IPT outweigh the risks of side effects [14, 53, 68] or of promoting INH resistance [45, 54, 64, 68]. Studies carried out in sub-Saharan Africa reported that some health providers had refused to prescribe IPT because of their fear of promoting INH resistance [54]. Overall, the use of preventive therapies has often been controversial, with concerns about drug resistance forming one major barrier to policy development and implementation [75]. However, it was somewhat surprising that studies reported health providers’ concern about bacterial resistance for IPT [45, 54, 68], but not for CPT. Notably, many high TB/HIV burden countries witness a high burden of MDR-TB [2]. Instead of INH monoresistance, health providers’ genuine concern may thus lay in promoting MDR-TB among patients with HIV [54]. Another surprising barrier theme identified specifically for IPT was provider-patient communication [50, 51, 58], which could indicate that communication needs are greater for IPT. However, only three studies reported issues with provider-patient communication regarding IPT [50, 51, 58]. Therefore, this barrier could also be indicative for changing patient expectations. A popular explanation is that the patient-provider relationship has changed over time. Health providers are no longer seen as the dominant decision-makers and patients as passive service recipients (i.e. paternalistic model) [76]. Instead, patients expect and are expected to be actively involved in their therapy decision [76]. Based on our review findings, patients were often afraid to speak to health providers about IPT [50, 51, 58], suggesting that providers may require additional training in how to develop a trusting provider-patient relationship and encourage their patients to address any arising concern. With the increasing global interest in TB prevention [2], several studies have been dedicated to assessing socio-demographic, lifestyle- and clinical factors associated with HIV patients’ chances of initiating, receiving, completing or adhering to IPT [14, 46, 48, 55, 58–60, 62, 66, 68]. These studies highlight key populations that may benefit from tailored implementation approaches. Another important factor for the implementation of IPT is that, in contrast to CPT, IPT requires greater coordination between HIV and TB program at all levels. Many high TB/HIV burden countries have implemented symptom-based screening to determine patients eligible for IPT [47], which is believed to have significantly advanced IPT uptake in low-resource settings [53]. On the one hand, symptom-based screening enabled HIV services units to take greater responsibility for implementing IPT. However, findings of the studies included in this review suggest that TB services’ capacity and linkage was too weak to follow-up patients with presumptive TB and to ensure immediate TB treatment for patients with confirmed TB [14, 38, 45, 49, 52, 54, 64, 67]. This leads to another challenge that is particular for IPT; it requires collaboration and capacity for the co-management of TB.

Considering challenges specific to the implementation of CPT, we found that bacterial resistance was a concern raised by the authors of a study included in our review [35]. Mwambete and Kamuhabwa (2016) questioned the efficacy of CTZ in areas of high bacterial resistance as a result of their research findings. Based on the disk diffusion method, their study identified an overall high resistance of isolated enteric *E. coli* among HIV patients in Tanzania [35]. Although antimicrobial resistance is a topic of utmost urgency, there are reasons to doubt the urgent need to reconsider the use of CPT in high TB/HIV burden countries. On the one hand, the in-vitro assay applied in the study [35] is no longer the gold standard for antimicrobial susceptibility testing because it often disqualifies antibiotics that are, in fact, effective in-vivo [77, 78]. On the other hand, several studies have shown significant reductions in morbidity and mortality among PLHIV on CPT, despite the fact that they were carried out in settings with high bacterial resistance. Therefore, others have argued that in-vitro resistance testing undermines the prophylactic ability of CTZ [79]. Finally, one study included in our review suggested a lack of written instructions for patients, referring to missing documented strategies for ensuring the availability of CTZ at the health facilities and patient leaflets that list potential adverse reactions that require therapy interruption [36]. Although we revealed several knowledge gaps and misperceptions among patients in our review that could be addressed in written instructions, the provision of patient information leaflets, in general, is not yet standard practice in many low- and middle-income countries [80]. One popular explanation for the under-utilisation of information leaflets at public pharmacies is that they are poorly understood [81]. Poor understanding may be more pronounced among populations with low literacy, which may have led to low prioritisation of creating such written information in the concerned countries. However, in combination with pictograms, basic written information has shown valuable for educating patients about CPT [80]. Nevertheless, such patient leaflets are still not available in all countries, reemphasising the importance of effective provider-patient communication during direct patient contact.

### Facilitators to preventive therapies

As barriers vary between and within high TB/HIV burden countries, there is not one strategy for improving the implementation of either or both preventive therapies that fits all settings. We, therefore, encourage redesigning an implementation strategy that addresses barriers identified in a specific context. The awareness of implementation challenges and political willigness to tackle these issues are critical first steps for continuous improvement. Routinely collected indicators, district-level evaluations, dialogues with implementers or local research findings may indicate difficulties with the implementation of one or both preventive therapies on a national level or in a specific setting. Our review identified only a few intervention-specific barriers and highlighted that cross-cutting barriers often affect the implementation of both preventive therapies and potentially other interventions in high TB/HIV burden countries. On the one hand, the resulting awareness that strategies are needed to target systematic weaknesses, as well as an intervention on its own, can be daunting. On the other hand, integrating the concept of health systems strengthening into an implementation strategy provides new opportunities for policymakers and health systems. These opportunities include improvements in health system sustainability, effectiveness and the availability of international support for health systems strengthening.

First, health systems strengthening may still appear unrealistic in low-income countries where primary care facilities struggle to provide basic health services and essential medicines. Additionally, funding mechanisms have partially encouraged vertical health system organisation and made it more difficult for health systems strengthening to be achieved in low- and middle-income countries. However, the decision for ‘organisational transformation’ is not based on the capacity of an individual primary care facility but rather, the potential behind removing cross-programmatic duplications and leveraging the existing resources. Transforming a health system structurally from fully vertical (e.g. disease-specific) programmes into a horizontally organised health system should be a gradual process. Similarly, others have argued that a phased, targeted and incremental transition from a vertical HIV response to a more efficient and sustainable health system response should be the way forward [71]. Adapting the implementation strategy of a high-impact multi-programmatic intervention, such as IPT, provides an opportunity to contribute to a more sustainable health system transformation.

Second, we understand that health systems strengthening is ambitious to achieve holistically, is difficult to measure and can be expected to take a long time. However, targeted strengthening of a selected health system component, such as up-skilling leadership and governance, reinforcing health providers’ capacity and knowledge, strengthening supply chains or health information systems comprise examples frequently identified in this review with the potential to yield benefits across both preventive therapies and a wide range of other health interventions. In addition, strengthening an individual health system component can help relax capacity constraints, result in cost-sharing, increase efficiency and cost-effectiveness of individual interventions. Cost-effectiveness analysis can assist in establishing the optimal balance between systems strengthening- and intervention-specific approaches to be included in an implementation strategy [82].

Third, vertical programmes of the past were popular for their ‘quantifying of lives saved’ approach that promised and short term objectives that promised quick and measurable results [83]. However, disease-specific programmes and initiatives were criticised for their costly parallel structures, less sustained impact, and for negatively affecting non-HIV care [72, 83]. In high TB/HIV burden countries that depend on external funds to finance their HIV response, vertical programming was associated with donor-driven policy evolution and investment decisions that were not always aligned with national priorities [56]. ‘Pushing’ global recommendations without considering context-specific constraints undermines the competence of local policymakers and can create tension between donors and recipient countries’ governments and weaken local policy ownership. For example, instead of prioritising prevention activities, donors’ investment focus was considered primarily strong on medicines, without adequate consideration of health system capacity (e.g. human resources) on which the delivery of these medicines is dependent [14, 56, 71].

However, many of these issues have been recognised internationally, and the global health systems strengthening movement has finally gained momentum [84, 85]. Major HIV funding bodies (i.e. USAID, Global Fund) have begun promoting the ‘de-verticalisation’ of fragmented programming [84, 86]. Both, the U.S. Agency for International Development (USAID) and the Global Fund to fight AIDS, TB and Malaria (Global Fund) have incorporated systems strengthening objectives into their strategy [84, 86] and updated their policies to include funding agreements for health systems strengthening that allow direct disbursement of funds to the recipient country’s state budget [83, 87]. Dialogues between donors and local system stakeholders (e.g. USAID’s Government to Government (G2G) assistance, Global Fund’s country coordinating mechanism (CCM)) support alignment of policies with country priorities [83, 87], encouraging mutually beneficial decisions and better relationships between donors and recipient countries. Additionally, the new implementation science culture encourages consideration of national, regional and local lessons learnt [46, 63, 88]. To sum up, the foundation for change has been laid. It is now up to high burden countries’ governments to make best use of the support and domestic resources available for health to collaboratively select, plan and coordinate an implementation strategy that addresses context-specific barriers and maximises the likelihood of long-term sustainability of both interventions.

### Strengths and limitations

The main strength of this review is the in-depth understanding that this review provides about the challenges associated with the implementation of CPT and IPT in countries with a high burden of TB and HIV. To explore the existing body of evidence holistically, (1) we applied a comprehensive search strategy, (2) included all stakeholders involved in the implementation process as study population, (3) integrated research with different study designs, and (4) presented the identified barriers within each of the building blocks of the health system. First, we systematically searched three databases, using a combination of MeSH and free-text terms. In addition, we manually screened reference lists of systematic reviews for original research eligible for this review. Second, our review included studies that reported the perspectives and experiences of any stakeholder involved in the implementation process, encompassing patients’ and health providers’ views that are otherwise often disregarded in the implementation process. Third, the integration of studies with different study designs further enhanced the richness of our findings. Mixed methods systematic reviews have gained increasing scientific attention in the areas of public health and complex interventions, where typically decision makers require consideration of different types of data and information (e.g. accessibility, feasibility, patient values and preferences) [18]. Fourth, for the qualitative evidence synthesis of barrier information, we applied a highly transparent deductive approach using an a priori defined coding framework of seven health system components (a modification of the WHO’s Framework for action) [27]. Qualitative evidence synthesis is relatively new compared to quantitatively oriented systematic reviews and the application of metasummary to compare interventions with respect to similarities and differences of barriers employed in our review is innovative.

Potential limitations of our review include (1) restrictions in our search strategy, (2) methodological limitations of the included studies, (3) heterogeneity of study characteristics, and (4) potential biases introduced through the non-availability of studies or the non-dissemination of study findings. First, although our search strategy was systematic and rigorous, it only included peer-reviewed publications reported in the English language. This is unlikely to be a major limitation considering that twenty-three high burden countries were represented in this review. However, this may explain why most studies included in this review, were carried out in South Africa, Uganda and Tanzania. Only including peer-reviewed published studies may have led to the exclusion of potentially useful grey reports (e.g. HIV progress reports). Second, we did not exclude any study from our analysis regardless of the methodological limitations. However, we systematically assessed each study’s strengths and limitations and carefully considered issues of bias, validity, and trustworthiness. Third, we identified a heterogeneous representation of the study populations represented in our review. Most of the included studies presented the perspectives of patients and health providers. Consequently, the views of caregivers, community members and other stakeholders involved in the implementation process were less frequently represented, limiting the generalisability of our findings. However, generalising findings was outside the scope of this review. Therefore, the primary concern about this observation is the extent to which our findings holistically represent the phenomena of interest. Fourth, non-availability or non-dissemination of studies may have limited the completeness of our study findings [30]. Studies on the subject of this review may be less likely to be available in countries with limited political interest to implement the concerned preventive therapies. In high TB/HIV burden countries with little research capacity, available study findings may be more likely to be published in local journals that are not indexed in major databases.

## Conclusion

There is little benefit to any highly efficacious therapy as long as we are unable to effectively implement it under ‘real-world’ conditions.

For policymakers who encounter challenges with the implementation of either or both preventive therapies, this review offers a list of strategies for improving the implementation of both preventive therapies. Based on evidence from high TB/HIV burden countries, this list includes specific direction for improving the delivery of IPT (or any newer therapy regimen) for the prevention of TB. For researchers with limited working experience in high TB/HIV burden countries, this review can provide useful insights regarding to which barriers may arise at different levels of the health system. The barrier description provided in this review highlights the complexity of social interactions involved in the delivery of preventive therapies.

Overall, our review showed that until today, many high TB/HIV burden countries’ health systems are not prepared to ensure appropriate public healthcare for the ever increasing number of patients in need of HIV services. From this standpoint, health system strengthening is imperative for the sustainable delivery of both preventive therapies for PLHIV, but particularly for IPT. Based on our findings, we suggest consideration of two important aspects to implementation. First, we recommend early engagement of stakeholders when shaping an implementation strategy. Involving patients and health providers in the process of policy development and planning may increase awareness and understanding about the intervention and help to ensure its acceptability. Additional stakeholders (e.g. church leaders, traditional healers) may be involved, if appropriate. Second, during the adaptation of an implementation strategy, we urge that attention is paid to the fact that many of the countries with the highest burden of TB and HIV to date are also among the most resource-constrained countries. Thus, novel strategies for the implementation of preventive therapies may appear encouraging at first, but in high TB/HIV burden settings, they are often far from feasible or sustainable on a big scale. Instead, we encourage innovative approaches that consider the resources available at the health facility level, and investing into strengthening the existing capacities.

Future research is warranted to test and evaluate alternative service delivery approaches that reduce the financial and organisational burden that patients face during the course of preventive therapy. Replacing INH with one of the shorter treatment regimen recommended by WHO for the prevention of TB [13] and the community-based delivery of selected activities related to the provision of PT’s may comprise two options with the additional benefit of reducing the patient load at the overburdened health facilities.

## Supporting information

S1 Additional file. PICo Framework (Modified PICO).

S2 Additional file. Systematic review protocol.

S3 Additional file. PRISMA checklist.

S4 Additional file. Database searches and results.

S5 Additional file. Mixed method quality appraisal tool (MMAT) and user guide.

S6 Additional file. Detailed description of studies included in this review.

S7 Additional file. Assessment of methodological strengths and limitations.

S8 Additional file. Metasummary tables.

## Data Availability

Yes, all data are fully available without restriction. All relevant data are within the paper and its Supporting information files. 
Peer-reviewed studies included in our review can be obtained through either MEDLINE, Web of Science or Scopus database searching. 

https://www.crd.york.ac.uk/prospero/display_record.php?RecordID=137778

## Acknowledgement

We would like to express our gratitude to Jonathan O’ Mahony for proofreading the manuscript. In addition, we would like to thank Carlos Rodrigues, IT specialist at the Institute of Hygiene & Tropical Medicine in Lisbon for his support with formatting the graphics included in this manuscript.

## Supporting information

### Additional files

S1 Additional file. PICo Framework (Modified PICO).

S2 Additional file. Systematic review protocol.

S3 Additional file. PRISMA checklist.

S4 Additional file. Database searches and results.

S5 Additional file. Mixed method quality appraisal tool (MMAT) and user guide.

S6 Additional file. Detailed description of studies included in this review.

S7 Additional file. Assessment of methodological strengths and limitations.

S8 Additional file. Metasummary tables.

