## Supplementary material for "Mixed methods systematic review and metasummary about barriers and facilitators for the implementation of cotrimoxazole and isoniazid – preventive therapies for people living with HIV": S1 Additional file. PICo Framework (Modified PICO).

Additional file 1. PICo Framework (Modified PICO).

| | |
|---|---|
| **P**opulation | **TB/HIV high burden countries as per WHO for the period 2016 to 2020**<br>• Angola<br>• Botswana<br>• Brazil<br>• Cameroon<br>• The central African Republic<br>• Chad<br>• China<br>• Congo<br>• Democratic Republic of the Congo, DR Congo, DRC<br>• Ethiopia<br>• Ghana<br>• Guinea-Bissau<br>• India<br>• Indonesia<br>• Kenya<br>• Lesotho<br>• Liberia<br>• Malawi<br>• Mozambique<br>• Myanmar<br>• Namibia<br>• Nigeria<br>• Papua New Guinea, PNG<br>• South Africa<br>• Swaziland<br>• Thailand<br>• Uganda<br>• United Republic of Tanzania, UR Tanzania, Tanzania<br>• Zambia<br>• Zimbabwe |
| **I**nterest | **Barriers and Facilitators**<br>barrier*, facilitator* |
| **Co**ntext | **Cotrimoxazole preventive therapy (CPT), Isoniazid preventive therapy (IPT)**<br>Cotrimoxazol*, Trimethoprim, Sulfamethoxazole Drug Combination, Isoniazid |

Note: The asterisk (*) is used as a truncation indicator.
