## Supplementary material for "Mixed methods systematic review and metasummary about barriers and facilitators for the implementation of cotrimoxazole and isoniazid – preventive therapies for people living with HIV": S2 Additional file. Systematic review protocol.

### 1. * Review title.

Give the title of the review in English

Barriers and facilitators for cotrimoxazole and isoniazid in high burden countries for human immunodeficiency virus and tuberculosis. A systematic review and metasummary of qualitative findings from peer-reviewed literature published

### 2. Original language title.

For reviews in languages other than English, give the title in the original language. This will be displayed with the English language title.

### 3. * Anticipated or actual start date.

Give the date the systematic review started or is expected to start.

01/05/2018

### 4. * Anticipated completion date.

Give the date by which the review is expected to be completed.

31/07/2019

### 5. * Stage of review at time of this submission.

Tick the boxes to show which review tasks have been started and which have been completed. Update this field each time any amendments are made to a published record.

**Reviews that have started data extraction (at the time of initial submission) are not eligible for inclusion in PROSPERO**. If there is later evidence that incorrect status and/or completion date has been supplied, the published PROSPERO record will be marked as retracted.

This field uses answers to initial screening questions. It cannot be edited until after registration.

The review has not yet started: No

| Review stage | Started | Completed |
|---|---|---|
| Preliminary searches | No | No |
| Piloting of the study selection process | Yes | Yes |
| Formal screening of search results against eligibility criteria | Yes | No |
| Data extraction | Yes | No |
| Risk of bias (quality) assessment | No | No |
| Data analysis | No | No |

Provide any other relevant information about the stage of the review here.

### 6. * Named contact.

The named contact is the guarantor for the accuracy of the information in the register record. This may be any member of the review team.

Pia Mueller

### Email salutation (e.g. "Dr Smith" or "Joanne") for correspondence:

Mrs Pia Mueller

### 7. * Named contact email.

Give the electronic email address of the named contact.

### 8. Named contact address

Give the full institutional/organisational postal address for the named contact.

Instituto de Higiene e Medicina Tropical (IHMT), Rua Junqueira 1001349-008 Lisbon

### 9. Named contact phone number.

Give the telephone number for the named contact, including international dialling code.

+351935251055

### 10. * Organisational affiliation of the review.

Full title of the organisational affiliations for this review and website address if available. This field may be completed as 'None' if the review is not affiliated to any organisation.

Instituto de Higiene e Medicina Tropical (IHMT), Lisboa

### Organisation web address:

https://www.ihmt.unl.pt/

### 11. * Review team members and their organisational affiliations.

Give the personal details and the organisational affiliations of each member of the review team. Affiliation refers to groups or organisations to which review team members belong. **NOTE: email and country now MUST be entered for each person, unless you are amending a published record.**

Mrs Pia Müller. Instituto de Higiene e Medicina Tropical Lisboa (IHMT)
Professor Luís Velez Lapão. Instituto de Higiene e Medicina Tropical Lisboa (IHMT)

### 12. * Funding sources/sponsors.

Details of the individuals, organizations, groups, companies or other legal entities who have funded or sponsored the review.

Pia Mueller is a full-time PhD research fellow, funded by the Hans-Böckler Foundation.

Luís Velez Lapão is a full-time Professor and Researcher employed at the Instituto de Higiene e Medicina Tropical.

### Grant number(s)

State the funder, grant or award number and the date of award

### 13. * Conflicts of interest.

List actual or perceived conflicts of interest (financial or academic).

None

### 14. Collaborators.

Give the name and affiliation of any individuals or organisations who are working on the review but who are not listed as review team members. **NOTE: email and country must be completed for each person, unless you are amending a published record.**

### 15. * Review question.

State the review question(s) clearly and precisely. It may be appropriate to break very broad questions down into a series of related more specific questions. Questions may be framed or refined using PI(E)COS or similar where relevant.

The research question “Which are the barriers and facilitators to preventive treatment with cotrimoxazole and isoniazid in TB/HIV high burden countries?” was designed using the PICo framework: Population, Interest, Context.

Key concepts:

- Preventive treatments: by nature aim to prevent, rather than treat disease.
- Cotrimoxazole preventive treatment (CPT): see intervention section for more details
- Isoniazid preventive treatment (IPT): see intervention section for more details
- Barriers: are defined as negative factors that limit, challenge, or inhibit access, provision, delivery, implementation or adherence to CPT or IPT.
- Facilitators: are defined as positive factors that facilitate, support, encourage, or enable the access,

provision, delivery, implementation or adherence to CPT or IPT.

- High burden countries for TB/ HIV: 30 countries as per WHO definition for the period 2016-2020.
- WHO Framework for action (2010): framework previously adapted by authors (Getahun and Granich et al 2010) to present barriers and solutions for the implementation of IPT. The original framework describes health systems in terms of six core components: (1) service delivery, (2) health workforce, (3) health information systems, (4) access to essential medicines, (5) financing, and (6) leadership/governance.

The primary objective of this review is to systematically search for, appraise, synthesize, and present health system barriers and facilitators for both preventive treatments (Cotrimoxazole, Isoniazid) in TB/ HIV high burden countries.

The secondary objective is to assess and discuss the relative magnitude of our findings using Sandelowski and Barroso´s aggregation method. (Metasummary)

### 16. * Searches.

State the sources that will be searched (e.g. Medline). Give the search dates, and any restrictions (e.g. language or publication date). Do NOT enter the full search strategy (it may be provided as a link or attachment below.)

Peer-reviewed literature will be systematically identified through MEDLINE, Web of Science and Scopus databases searching. Because of the limited number of qualitative studies published on this topic, we included primary studies of any design (qualitative, quantitative, mixed-method).

Inclusion criteria were:

- Language of publication: English
- Years published: all articles published until September 2020
- Countries of interest: all (thirty) TB/HIV high burden countries defined by WHO in the period 2016 to 2020
- Types of Data sources: Full-text peer-reviewed scientific publications, primary data, reference lists of systematic reviews will be screened for additional publications if the systematic review is relevant to the study question.

The PRISMA Flow Diagram will be applied to present the number of papers included throughout the selection process.

### 17. URL to search strategy.

Upload a file with your search strategy, or an example of a search strategy for a specific database, (including the keywords) in pdf or word format. In doing so you are consenting to the file being made publicly accessible. Or provide a URL or link to the strategy. Do NOT provide links to your search **results**.

https://www.crd.york.ac.uk/PROSPEROFILES/137778_STRATEGY_20190605.pdf

Alternatively, upload your search strategy to CRD in pdf format. Please note that by doing so you are consenting to the file being made publicly accessible.

Do not make this file publicly available until the review is complete

### 18. * Condition or domain being studied.

Give a short description of the disease, condition or healthcare domain being studied in your systematic review.

This review focusses on two infectious diseases, Tuberculosis (TB) and the Human immunodeficiency virus (HIV).

In 2017, worldwide 36.9 million people were living with the HIV (UNAIDS 2018). An estimated quarter of the world's population was latently infected with mycobacterium tuberculosis, of which 10 million developed active Tuberculosis (Global TB Report 2018).

The fact that HIV positive patients have a 20 to 30 times higher risk of developing active TB disease compared to HIV negative patients has been described as deadly synergy (Kwan and Ernst 2011) which has transformed many low- and middle-income countries into TB/ HIV high-burden countries, where TB remains the leading cause of death among PLWH, accounting for around one in three AIDS-related deaths (UNAIDS Global HIV Statistics 2017).

### 19. * Participants/population.

Specify the participants or populations being studied in the review. The preferred format includes details of both inclusion and exclusion criteria.

This review focuses on TB/ HIV high burden countries as defined by WHO for the period 2016 to 2020, listed below. At highest risk of opportunistic infections and of acquiring Tuberculosis disease are those individuals living in regions with a high burden of both infectious diseases, HIV and Tuberculosis. High burden countries share similar disease profiles and partially similar economic and cultural contexts, offering an opportunity to compare and share their lessons learnt.

30 TB/ HIV high burden countries:

Angola

Botswana

Brazil

Cameroon

Central African Republic

Chad

China

Congo

DR Congo

Ethiopia

Ghana

Guinea-Bissau

India

Indonesia

Kenya

Lesotho

Liberia

Malawi

Mozambique

Myanmar

Namibia

Nigeria

Papua New Guinea

South Africa

Swaziland

Thailand

Uganda

UR Tanzania

Zambia

Zimbabwe

To explore potential barriers and facilitators from different perspectives of the health system a broad approach will be applied including any study population reporting barrier(s) or facilitator(s) for any of the two preventive treatments.

Study population; the intended recipients of preventive treatments:

- CPT: patients with HIV or TB-HIV co-infection, including HIV-exposed babies (HIV suspects).

- IPT: patients with HIV, excluding HIV-negative high-risk groups for active TB (i.e. contacts of TB index cases).

• Health care providers

• Caregivers (parents or guardians of child patients) involved in the preventive treatment collection or administration process

• Any other stakeholder identified as influential in the overall implementation process of IPT, CPT or both

### 20. * Intervention(s), exposure(s).

Give full and clear descriptions or definitions of the interventions or the exposures to be reviewed. The preferred format includes details of both inclusion and exclusion criteria.

This review focusses on primary studies that report health system barriers and/ or facilitators to either of the two most important preventive treatments in the TB/ HIV high burden context:

Cotrimoxazole (sulfamethoxazole/ trimethoprim 800mg/ 160mg daily) preventive treatment (CPT) is an efficacious intervention for the prevention of opportunistic infections, while Isoniazid (300mg daily) preventive treatment (IPT) has shown successful in preventing Tuberculosis, which is particularly important for people living with HIV (PLHIV) due to their twenty to thirty times increased risk of developing active TB disease compared to people without HIV (WHO 2017).

In more than two decades, clinical trials and observational studies have shown that both preventive treatments have been associated with significantly reduced morbidity and mortality among PLHIV, resulting in current World Health Organization (WHO) recommendations to include both preventive treatments as a standard package of care for this patient group and setting (WHO Guidelines 2014, 2018).

Governments of high burden countries face major challenges in implementing both treatments, particularly due to pharmaceutical supply issues, patients' attitudes and service delivery issues (Rapid review carried out by Pia Müller prior to the development of this protocol, 2018).

### 21. * Comparator(s)/control.

Where relevant, give details of the alternatives against which the intervention/exposure will be compared (e.g. another intervention or a non-exposed control group). The preferred format includes details of both inclusion and exclusion criteria.

None

### 22. * Types of study to be included.

Give details of the study designs (e.g. RCT) that are eligible for inclusion in the review. The preferred format includes both inclusion and exclusion criteria. If there are no restrictions on the types of study, this should be stated.

Metasummary is quantitatively oriented technique to aggregate qualitative findings.

In other words, this study applies a highly transparent aggregative process to identify studies which utilize QUALITATIVE, MIXED-METHODS and QUANTITATIVE methodologies, extract evidence from studies included in this review, to then categorize and synthesize these as qualitative findings.

Studies to be included may focus on:

• Barriers or facilitators of CPT and/or IPT (obvious findings)

• Implementation or provision of TB/ HIV collaborative services (less obvious findings)

• Implementation or provision of HIV services (less obvious findings)

• Implementation or provision of TB services (less obvious findings)

Implementation related studies (e.g. evaluation study, implementation study, pilot study) commonly present an increase or decrease in coverage, prescription or adherence to preventive treatment. In this scenario the intervention can result as either a barrier or a facilitator.

### 23. Context.

Give summary details of the setting or other relevant characteristics, which help define the inclusion or exclusion criteria.

The context of each study included in the systematic review will be recorded (e.g. urban, rural, sub-urban). (see data extraction).

### 24. * Main outcome(s).

Give the pre-specified main (most important) outcomes of the review, including details of how the outcome is defined and measured and when these measurement are made, if these are part of the review inclusion criteria.

Barriers to or facilitator for preventive treatments (Cotrimoxazol, Isoniazid)

#### * Measures of effect

Please specify the effect measure(s) for you main outcome(s) e.g. relative risks, odds ratios, risk difference, and/or 'number needed to treat.

Not applicable

### 25. * Additional outcome(s).

List the pre-specified additional outcomes of the review, with a similar level of detail to that required for main outcomes. Where there are no additional outcomes please state ‘None’ or ‘Not applicable’ as appropriate to the review

None

#### * Measures of effect

Please specify the effect measure(s) for you additional outcome(s) e.g. relative risks, odds ratios, risk difference, and/or 'number needed to treat.

Not applicable

### 26. * Data extraction (selection and coding).

Describe how studies will be selected for inclusion. State what data will be extracted or obtained. State how this will be done and recorded.

During the first selection the citations of all potentially eligible studies will be downloaded into EndnoteX9 reference management software, and titles and abstracts will be screened in duplicate by two researchers according to the predefined inclusion criteria (see searches section).

Duplicates, non-relevant studies, systematic reviews, and studies that report from countries other than the thirty high TB/HIV burden countries will be excluded during the initial screening process. The second selection will be based on the full-text of articles identified during initial screening.
Publications will be included by P.M. if they report barriers to or facilitators for CPT and/ or IPT in the result or findings section of the publication. The article screening process will be reviewed by LL. In the case of disagreement, full-text analyses will be discussed until consensus is reached. The PRISMA Flow Diagram will be applied to present the number of papers included throughout the selection process (PRISMA 2009).

An extraction table will be developed and used to record study characteristics of the included studies:
(1) First Author
(2) Year of publication (all scientific papers published until September 2020)
(3) Country of study (one or multiple countries defined as high burden country for TB/ HIV)
(4) Study Population (e.g. patients, caregivers, providers, health facilities)
(5) sample size (e.g. 590 TB patients)
(6) Research design and data collected (qualitative, quantitative, mix)
(7) Study aim
(8) Data Collection approach (e.g. interview, focus group discussion)
(9) Context (e.g. urban, sub-urban rural area, /if information provided)
(10) Prophylaxis concerned (Cotrimoxazol and/ or Isoniazid)
(11) Finding (barrier and/ or facilitator)
(12) Quality score (to indicate methodological quality of the study/ see section risk of bias)

The extraction table will be tested on three studies to ensure all relevant characteristics can be extracted. The abstraction process aims to preserve the meaning and complexity of the findings in the original study to optimize the validity.

### 27. * Risk of bias (quality) assessment.

State which characteristics of the studies will be assessed and/or any formal risk of bias/quality assessment tools that will be used.

1. Assessment of the quality of included studies

Eligible studies will be critically appraised by two independent reviewers (PM and LL) for methodological quality applying an adapted tool that incorporates items from the “Mixed methods appraisal tool (MMAT 2018)” and the NICE Quality appraisal checklist (National Institute for Clinical Excellence 2012). Each reviewer will provide an overall rating for the methodological quality according to the list below:

++ All or most of the checklist criteria have been fulfilled, where they have not been fulfilled the conclusions are very unlikely to alter.

+ Some of the checklist criteria have been fulfilled, where they have not been fulfilled, or not adequately described the conclusions are unlikely to alter.

- Few or no checklist criteria have been fulfilled and the conclusion are likely, or very likely to alter.

The appraisal tool will be tested on three studies to ensure feasibility, applicability of the instrument, and utility in assessing the overall quality of each study.

Any disagreement between the reviewers will be resolved through discussion, or with a third reviewer.

No studies will be excluded on the basis of quality assessment.

### 28. * Strategy for data synthesis.

Describe the methods you plan to use to synthesise data. This **must not be generic text** but should be **specific to your review** and describe how the proposed approach will be applied to your data. If meta-analysis is planned, describe the models to be used, methods to explore statistical heterogeneity, and software package to be used.

A narrative approach will be used to abstract the information, initially recorded on an Excel spread sheet (abstraction table) and eventually imported and synthesized using the qualitative data analysis software MAXQDA Analytics Pro (Release 18.1.1.).

A convergent integrated approach will be applied, to combine findings from qualitative, quantitative and mixed-methods studies into one set of qualitative data. The following example will demonstrate how a facilitator should be obtained from a quantitative study.

"An observational study publication shows that health facilities that carried out stock-counts twice per month reported 20% decrease in stock-outs (p0.05)."

The result (outcome of interest) to be abstracted:

- Facilitator: stock-counts twice per month at the health facility level to reduce (overcome) stock-outs (as barrier).

This review is therefore disregarding the effect size (here 20% decrease in stock-outs).

Through several rounds of coding, both reviewers will identify patterns among the extracted results based on similarity in meaning and thematically synthesize (define and refine the code system). The resulting themes will be assigned to the health system component most closely related to each barrier category (e.g. Access to essential medicines).

However, health system components may be modified to best represent the resulting themes. Thus, the component "access to essential medicines" may be renamed "pharmaceutical supply".

### 29. * Analysis of subgroups or subsets.

State any planned investigation of 'subgroups'. Be clear and specific about which type of study or participant will be included in each group or covariate investigated. State the planned analytic approach.

For the description of the studies included, we aim to analyse the following study characteristics to be considered for presentation and discussion:

- Number of studies included identifying barriers and facilitators for both treatments (IPT, CPT)
- Study populations
- Countries represented
- Context represented
- Data collection approaches applied
- Quality of the studies (summary of overall rating)

### 30. * Type and method of review.

Select the type of review, review method and health area from the lists below.

#### Type of review

Cost effectiveness
No

Diagnostic
No

Epidemiologic
No

Individual patient data (IPD) meta-analysis
No

Intervention
No

Meta-analysis
No

Methodology
No

Narrative synthesis
No

Network meta-analysis
No

Pre-clinical
No

Prevention
No

Prognostic
No

Prospective meta-analysis (PMA)
No

Review of reviews
No

Service delivery
No

Synthesis of qualitative studies
No

Systematic review
Yes

Other
Yes

Systematic review and Metasummary

### Health area of the review

Alcohol/substance misuse/abuse
No

Blood and immune system
No

Cancer
No

Cardiovascular
No

Care of the elderly
No

Child health
No

Complementary therapies
No

COVID-19
No

Crime and justice
No

Dental
No

Digestive system
No

Ear, nose and throat
No

Education

No

Endocrine and metabolic disorders
No

Eye disorders
No

General interest
No

Genetics
No

Health inequalities/health equity
No

Infections and infestations
Yes

International development
No

Mental health and behavioural conditions
No

Musculoskeletal
No

Neurological
No

Nursing
No

Obstetrics and gynaecology
No

Oral health
No

Palliative care
No

Perioperative care
No

Physiotherapy
No

Pregnancy and childbirth
No

Public health (including social determinants of health)
Yes

Rehabilitation
No

Respiratory disorders
No

Service delivery
Yes

Skin disorders
No

Social care
No

Surgery
No

Tropical Medicine
No

Urological
No

Wounds, injuries and accidents
No

Violence and abuse
No

### 31. Language.

Select each language individually to add it to the list below, use the bin icon to remove any added in error.
English

There is not an English language summary

### 32. * Country.

Select the country in which the review is being carried out. For multi-national collaborations select all the countries involved.

Portugal

### 33. Other registration details.

Name any other organisation where the systematic review title or protocol is registered (e.g. Campbell, or The Joanna Briggs Institute) together with any unique identification number assigned by them. If extracted data will be stored and made available through a repository such as the Systematic Review Data Repository (SRDR), details and a link should be included here. If none, leave blank.

### 34. Reference and/or URL for published protocol.

If the protocol for this review is published provide details (authors, title and journal details, preferably in Vancouver format)

Add web link to the published protocol.

Or, upload your published protocol here in pdf format. Note that the upload will be publicly accessible.

No I do not make this file publicly available until the review is complete

Please note that the information required in the PROSPERO registration form must be completed in full even if access to a protocol is given.

### 35. Dissemination plans.

Do you intend to publish the review on completion?

Yes

Give brief details of plans for communicating review findings.?

Publication in scientific journal

### 36. Keywords.

Give words or phrases that best describe the review. Separate keywords with a semicolon or new line. Keywords help PROSPERO users find your review (keywords do not appear in the public record but are included in searches). Be as specific and precise as possible. Avoid acronyms and abbreviations unless these are in wide use.

Cotrimoxazol, Isoniazid, HIV, TB, barriers, facilitators, implementation science, service delivery

### 37. Details of any existing review of the same topic by the same authors.

If you are registering an update of an existing review give details of the earlier versions and include a full bibliographic reference, if available.

### 38. * Current review status.

Update review status when the review is completed and when it is published.New registrations must be ongoing.
Please provide anticipated publication date

Review_Ongoing

### 39. Any additional information.

Provide any other information relevant to the registration of this review.

### 40. Details of final report/publication(s) or preprints if available.

Leave empty until publication details are available OR you have a link to a preprint. List authors, title and journal details preferably in Vancouver format.

Give the link to the published review or preprint.
