## Supplementary material for "Mixed methods systematic review and metasummary about barriers and facilitators for the implementation of cotrimoxazole and isoniazid – preventive therapies for people living with HIV": S4 Additional file. Database searches and results.

Additional file 4. Database searches and results.

| Database | Detailed Search expression | Results 8th of February 2018 | Results 4th of September 2020 |
|---|---|---|---|
| MEDLINE (via Pubmed) | #1 Cotrimoxazol* OR Co-trimoxazol* OR "Trimethoprim, Sulfamethoxazole Drug Combination"[Mesh].<br>#2 Isoniazid OR "Isoniazid"[Mesh].<br>#3 barrier* OR facilitat* OR challeng*.<br>#4 implementation OR provision<br>**(#1 OR #2) AND #3 AND #4**<br><br>(Cotrimoxazol* OR Co-trimoxazol* OR "Trimethoprim, Sulfamethoxazole Drug Combination"[Mesh] OR Isoniazid OR "Isoniazid"[Mesh]) AND (barrier* OR facilitat* OR challeng*) AND (implementation OR provision) | n= 95 | n = 126 |
| Web of Science (all databases) | #1 Cotrimoxazol* OR Co-trimoxazol* OR "Trimethoprim, Sulfamethoxazole Drug Combination".<br>#2 Isoniazid.<br>#3 barrier* OR facilitat* OR challeng*.<br>#4 implementation OR provision.<br>**#1 AND #2 AND #3 AND #4**<br><br>TOPIC: (Cotrimoxazol* OR Co-trimoxazol* OR "Trimethoprim, Sulfamethoxazole Drug Combination" OR Isoniazid) AND TOPIC: (barrier* OR facilitat* OR challeng*) AND TOPIC: (implementation OR provision)<br>Indexes=SCI-EXPANDED, SSCI, A&HCI, CPCI-S, CPCI-SSH, ESCI, CCR-EXPANDED, IC<br>Timespan=All years | n= 143 | n = 156 |
| Scopus | #1 Cotrimoxazol* OR Co-trimoxazol* OR "Trimethoprim, Sulfamethoxazole Drug Combination".<br>#2 Isoniazid.<br>#3 barrier* OR facilitat* OR challeng*.<br>#4 implementation OR provision.<br>**(#1 OR #2) AND #3 AND #4**<br><br>( TITLE-ABS-KEY ( ( cotrimoxazol* OR co-trimoxazol* OR "Trimethoprim,Sulfamethoxazole Drug Combination" OR isoniazid ) ) AND TITLE-ABS-KEY ( barrier* OR facilitat* OR challeng* ) AND TITLE-ABS-KEY ( implementation OR provision ) ) AND DOCTYPE ( ar ) | n = 146 | n = 200 |
