## Supplementary material for "Mixed methods systematic review and metasummary about barriers and facilitators for the implementation of cotrimoxazole and isoniazid – preventive therapies for people living with HIV": S5 Additional file. Mixed method quality appraisal tool (MMAT) and user guide.

### MIXED METHODS APPRAISAL TOOL (MMAT) VERSION 2018

#### User guide

Prepared by
Quan Nha HONG[a], Pierre PLUYE[a,], Sergi FÀBREGUES[b], Gillian BARTLETT[a], Felicity BOARDMAN[c], Margaret CARGO[d], Pierre DAGENAIS[e], Marie-Pierre GAGNON[f], Frances GRIFFITHS[c], Belinda NICOLAU[a], Alicia O'CATHAIN[g], Marie-Claude ROUSSEAU[h], & Isabelle VEDEL[a]

[a]McGill University, Montréal, Canada; [b]Universitat Oberta de Catalunya, Barcelona, Spain; [c]University of Warwick, Coventry, England; [d]University of Canberra, Canberra, Australia; [e]Université de Sherbrooke, Sherbrooke, Canada; [f]Université Laval, Québec, Canada; [g]University of Sheffield, Sheffield, England; [h]Institut Armand-Frappier Research Centre, Laval, Canada

McGill
Department of Family Medicine
Département de médecine de famille
Academic excellence and innovation in care, teaching and research
Innovation et excellence académique dans les soins, l'enseignement et la recherche

Last update: August 1st, 2018

#### What is the MMAT?

The MMAT is a critical appraisal tool that is designed for the appraisal stage of systematic mixed studies reviews, i.e., reviews that include qualitative, quantitative and mixed methods studies. It permits to appraise the methodological quality of five categories to studies: qualitative research, randomized controlled trials, non-randomized studies, quantitative descriptive studies, and mixed methods studies.

#### How was the MMAT developed?

The MMAT was developed in 2006 (Pluye et al., 2009a) and was revised in 2011 (Pace et al., 2012). The present version 2018 was developed on the basis of findings from a literature review of critical appraisal tools, interviews with MMAT users, and an e-Delphi study with international experts (Hong, 2018). The MMAT developers are continuously seeking for improvement and testing of this tool. Users' feedback is always appreciated.

#### What the MMAT can be used for?

The MMAT can be used to appraise the quality of empirical studies, i.e., primary research based on experiment, observation or simulation (Abbott, 1998; Porta et al., 2014). It cannot be used for non-empirical papers such as review and theoretical papers. Also, the MMAT allows the appraisal of most common types of study methodologies and designs. However, some specific designs such as economic and diagnostic accuracy studies cannot be assessed with the MMAT. Other critical appraisal tools might be relevant for these designs.

#### What are the requirements?

Because critical appraisal is about judgment making, it is advised to have at least two reviewers independently involved in the appraisal process. Also, using the MMAT requires experience or training in these domains. For instance, MMAT users may be helped by a colleague with specific expertise when needed.

#### How to use the MMAT?

This document comprises two parts: checklist (Part I) and explanation of the criteria (Part II).

1. Respond to the two screening questions. Responding 'No' or 'Can't tell' to one or both questions might indicate that the paper is not an empirical study, and thus cannot be appraised using the MMAT. MMAT users might decide not to use these questions, especially if the selection criteria of their review are limited to empirical studies.
2. For each included study, choose the appropriate category of studies to appraise. Look at the description of the methods used in the included studies. If needed, use the algorithm at the end of this document.
3. Rate the criteria of the chosen category. For example, if the paper is a qualitative study, only rate the five criteria in the qualitative category. The 'Can't tell' response category means that the paper do not report appropriate information to answer 'Yes' or 'No', or that report unclear information related to the criterion. Rating 'Can't tell' could lead to look for companion papers, or contact authors to ask more information or clarification when needed. In Part II of this document, indicators are added for some criteria. The list is not exhaustive and not all indicators are necessary. You should agree among your team which ones are important to consider for your field and apply them uniformly across all included studies from the same category.

#### How to score?

It is discouraged to calculate an overall score from the ratings of each criterion. Instead, it is advised to provide a more detailed presentation of the ratings of each criterion to better inform the quality of the included studies. This may lead to perform a sensitivity analysis (i.e., to consider the quality of studies by contrasting their results). Excluding studies with low methodological quality is usually discouraged.

#### How to cite this document?

Hong QN, Pluye P, Fàbregues S, Bartlett G, Boardman F, Cargo M, Dagenais P, Gagnon M-P, Griffiths F, Nicolau B, O'Cathain A, Rousseau M-C, Vedel I. Mixed Methods Appraisal Tool (MMAT), version 2018. Registration of Copyright (#1148552), Canadian Intellectual Property Office, Industry Canada.

**Part I: Mixed Methods Appraisal Tool (MMAT), version 2018**

| Category of study designs | Methodological quality criteria | Responses | | | |
|---|---|---|---|---|---|
| | | Yes | No | Can't tell | Comments |
| Screening questions (for all types) | S1. Are there clear research questions? | | | | |
| | S2. Do the collected data allow to address the research questions? | | | | |
| | *Further appraisal may not be feasible or appropriate when the answer is 'No' or 'Can't tell' to one or both screening questions.* | | | | |
| 1. Qualitative | 1.1. Is the qualitative approach appropriate to answer the research question? | | | | |
| | 1.2. Are the qualitative data collection methods adequate to address the research question? | | | | |
| | 1.3. Are the findings adequately derived from the data? | | | | |
| | 1.4. Is the interpretation of results sufficiently substantiated by data? | | | | |
| | 1.5. Is there coherence between qualitative data sources, collection, analysis and interpretation? | | | | |
| 2. Quantitative randomized controlled trials | 2.1. Is randomization appropriately performed? | | | | |
| | 2.2. Are the groups comparable at baseline? | | | | |
| | 2.3. Are there complete outcome data? | | | | |
| | 2.4. Are outcome assessors blinded to the intervention provided? | | | | |
| | 2.5 Did the participants adhere to the assigned intervention? | | | | |
| 3. Quantitative non-randomized | 3.1. Are the participants representative of the target population? | | | | |
| | 3.2. Are measurements appropriate regarding both the outcome and intervention (or exposure)? | | | | |
| | 3.3. Are there complete outcome data? | | | | |
| | 3.4. Are the confounders accounted for in the design and analysis? | | | | |
| | 3.5. During the study period, is the intervention administered (or exposure occurred) as intended? | | | | |
| 4. Quantitative descriptive | 4.1. Is the sampling strategy relevant to address the research question? | | | | |
| | 4.2. Is the sample representative of the target population? | | | | |
| | 4.3. Are the measurements appropriate? | | | | |
| | 4.4. Is the risk of nonresponse bias low? | | | | |
| | 4.5. Is the statistical analysis appropriate to answer the research question? | | | | |
| 5. Mixed methods | 5.1. Is there an adequate rationale for using a mixed methods design to address the research question? | | | | |
| | 5.2. Are the different components of the study effectively integrated to answer the research question? | | | | |
| | 5.3. Are the outputs of the integration of qualitative and quantitative components adequately interpreted? | | | | |
| | 5.4. Are divergences and inconsistencies between quantitative and qualitative results adequately addressed? | | | | |
| | 5.5. Do the different components of the study adhere to the quality criteria of each tradition of the methods involved? | | | | |

#### Part II: Explanations

| 1. Qualitative studies | Methodological quality criteria |
|---|---|
| "Qualitative research is an approach for exploring and understanding the meaning individuals or groups ascribe to a social or human problem" (Creswell, 2013b, p. 3).<br><br>Common qualitative research approaches include (this list if not exhaustive):<br><br>**Ethnography**<br>The aim of the study is to describe and interpret the shared cultural behaviour of a group of individuals.<br><br>**Phenomenology**<br>The study focuses on the subjective experiences and interpretations of a phenomenon encountered by individuals.<br><br>**Narrative research**<br>The study analyzes life experiences of an individual or a group.<br><br>**Grounded theory**<br>Generation of theory from data in the process of conducting research (data collection occurs first).<br><br>**Case study**<br>In-depth exploration and/or explanation of issues intrinsic to a particular case. A case can be anything from a decision-making process, to a person, an organization, or a country.<br><br>**Qualitative description**<br>There is no specific methodology, but a qualitative data collection and analysis, e.g., in-depth interviews or focus groups, and hybrid thematic analysis (inductive and deductive).<br><br>Key references: Creswell (2013a); Sandelowski (2010); Schwandt (2015) | 1.1. Is the qualitative approach appropriate to answer the research question?<br><br>Explanations<br>The qualitative approach used in a study (see non-exhaustive list on the left side of this table) should be appropriate for the research question and problem. For example, the use of a grounded theory approach should address the development of a theory and ethnography should study human cultures and societies.<br><br>This criterion was considered important to add in the MMAT since there is only one category of criteria for qualitative studies (compared to three for quantitative studies).<br><br>1.2. Are the qualitative data collection methods adequate to address the research question?<br><br>Explanations<br>This criterion is related to data collection method, including data sources (e.g., archives, documents), used to address the research question. To judge this criterion, consider whether the method of data collection (e.g., in depth interviews and/or group interviews, and/or observations) and the form of the data (e.g., tape recording, video material, diary, photo, and/or field notes) are adequate. Also, clear justifications are needed when data collection methods are modified during the study.<br><br>1.3. Are the findings adequately derived from the data?<br><br>Explanations<br>This criterion is related to the data analysis used. Several data analysis methods have been developed and their use depends on the research question and qualitative approach. For example, open, axial and selective coding is often associated with grounded theory, and within- and cross-case analysis is often seen in case study.<br><br>1.4. Is the interpretation of results sufficiently substantiated by data?<br><br>Explanations<br>The interpretation of results should be supported by the data collected. For example, the quotes provided to justify the themes should be adequate.<br><br>1.5. Is there coherence between qualitative data sources, collection, analysis and interpretation?<br><br>Explanations<br>There should be clear links between data sources, collection, analysis and interpretation. |

| 2. Quantitative randomized controlled trials | Methodological quality criteria |
|---|---|
| **Randomized controlled clinical trial**: A clinical study in which individual participants are allocated to intervention or control groups by randomization (intervention assigned by researchers).<br><br>Key references: Higgins and Green (2008); Higgins et al. (2016); Oxford Centre for Evidence-based Medicine (2016); Porta et al. (2014) | 2.1. Is randomization appropriately performed?<br><br>Explanations<br>In a randomized controlled trial, the allocation of a participant (or a data collection unit, e.g., a school) into the intervention or control group is based solely on chance. Researchers should describe how the randomization schedule was generated. A simple statement such as 'we randomly allocated' or 'using a randomized design' is insufficient to judge if randomization was appropriately performed. Also, assignment that is predictable such as using odd and even record numbers or dates is not appropriate. At minimum, a simple allocation (or unrestricted allocation) should be performed by following a predetermined plan/sequence. It is usually achieved by referring to a published list of random numbers, or to a list of random assignments generated by a computer. Also, restricted allocation can be performed such as blocked randomization (to ensure particular allocation ratios to the intervention groups), stratified randomization (randomization performed separately within strata), or minimization (to make small groups closely similar with respect to several characteristics). Another important characteristic to judge if randomization was appropriately performed is allocation concealment that protects assignment sequence until allocation. Researchers and participants should be unaware of the assignment sequence up to the point of allocation. Several strategies can be used to ensure allocation concealment such relying on a central randomization by a third party, or the use of sequentially numbered, opaque, sealed envelopes (Higgins et al., 2016). |
| | 2.2. Are the groups comparable at baseline?<br><br>Explanations<br>Baseline imbalance between groups suggests that there are problems with the randomization. Indicators from baseline imbalance include: "(1) unusually large differences between intervention group sizes; (2) a substantial excess in statistically significant differences in baseline characteristics than would be expected by chance alone; (3) imbalance in key prognostic factors (or baseline measures of outcome variables) that are unlikely to be due to chance; (4) excessive similarity in baseline characteristics that is not compatible with chance; (5) surprising absence of one or more key characteristics that would be expected to be reported" (Higgins et al., 2016, p. 10). |
| | 2.3. Are there complete outcome data?<br><br>Explanations<br>Almost all the participants contributed to almost all measures. There is no absolute and standard cut-off value for acceptable complete outcome data. Agree among your team what is considered complete outcome data in your field and apply this uniformly across all the included studies. For instance, in the literature, acceptable complete data value ranged from 80% (Thomas et al., 2004; Zaza et al., 2000) to 95% (Higgins et al., 2016). Similarly, different acceptable withdrawal/dropouts rates have been suggested: 5% (de Vet et al., 1997; MacLehose et al., 2000), 20% (Sindhu et al., 1997; Van Tulder et al., 2003) and 30% for a follow-up of more than one year (Viswanathan and Berkman, 2012). |
| | 2.4. Are outcome assessors blinded to the intervention provided?<br><br>Explanations<br>Outcome assessors should be unaware of who is receiving which interventions. The assessors can be the participants if using participant reported outcome (e.g., pain), the intervention provider (e.g., clinical exam), or other persons not involved in the intervention (Higgins et al., 2016). |
| | 2.5 Did the participants adhere to the assigned intervention?<br><br>Explanations<br>To judge this criterion, consider the proportion of participants who continued with their assigned intervention throughout follow-up. "Lack of adherence includes imperfect compliance, cessation of intervention, crossovers to the comparator intervention and switches to another active intervention." (Higgins et al., 2016, p. 25). |

<table>
<tr><th>3. Quantitative non-randomized studies</th><th>Methodological quality criteria</th></tr>
<tr><td rowspan="5">Non-randomized studies are defined as any quantitative studies estimating the effectiveness of an intervention or studying other exposures that do not use randomization to allocate units to comparison groups (Higgins and Green, 2008).<br><br>Common designs include (this list if not exhaustive):<br><br>Non-randomized controlled trials<br>The intervention is assigned by researchers, but there is no randomization, e.g., a pseudo-randomization. A non-random method of allocation is not reliable in producing alone similar groups.<br><br>Cohort study<br>Subsets of a defined population are assessed as exposed, not exposed, or exposed at different degrees to factors of interest. Participants are followed over time to determine if an outcome occurs (prospective longitudinal).<br><br>Case-control study<br>Cases, e.g., patients, associated with a certain outcome are selected, alongside a corresponding group of controls. Data is collected on whether cases and controls were exposed to the factor under study (retrospective).<br><br>Cross-sectional analytic study<br>At one particular time, the relationship between health-related characteristics (outcome) and other factors (intervention/exposure) is examined. E.g., the frequency of outcomes is compared in different population subgroups according to the presence/absence (or level) of the intervention/exposure.<br><br>Key references for non-randomized studies: Higgins and Green (2008); Porta et al. (2014); Sterne et al. (2016); Wells et al. (2000)</td><td>3.1. Are the participants representative of the target population?<br><br>Explanations<br>Indicators of representativeness include: clear description of the target population and of the sample (inclusion and exclusion criteria), reasons why certain eligible individuals chose not to participate, and any attempts to achieve a sample of participants that represents the target population.</td></tr>
<tr><td>3.2. Are measurements appropriate regarding both the outcome and intervention (or exposure)?<br><br>Explanations<br>Indicators of appropriate measurements include: the variables are clearly defined and accurately measured; the measurements are justified and appropriate for answering the research question; the measurements reflect what they are supposed to measure; validated and reliability tested measures of the intervention/exposure and outcome of interest are used, or variables are measured using ‘gold standard’.</td></tr>
<tr><td>3.3. Are there complete outcome data?<br><br>Explanations<br>Almost all the participants contributed to almost all measures. There is no absolute and standard cut-off value for acceptable complete outcome data. Agree among your team what is considered complete outcome data in your field (and based on the targeted journal) and apply this uniformly across all the included studies. For example, in the literature, acceptable complete data value ranged from 80% (Thomas et al., 2004; Zaza et al., 2000) to 95% (Higgins et al., 2016). Similarly, different acceptable withdrawal/dropouts rates have been suggested: 5% (de Vet et al., 1997; MacLehose et al., 2000), 20% (Sindhu et al., 1997; Van Tulder et al., 2003) and 30% for follow-up of more than one year (Viswanathan and Berkman, 2012).</td></tr>
<tr><td>3.4. Are the confounders accounted for in the design and analysis?<br><br>Explanations<br>Confounders are factors that predict both the outcome of interest and the intervention received/exposure at baseline. They can distort the interpretation of findings and need to be considered in the design and analysis of a non-randomized study. Confounding bias is low if there is no confounding expected, or appropriate methods to control for confounders are used (such as stratification, regression, matching, standardization, and inverse probability weighting).</td></tr>
<tr><td>3.5 During the study period, is the intervention administered (or exposure occurred) as intended?<br><br>Explanations<br>For intervention studies, consider whether the participants were treated in a way that is consistent with the planned intervention. Since the intervention is assigned by researchers, consider whether there was a presence of contamination (e.g., the control group may be indirectly exposed to the intervention) or whether unplanned co-interventions were present in one group (Sterne et al., 2016).<br><br>For observational studies, consider whether changes occurred in the exposure status among the participants. If yes, check if these changes are likely to influence the outcome of interest, were adjusted for, or whether unplanned co-exposures were present in one group (Morgan et al., 2017).</td></tr>
</table>

<table>
<tr><th>4. Quantitative descriptive studies</th><th>Methodological quality criteria</th></tr>
<tr><td rowspan="5">Quantitative descriptive studies are “concerned with and designed only to describe the existing distribution of variables without much regard to causal relationships or other hypotheses” (Porta et al., 2014, p. 72). They are used to monitoring the population, planning, and generating hypothesis (Grimes and Schulz, 2002).<br><br>Common designs include the following single-group studies (this list if not exhaustive):<br><br>Incidence or prevalence study without comparison group<br>In a defined population at one particular time, what is happening in a population, e.g., frequencies of factors (importance of problems), is described (portrayed).<br><br>Survey<br>“Research method by which information is gathered by asking people questions on a specific topic and the data collection procedure is standardized and well defined.” (Bennett et al., 2011, p. 3).<br><br>Case series<br>A collection of individuals with similar characteristics are used to describe an outcome.<br><br>Case report<br>An individual or a group with a unique/unusual outcome is described in detail.<br><br>Key references: Critical Appraisal Skills Programme (2017); Draugalis et al. (2008)</td><td>4.1. Is the sampling strategy relevant to address the research question?<br><br>Explanations<br>Sampling strategy refers to the way the sample was selected. There are two main categories of sampling strategies: probability sampling (involve random selection) and non-probability sampling. Depending on the research question, probability sampling might be preferable. Non-probability sampling does not provide equal chance of being selected. To judge this criterion, consider whether the source of sample is relevant to the target population; a clear justification of the sample frame used is provided; or the sampling procedure is adequate.</td></tr>
<tr><td>4.2. Is the sample representative of the target population?<br><br>Explanations<br>There should be a match between respondents and the target population. Indicators of representativeness include: clear description of the target population and of the sample (such as respective sizes and inclusion and exclusion criteria), reasons why certain eligible individuals chose not to participate, and any attempts to achieve a sample of participants that represents the target population.</td></tr>
<tr><td>4.3. Are the measurements appropriate?<br><br>Explanations<br>Indicators of appropriate measurements include: the variables are clearly defined and accurately measured, the measurements are justified and appropriate for answering the research question; the measurements reflect what they are supposed to measure; validated and reliability tested measures of the outcome of interest are used, variables are measured using ‘gold standard’, or questionnaires are pre-tested prior to data collection.</td></tr>
<tr><td>4.4. Is the risk of nonresponse bias low?<br><br>Explanations<br>Nonresponse bias consists of “an error of nonobservation reflecting an unsuccessful attempt to obtain the desired information from an eligible unit.” (Federal Committee on Statistical Methodology, 2001, p. 6). To judge this criterion, consider whether the respondents and non-respondents are different on the variable of interest. This information might not always be reported in a paper. Some indicators of low nonresponse bias can be considered such as a low nonresponse rate, reasons for nonresponse (e.g., noncontacts vs. refusals), and statistical compensation for nonresponse (e.g., imputation).<br><br>The nonresponse bias is might not be pertinent for case series and case report. This criterion could be adapted. For instance, complete data on the cases might be important to consider in these designs.</td></tr>
<tr><td>4.5. Is the statistical analysis appropriate to answer the research question?<br><br>Explanations<br>The statistical analyses used should be clearly stated and justified in order to judge if they are appropriate for the design and research question, and if any problems with data analysis limited the interpretation of the results.</td></tr>
</table>

| 5. Mixed methods studies | Methodological quality criteria |
|---|---|
| Mixed methods (MM) research involves combining qualitative (QUAL) and quantitative (QUAN) methods. In this tool, to be considered MM, studies have to meet the following criteria (Creswell and Plano Clark, 2017): (a) at least one QUAL method and one QUAN method are combined; (b) each method is used rigorously in accordance to the generally accepted criteria in the area (or tradition) of research invoked; and (c) the combination of the methods is carried out at the minimum through a MM design (defined *a priori*, or emerging) and the integration of the QUAL and QUAN phases, results, and data.<br><br>Common designs include (this list if not exhaustive):<br><br>**Convergent design**<br>The QUAL and QUAN components are usually (but not necessarily) concomitant. The purpose is to examine the same phenomenon by interpreting QUAL and QUAN results (bringing data analysis together at the interpretation stage), or by integrating QUAL and QUAN datasets (e.g., data on same cases), or by transforming data (e.g., quantization of qualitative data).<br><br>**Sequential explanatory design**<br>Results of the phase 1 - QUAN component inform the phase 2 - QUAL component. The purpose is to explain QUAN results using QUAL findings. E.g., the QUAN results guide the selection of QUAL data sources and data collection, and the QUAL findings contribute to the interpretation of QUAN results.<br><br>**Sequential exploratory design**<br>Results of the phase 1 - QUAL component inform the phase 2 - QUAN component. The purpose is to explore, develop and test an instrument (or taxonomy), or a conceptual framework (or theoretical model). E.g., the QUAL findings inform the QUAN data collection, and the QUAN results allow a statistical generalization of the QUAL findings.<br><br>Key references: Creswell et al. (2011); Creswell and Plano Clark, (2017); O'Cathain (2010) | 5.1. Is there an adequate rationale for using a mixed methods design to address the research question?<br><br>Explanations<br>The reasons for conducting a mixed methods study should be clearly explained. Several reasons can be invoked such as to enhance or build upon qualitative findings with quantitative results and vice versa; to provide a comprehensive and complete understanding of a phenomenon or to develop and test instruments (Bryman, 2006). |
| | 5.2. Are the different components of the study effectively integrated to answer the research question?<br><br>Explanations<br>Integration is a core component of mixed methods research and is defined as the “explicit interrelating of the quantitative and qualitative component in a mixed methods study” (Plano Clark and Ivankova, 2015, p. 40). Look for information on how qualitative and quantitative phases, results, and data were integrated (Pluye et al., 2018). For instance, how data gathered by both research methods was brought together to form a complete picture (e.g., joint displays) and when integration occurred (e.g., during the data collection-analysis or/and during the interpretation of qualitative and quantitative results). |
| | 5.3. Are the outputs of the integration of qualitative and quantitative components adequately interpreted?<br><br>Explanations<br>This criterion is related to meta-inference, which is defined as the overall interpretations derived from integrating qualitative and quantitative findings (Teddlie and Tashakkori, 2009). Meta-inference occurs during the interpretation of the findings from the integration of the qualitative and quantitative components, and shows the added value of conducting a mixed methods study rather than having two separate studies. |
| | 5.4. Are divergences and inconsistencies between quantitative and qualitative results adequately addressed?<br><br>Explanations<br>When integrating the findings from the qualitative and quantitative components, divergences and inconsistencies (also called conflicts, contradictions, discordances, discrepancies, and dissonances) can be found. It is not sufficient to only report the divergences; they need to be explained. Different strategies to address the divergences have been suggested such as reconciliation, initiation, bracketing and exclusion (Pluye et al., 2009b). Rate this criterion ‘Yes’ if there is no divergence. |
| | 5.5. Do the different components of the study adhere to the quality criteria of each tradition of the methods involved?<br><br>Explanations<br>The quality of the qualitative and quantitative components should be individually appraised to ensure that no important threats to trustworthiness are present. To appraise 5.5, use criteria for the qualitative component (1.1 to 1.5), and the appropriate criteria for the quantitative component (2.1 to 2.5, or 3.1 to 3.5, or 4.1 to 4.5). The quality of both components should be high for the mixed methods study to be considered of good quality. The premise is that the overall quality of a mixed methods study cannot exceed the quality of its weakest component. For example, if the quantitative component is rated high quality and the qualitative component is rated low quality, the overall rating for this criterion will be of low quality. |

**Algorithm for selecting the study categories to rate in the MMAT***

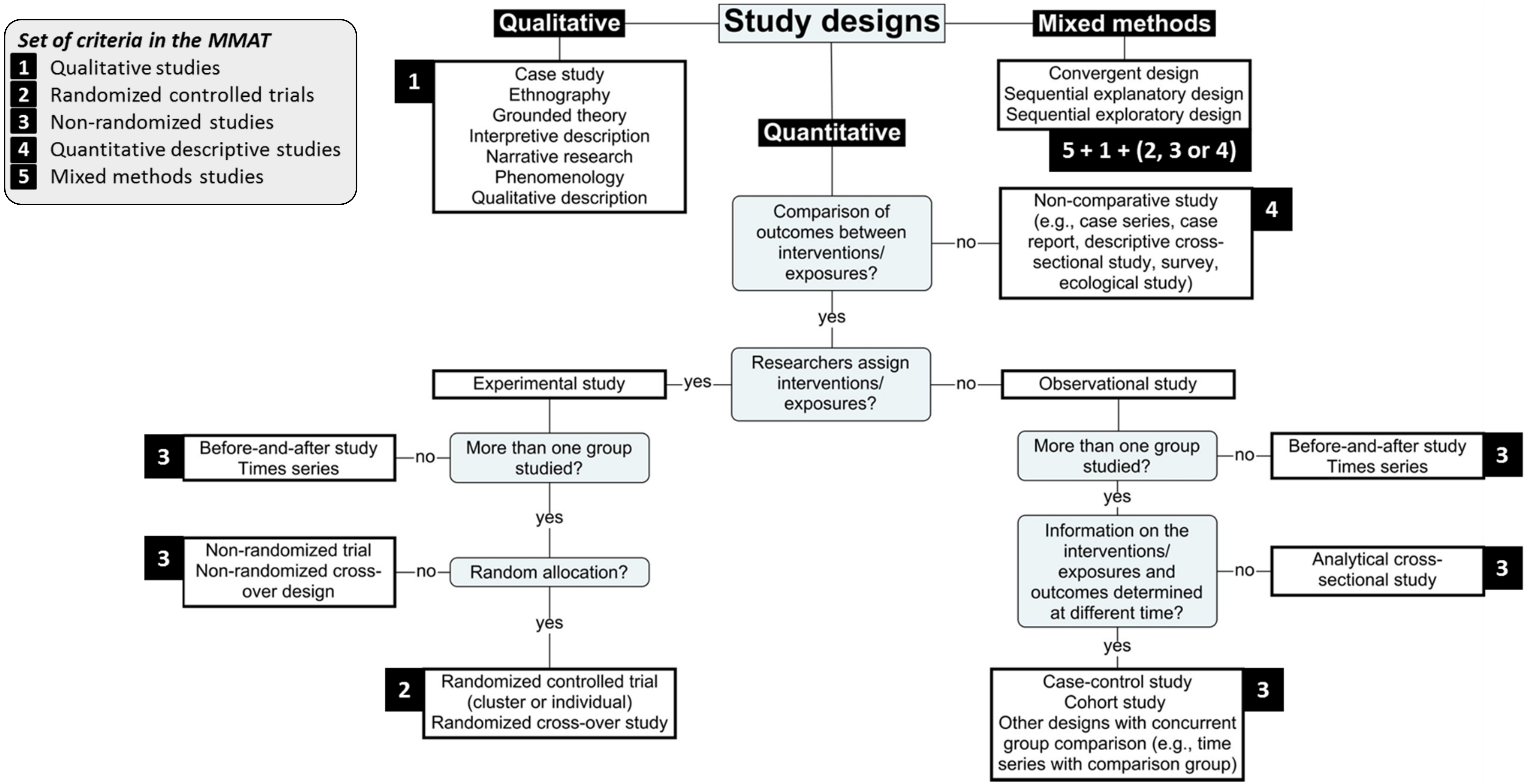


.

*Adapted from National Institute for Health Care Excellence. (2012). *Methods for the development of nice public health guidance*. London: National Institute for Health and Care Excellence; and Scottish Intercollegiate Guidelines Network. (2017). *Algorithm for classifying study design for questions of effectiveness*. Retrieved December 1, 2017, from http://www.sign.ac.uk/assets/study_design.pdf.
