## Supplementary material for "Mixed methods systematic review and metasummary about barriers and facilitators for the implementation of cotrimoxazole and isoniazid – preventive therapies for people living with HIV": S6 Additional file. Detailed description of studies included in this review.

Additional file 6. Detailed description of studies included in this review.

**Table 1. Description of studies included on cotrimoxazole preventive therapy (n= 14).** *incl. papers of Okot-Chono (2009), Mugomeri (2018) which refer to both preventive therapies.

| First Author (Year) | Country | Context | Study type | Study subject(s) of interest | | Study aims | Data collection approach |
|---|---|---|---|---|---|---|---|
| Ansa (2014) | Ghana | urban | Comparative research | Facilities | (3 hospitals with different delivery models including one-stop-shop, partial integration and referral model) | To compare the impact of increasing integration on TB/HIV service delivery including HIV screening, ART and CPT. | Review of 590 TB patients’ health records |
| Chan (2014) | Malawi<br>Uganda<br>Zimbabwe | rural, semi-urban, urban | Multi-country comparative study | Facilities | (81 health facilities, including 53 primary, 25 secondary and 3 tertiary care facilities) | To describe and compare national and inter-country delivery of training, clinical care, use of laboratories, and monitoring of ART roll-out. | Interviewer administered mixed methods questionnaire for nurses, doctors, and laboratory personnel,<br>Review of ART registers, health records, pharmacy records |
| Chang (2015) | Uganda | - | Randomised trial | Patients | (442 pre-ART HIV+ adults, including 221 in the intervention group and 221 in the control group) | To assess the effect of peer support on HIV clinic attendance, preventive care use including CPT, risky sexual behaviour and ART initiation. | Survey-based outcome assessment at baseline and after one-year follow-up |
| Horwood (2010) | South Africa | peri-urban, rural | Cross-sectional descriptive study/ evaluation | Facilities | (6 hospital-based post-natal wards and 27 immunization units of primary health care facilities) | To evaluate the implementation of prevention of mother to child transmission of HIV services and the integration of PMTCT services with routine maternal and child health services.<br>To compare health interventions provided at both types of health care units according to patients’ health records with mothers reporting. | Review of 872 HIV+ mothers’ health records,<br>Survey-based interviews with<br>872 mothers regarding the services received,<br>26 nurses, and 27 lay counsellors regarding their work activities |
| Kamuhabwa (2015) | Tanzania | - | Retrospective descriptive study | Facilities | (PMTCT units at 4 public hospitals) | To assess CPT delivery in PMTCT units (including initiation, doses prescribed, duration, drug tolerance, adherence). | Review of notes of 498 infants born to HIV+ mothers recorded in PMTCT register books, |
| | | | | Caregivers<br>Providers | (321 parents/ guardians)<br>(126 healthcare workers) | To identify challenges in the provision of CPT among children born to HIV-infected mothers. | Interviewer administered questionnaire with structured open and closed questions |
| Kamuhabwa (2016) | Tanzania | urban | Descriptive cross-sectional study | Patients | (353 HIV+ pregnant women) | To assess the level of implementation of CPT for malaria prevention among HIV+ pregnant women. | Review of patient health records, |
| | | | | Patients | (353 HIV+ pregnant women) | To assess self-reported adherence and reasons for non-adherence. | Interviews using a structured open and closed questions questionnaire, |
| | | | | Providers | (26 health providers) | | |
| | | | | Providers | (18 health providers) | To assess the level of knowledge of health providers regarding the new CPT policy. | Self-administered questionnaires, |
| | | | | Facilities | (3 public health facilities, incl. two hospitals and one health centre) | To identify challenges encountered regarding CPT provision at the health facility level. | Focus Group Discussion, |
| | | | | Facilities | | To identify challenges encountered regarding the new CPT policy at the health facility level. | Facility assessment, |
| Louwagie (2012) | South Africa | - | Historical cohort study | Facilities | (46 health facilities with different delivery models incl. 4 semi-integrated and 42 separate facilities) | To compare access to HIV-related care, including CPT provision between non-integrated (separate) facilities and semi-integrated facilities. | Review of TB Registers, TB treatment records, electronic & paper based ART patient records of 636 TB patients from semi-integrated, and 1297 TB patients from separate facilities |
| *Mugomeri (2018) | Lesotho | - | Qualitative study | Providers<br>Stake-holders | (42 public healthcare workers, 2 Ministry of Health key informants, 6 representatives of partner organizations) | To establish barriers to the implementation of IPT within the health system context of Lesotho. | Individual semi-structured interviews |

Additional file 6. Detailed description of studies included in this review.

**Table 1. (…continued)**

| First Author (Year) | Country | Context | Study type | Study subject(s) of interest | | Study aims | Data collection approach |
|---|---|---|---|---|---|---|---|
| Luyirika (2013) | Uganda | - | Retrospective case study | Facilities | (public and private health facilities including 10 community clinics supported by NGO and 9 or 10? NGO partner health facilities) | To examine retrospectively how family-based care approach (which aims to integrate paediatric HIV care into adult HIV and maternal and child health services) was utilized to integrate HIV services. | Key documents review (i.e. programme strategies, guidelines, national HIV Policies, memos on CPT scale-up), |
| | | | | | | To examine operational successes and challenges across sites and identify lessons and recommendations for a family-centred approach. | Key informant interviews with programme management, and patients' families, |
| | | | | | | To assess changes in service uptake since the introduction of the model. | Record review using hospital management information system |
| Mwambete (2013) | Tanzania | urban | Serial clinical and cross-sectional resistance study | Patients | (188 HIV+ adults eligible for CPT ) | To investigate incidences of faecal *E. coli* resistance to CTZ in HIV+ patients. | Stool samples for resistance profiling at the baseline visit and 4 follow-up visits, |
| | | | | | | To compare and determine changes in CTZ resistance over time.<br>To explore factors that may have contributed to resistance to CTZ and other commonly used antibiotics. | Patient self-reported adherence to CPT during each follow-up visit, questionnaire for initial collection of socio-demographic data |
| Naikoba (2017) | Uganda | rural | Cluster-randomized trial | Providers | (40 mid-level providers including 20 providers in the 5 intervention facilities and 20 providers in the 5 control facilities) | To assess the effect of one-on-one mentorship for task shifting on clinical knowledge, competence scores and facility performance. | Practical Assessment (using Case-Scenarios and clinical observation to determine mean scores for clinical knowledge and competence, |
| | | | | Facilities | (10 facilities; incl. 5 intervention, 5 control facilities) | To assess health facility performance (i.e. prescription of CPT) in both arms. | Review of electronic HIV care & treatment database and TB registers |
| *Okot-Chono (2009) | Uganda | peri-urban, rural | Record review, qualitative study | Districts | (5 districts) | To assess the proportions of patients utilising TB-HIV collaborative services. | Review of TB registers to assess the TB-HIV services utilization (incl. CPT) among 333 TB patients, |
| | | | | Patients<br>Providers<br>CM | (125 TB patients)<br>(65 health providers)<br>(70 community members) | To assess respondents knowledge, attitudes, practices and beliefs about TB-HIV collaborative services. | Standardised tools applied for 26 Focus Group Discussions, and 34 Key informant interviews,<br><br>28 In-depth interviews |
| | | | | Providers | (? facility in-charges, ? focal persons for TB/HIV, ? district health officers, ? expert patients) | To identify reasons explaining the level of implementation of TB-HIV collaborative services. | |
| Okwera (2015) | Uganda | - | Qualitative study | Patients | (n= 30 HIV+ pulmonary TB suspects) | To assess knowledge, attitudes, and practices of Cotrimoxazole use among HIV infected adults evaluated for TB. | 5 Focus Group Discussions |
| Sibanda (2015) | Zimbabwe | - | Qualitative study | Caregivers | (n= 20 HIV+ mothers whose babies had been initiated CPT) | To explore women´s ability to maintain their babies on CPT after they had been initiated at age six weeks. | In-depth interviews |

Additional file 6. Detailed description of studies included in this review.

**Table 2. Description of studies included on isoniazid preventive therapy (n= 28).** *incl. papers of Okot-Chono (2009), Mugomeri (2018) which refer to both preventive therapies.

| First Author (Year) | Country | Context | Study type | Study subject(s) of interest | | Study aims | Data collection approach |
|---|---|---|---|---|---|---|---|
| Adepoju (2020) | Nigeria | - | Retrospective cohort study | Patients | (1134 HIV+ adult attendees of 6 governmental hospitals enrolled in HIV care on IPT) | To assess IPT completion rate for 6 months IPT among PLHIV.<br>To determine predictive factors for IPT completion. | Review of monthly prescription refill records in ART care card, IPT register, and pharmacy order form |
| Aisu (1995) | Uganda | urban, sub-urban | Operational Assessment | Patients | (520 HIV+ attendees of a voluntary counselling and testing centre, eligible for IPT) | To assess compliance to 6 months IPT among HIV+ individuals with latent TB. | Review of standardized forms (compliance based on attendance to follow-up visits for pill collection and pill count at each follow-up visit), |
| | | | | Patients | (198 non-compliant patients) | To determine the reasons for missing appointments, to assess toxicity and determine the reasons for missing the follow-up appointment. | Interviews, |
| | | | | Facility | (1 Voluntary HIV counselling and testing centre) | To assess the incremental unit cost at each step from counselling, testing and ending with completion of IPT course. | Review of expenditure reports, project budgets, UN essential drugs price list |
| Catalani (2014) | Kenya | rural, urban | Mixed methods assessment | Providers Stake-holders | (24 stakeholders with various background) | To explore the social context of HIV and TB care, and strategies to influence providers care practice. | Key informant interviews using an unstructured interview guide |
| | | | | Facilities | (9 sites) | To explore the existing electronic medical record system and other existing recording systems. | Qualitative field notes of site observations using a semi-structured observation guide |
| | | | | Providers | (9 or 10? HIV clinicians) | To evaluate a new clinical decision support system for integrating TB and HIV care and to assess clinicians perceptions of the helpfulness and accuracy of the TB reminder messages. | Mixed methods usability surveys regarding in-context usability of the support system, In-depth interviews |
| Durovni (2010) | Brazil | - | Preliminary findings of phased cluster-randomised trial and qualitative study | Patients | (1670 HIV+ THRio trial participants of in 29 facilities) | To assess side effects reported during the ongoing trial and to compare IPT completion prior and post training. | Analysis of Trial data set (Trial aimed at testing the effect of provider training & new IPT policy implementation on the incidence of active TB in the HIV+ clinic population), |
| | | | | Providers | (10 HIV and TB clinicians) | To compare preintervention and post-intervention time to TST and time to IPT. | Interviews, |
| | | | | Stake-holders | (6 administrators, 6 nurses) | To explore health providers motivations and practices concerning IPT. | 2 Focus Group Discussions |
| Faust (2020) | Ethiopia, Nigeria, India, Angola, Brazil, China, DRC, Indonesia, Kenya, Lesotho, Liberia, Mozambique, Myanmar, South Africa, Tanzania, Thailand, Zambia, Zimbabwe | - | Survey | Countries | (18 high TB/HIV burden countries included in this systematic review) | To identify challenges experienced in TB HBCs with regard to the implementation of LTBI policies and tools. | 19-questions survey with open-ended and predefined response options via email |

Additional file 6. Detailed description of studies included in this review.

**Table 2. (…continued)**

| First Author (Year) | Country | Context | Study type | Study subject(s) of interest | | Study aims | Data collection approach |
|---|---|---|---|---|---|---|---|
| Gust (2011) | Botswana | - | Sub-study of the Botswana IPT prevention trial | Patients<br><br>Patients | (42 HIV+ trial participants)<br><br>(83 HIV+ participants lost to follow-up,<br>127 HIV+ non-adherent,<br>252 HIV+ completers) | To elicit information about participants' thoughts and opinions about the trial and the reasons for non-adherence.<br>To identify associations between loss to follow-up/ non-adherence and selected risk factors. | Individual interviews,<br>Focus Group Discussion,<br><br>Survey instrument with primarily close-ended questions (developed through Interviews & Focus Group Discussion) |
| Huerga (2016) | Kenya, Swaziland | urban, rural | Two Prospective cohort studies,<br><br>Operational assessment | Patients<br><br><br>Facilities | (n= 550 HIV+ adults in Kenya; and 696 HIV+ adults in Swaziland)<br>(1 urban private TB/HIV clinic in Kenya, two rural primary care clinics in Swaziland) | To assess the acceptability of TST, acceptability to return for TST reading, and initiation of 36 months IPT among PLHIV.<br>To assess the operational feasibility of TST (i.e. time spent on tuberculin skin testing and reading, staff involved, working space adjustments). | Cohort study data set,<br><br><br>Interviews with key personnel, direct observation, review of clinic registers |
| Jacobson (2017) | South Africa | rural | Qualitative study | Patients | (n= 30 HIV+ adults, including 17 IPT completers and 13 IPT defaulters) | To explore knowledge and attitudes of HIV+ adults towards TB, IPT, and HIV, and to explore reasons for initiating IPT and completing/ defaulting treatment. | Individual semi-structured interviews |
| Jarrett (2019) | South Africa | peri-urban | Multi-method assessment | Providers<br><br><br><br><br><br>Patients | (10 nurses working in HIV care, able to prescribe ART, who had participated in a nurse-centred intervention)<br><br><br>(10 HIV+ adults, who received HIV care at the intervention clinic, and had participated in a patient-centred intervention) | To examine nurse perceptions of clinical mentorship for promoting IPT uptake.<br>To examine continued barriers to change.<br>To assess adoption of the nurse-centred intervention.<br><br>To assess patient perceptions of in-queue health education for promoting IPT uptake.<br>To examine continued barriers to change.<br>To assess the fidelity to the patient-centred intervention. | Face-to-face in-depths interviews using semi-structured interview guides, Records of nurse attendance on workshops (training) or in-room consultations (mentoring),<br><br><br><br>Face-to-face in-depth interviews using semi-structured interview guides,<br>Records on patient attendance on in-queue education sessions |
| Khan (2014) | South Africa | - | Diagnostic performance evaluation | Patients | (737 HIV+ adults in two HIV clinics) | To determine the diagnostic performance of the symptom-based TB screening questionnaire recommended by WHO for people living with HIV in resource-limited settings among adults off and on ART.<br>To evaluate diagnostic performance of a screening strategy using chest radiology and screening questionnaire. | TB screening questionnaire,<br>Sputum specimens for microbiologic testing (smear microscopy, mycobacterial culture),<br>Patient information abstracted from medical charts,<br>Chest radiology |
| Lai (2019) | Ethiopia | - | Cross-sectional study | Providers | (106 TB-HIV service providers who worked at least 2 months in TB-HIV care and treatment service provision at one of 40 health facilities) | To examine clinician barriers to implementing IPT for people living with HIV, and to assess factors associated with high IPT coverage at the facility level. | Interviewer-administered questionnaire |
| Lester (2010) | South Africa | - | Qualitative methods study | Patients<br><br>Providers<br><br><br>Providers | (20 HIV+ adults attending for ART appointments)<br>(22 clinic health care providers incl. doctors, nurses, counsellors)<br>(9 doctors and 2 nurses) | To describe operational barriers to IPT delivery and uptake from a provider and user perspective, and to evaluate attitudes of clinic staff and patients to IPT. | 42 In-depth interviews,<br><br>42 In-depth interviews,<br><br><br>1 Focus group discussion with clinic staff |

Additional file 6. Detailed description of studies included in this review.

**Table 2. (…continued).**

| First Author (Year) | Country | Context | Study type | Study subject(s) of interest | | Study aims | Data collection approach |
|---|---|---|---|---|---|---|---|
| Little (2018) | Malawi | rural | Sub-study of the CHEPETSA trial | Patients | (974 adult participants newly diagnosed with HIV, initiated on IPT, including 732 IPT completers, 242 non-completers) | To evaluate predictors of IPT completion in individuals newly diagnosed with HIV (i.e. the relationship between IPT completion and individual and clinic-level predictors, including concomitant receipt of ART, age, pregnancy status, IPT side effects, and alcohol use). | Review of each participant's study file (adherence based on the number of INH pills dispensed at each visit)<br>Study team-administered questionnaires including questions about TB symptoms, possible IPT-related adverse events, ART status, pregnancy, and the number of INH pills dispensed). |
| McRobie (2017) | Uganda | - | Facility-level policy implementation assessment | Facilities | (24 facilities) | To assess the implementation of national policies about HIV testing, treatment and retention in 24 health facilities (policy and practice comparison). | Review of national HIV policies, 24 structured health facility surveys |
| | | | | Stake-holders | (7 Key stakeholders, incl. policymakers, implementers, researchers) | To explore site-specific implementation gaps and differences between sites. | 7 Key informant interviews |
| Meribe (2020) | Nigeria | - | Assessment of a provider-focused intervention to increase IPT initiation and completion | Facilities | (27 primary and secondary level of care military clinics and hospitals) | To describe the IPT scale-up intervention and results of the activities implemented at military healthcare facilities. | Review of routinely collected program data retrieved from the district health information system (DHIS-2), which were entered at the facility level from paper registers into DHIS-2, Quality assessments conducted at the sites during quarterly site visits |
| Mindachew (2011) | Ethiopia | urban | Analytical cross-sectional study | Patients | (319 HIV+ adults taking IPT for at least one month) at 4 hospitals | To determine self-reported adherence to IPT within seven days recall period among HIV+ adults and to assess factors associated with adherence to IPT. | Interviewer administered pre-tested structured questionnaire, including self-reported adherence to IPT |
| *Mugomeri (2018) | Lesotho | - | Qualitative study | Providers Stake-holders | (42 public healthcare workers, 2 Ministry of Health key informants, 6 representatives of partner organizations) | To establish barriers to the implementation of IPT within the health system context of Lesotho. | Individual semi-structured interviews |
| Mugomeri 2019 | Lesotho | - | Retrospective cohort study | Patients | 4122 HIV+ patients enrolled in HIV care, on ART at eight district hospitals | To assess the IPT initiation rate and retention of PLHIV on IPT, and to assess predictors of IPT initiation and interruption. | Review of the paper-based ART and IPT registers of HIV+ hospital attendees enrolled in HIV care between 2004 and 2016 |
| Munseri (2008) | Tanzania | - | Sub-study of the TB vaccine trial | Patients | (568 HIV+ adult vaccine trial participants who were offered IPT) | To assess factors related to completion of IPT among HIV+ adults. | Review of study records (i.e. the number of patients offered and accepted IPT, completion, non-completion), |
| | | | | Patients | (n= 109 HIV+ completers, 8 non-completers) | To assess knowledge, attitudes and beliefs regarding IPT. | Interviews using a standard questionnaire |
| Ngamvithaya pong (1997) | Thailand | - | Prospective cohort study | Patients | (n= 412 HIV+ adults initiating IPT, including blood donors, outpatients, female commercial sex workers, anonymous clinic clients) | To determine the completion rate and level of adherence to nine months IPT among HIV+ individuals in northern Thailand. | Adherence Assessment (based on monthly clinic attendance for IPT collection and pill count), |
| | | | | Patients | (72 HIV+ adults, incl. 50 defaulter and 22 who missed clinic appointments) | To determine the reasons for non-adherence. | Interviews, |
| | | | | Patients | (n= 28 completers) | To determine motivation and methods used in order to adhere to IPT. | 5 Focus Group Discussions |

Additional file 6. Detailed description of studies included in this review.

**Table 2. (…continued).**

| First Author (Year) | Country | Context | Study type | Study subject(s) of interest | | Study aims | Data collection approach |
|---|---|---|---|---|---|---|---|
| *Okot-Chono (2009) | Uganda | peri-urban, rural | Record review, qualitative study | Districts | (5 districts) | To assess barriers to implementation of TB-HIV collaborative services. | Review of TB registers to assess the TB-HIV services utilization (incl. CPT) among 333 TB patients, |
| | | | | Patients<br>Providers<br>CM | (125 TB patients)<br>(65 health providers)<br>(70 community members) | To assess respondents knowledge, attitudes, practices and beliefs about TB-HIV collaborative services. | Standardised tools applied for 26 Focus Group Discussions, and 34 Key informant interviews, |
| | | | | Providers | (? facility in-charges, ? focal persons for TB/HIV, ? district health officers, ? expert patients) | To identify reasons explaining the level of implementation of TB/HIV collaborative services. | 28 In-depth interviews |
| Reddy (2020) | India | 60% rural | Mixed-methods study | Patients | (4020 newly registered PLHIV started on ART in 2 districts) | To assess the implementation of IPT among PLHIV, who were newly initiated on ART between 01/2017 and 06/2018, including [%] eligible for, [%] who started IPT, and among those initiated, [%] who completed IPT.<br>To assess factors associated with initiation and non-completion of IPT. | Review of routinely collected program data, |
| | | | | Providers | (22 Healthcare providers involved in IPT implementation) | Reasons for non-initiation and non-completion from healthcare providers' perspective. | Face-to-face in-depth interviews, |
| | | | | Patients | (8 HIV+ adult patients, including 5 IPT completers, and 3 interrupters) | Reasons for non-initiation and non-completion from a patients' perspective. | Face-to-face in-depth interviews |
| Rowe (2005) | South Africa | rural | Record review, qualitative study | Patients | (n= 87 HIV+ adult clinic attendees who initiated 6-month IPT) | To describe the study population of HIV+ individuals attending the hospital-based HIV clinic that initiated IPT. | Review of patient records<br>(i.e. patients eligible, initiated, ongoing collection of medication, side effects), |
| | | | | Patients<br>Providers | (18 HIV+ adult clinic attendees, incl. 6 eligible for IPT, 6 IPT interrupters and 6 IPT completers)<br>(2 health care workers who routinely administered IPT) | To explore HIV+ patients and health workers' perspectives regarding the adherence to IPT and to derive lessons for improving access to care amongst HIV+ individuals in resource-poor settings. | In-depth interviews applying open-ended questions |
| Selehelo (2019) | South Africa | - | Qualitative study | Patients | (14 HIV+ community health centre attendees on IPT for at least 6 weeks) | To explore and describe the experiences of PLHIV regarding IPT provision to derive strategies to improve IPT uptake in a community health centre (CHC). | Face-to-face in-depth interviews |
| Szakacs (2006) | South Africa | sub-urban | Assessment of adherence to IPT, Cross-sectional study | Patients | (301 HIV+ adult outpatients receiving IPT at two hospitals) | To assess adherence to IPT (using in-house prepared INH urine test strips) among HIV+ adults and to identify predictors of positive urine test results. | Once off adherence assessment (urine-based INH metabolite testing) and interviewer-administered questionnaire (developed from the review of previous studies) |

Additional file 6. Detailed description of studies included in this review.

**Table 2. (…continued).**

| First Author (Year) | Country | Context | Study type | Study subject(s) of interest | | Study aims | Data collection approach |
|---|---|---|---|---|---|---|---|
| Tram (2019) | Uganda | rural | Cross-sectional study (sub-study of SEARCH HIV test and treat trial) | Patients | (305 adult HIV patients on ART who started IPT in 5 government-sponsored clinics) | To evaluate IPT completion rates among trial participants receiving IPT as part of a patient-centred, multicomponent HIV care model, and patients receiving IPT as routine non-differentiated HIV care in the same health centre. | Review of IPT treatment registers to determine IPT completion (based on receipt of 6 months of IPT), |
| | | | | Patients | (180 of the 305 adult HIV patients, including 94 non-differentiated care attendees, and 86 differentiated care attendees) | To assess individual, interpersonal, and structural predictors of IPT completion, as well as underlying potential differences in IPT completion by care model. | Nested (interviewer-administered) survey on barriers to IPT completion |
| Van Ginderdeuren (2019) | South Africa | urban | Before-and-after study) | Facilities | (n= 3 primary care health facilities) | To compare IPT and TST uptake at three primary care health facilities before and after the intervention. | Record review (IPT uptake and TST placement based on routine clinic registers in paper/ electronic format; INH prescription based on pharmacy records), |
| | | | | Facilities | (n= 2 primary care health facilities) | To determine fidelity to the 2014 South African IPT guidelines at two primary care health facilities, following the implementation of the intervention. | Review of paper medical files and electronic laboratory databases to compare HCWs decisions made with decisions expected, |
| | | | | Providers | (25 health care workers involved in IPT prescription | To identify remaining barriers for IPT and TST implementation perceived by HCWs. | Structured questionnaire survey |
| Wambiya (2018) | Kenya | urban (cities) | Qualitative study | Providers | (n= 18 healthcare providers working in three HIV clinics) | To explore factors influencing the acceptability of IPT among healthcare providers in selected HIV clinics in Nairobi. | Individual semi-structured in-depth interviews |
