## Supplementary material for "Mixed methods systematic review and metasummary about barriers and facilitators for the implementation of cotrimoxazole and isoniazid – preventive therapies for people living with HIV": S7 Additional file. Assessment of methodological strengths and limitations.

Additional file 7. Assessment of methodological strengths and limitations.

| Category of study designs | | QUAL | | | | | | | QUAN | | | | | | | | | | | | | | | | | | | | | | MULTI | | | | | | | | | MIX | |
|---|---|---|---|---|---|---|---|---|---|---|---|---|---|---|---|---|---|---|---|---|---|---|---|---|---|---|---|---|---|---|---|---|---|---|---|---|---|---|---|---|---|
| **First author [citation]** | **Methodological quality criteria** | Jacobson [1] | Lester [2] | Mugomeri [3] | Okwera [4] | Selehelo [5] | Sibanda [6] | Wambiya [7] | Chang [8] | Naikoba [9] | Adepoju [10] | Ansa [11] | Khan [12] | Little [13] | Lougwagie [14] | Mindachew [15] | Mugomeri [16] | Munseri [17] | Ngamvithayapong [18] | Szakacs [19] | Tram [20] | Van Ginderdeuren [21] | Aisu [22] | Chan [23] | Faust [24] | Horwood [25] | Kamuhabwa [26] | Lai [27] | Meribe [28] | Mwambete [29] | Durovni [30] | Gust [31] | Rowe [32] | Huerga [33] | Kamuhabwa [34] | Jarrett [35] | Luyirika [36] | McRobie [37] | Okot-Chono [38] | Catalani [39] | Reddy [40] |
| Screening questions (for all types of study designs) | S1 | Y | Y | Y | Y | Y | Y | Y | U | Y | Y | Y | Y | Y | Y | Y | U | Y | Y | Y | Y | Y | Y | Y | Y | Y | Y | Y | Y | U | Y | Y | Y | Y | Y | Y | Y | U | U | U | Y |
| | S2 | Y | Y | Y | Y | Y | Y | Y | Y | Y | Y | Y | Y | Y | Y | Y | Y | Y | Y | Y | Y | Y | Y | U | Y | Y | Y | Y | Y | Y | Y | Y | Y | Y | Y | Y | U | Y | Y | Y | Y |
| 1. Qualitative (ethnography, phenomenology, narrative research, grounded theory, case study, qualitative description) | 1.1. | Y | U | Y | U | Y | U | Y | | | | | | | | | | | | | | | | | | | | | | | U | Y | U | | U | U | U | U | U | Y | U |
| | 1.2. | Y | Y | U | Y | Y | Y | Y | | | | | | | | | | | | | | | | | | | | | | | U | Y | Y | | Y | Y | U | Y | Y | Y | Y |
| | 1.3. | Y | Y | Y | U | Y | Y | Y | | | | | | | | | | | | | | | | | | | | | | | U | Y | Y | | Y | Y | N | Y | Y | Y | Y |
| | 1.4. | Y | Y | Y | Y | Y | Y | Y | | | | | | | | | | | | | | | | | | | | | | | N | Y | Y | | Y | Y | N | Y | N | Y | Y |
| | 1.5. | Y | Y | Y | Y | Y | Y | Y | | | | | | | | | | | | | | | | | | | | | | | U | Y | Y | | Y | Y | N | Y | U | Y | Y |
| 2. Quantitative (randomised controlled clinical trial, cluster or individual, randomised cross-over study) | 2.1. | | | | | | | | Y | Y | | | | | | | | | | | | | | | | | | | | | U | | | | | | | | | | |
| | 2.2. | | | | | | | | Y | Y | | | | | | | | | | | | | | | | | | | | | U | | | | | | | | | | |
| | 2.3. | | | | | | | | Y | Y | | | | | | | | | | | | | | | | | | | | | Y | | | | | | | | | | |
| | 2.4. | | | | | | | | N | Y | | | | | | | | | | | | | | | | | | | | | U | | | | | | | | | | |
| | 2.5. | | | | | | | | Y | Y | | | | | | | | | | | | | | | | | | | | | Y | | | | | | | | | | |
| 3. Quantitative (non-randomised studies, e.g. non-randomised controlled trials, cohort study, case-control study, cross-sectional analytical study) | 3.1. | | | | | | | | | | Y | Y | Y | N | Y | Y | Y | N | Y | N | Y | Y | | | | | | | | | | Y | Y | Y | | | | | | | Y |
| | 3.2. | | | | | | | | | | Y | Y | Y | Y | Y | Y | Y | Y | Y | Y | Y | Y | | | | | | | | | | Y | Y | Y | | | | | | | Y |
| | 3.3. | | | | | | | | | | N | Y | Y | Y | Y | Y | Y | Y | Y | Y | Y | Y | | | | | | | | | | Y | Y | Y | | | | | | | Y |
| | 3.4. | | | | | | | | | | Y | N | Y | Y | Y | U | Y | Y | Y | Y | Y | Y | | | | | | | | | | Y | N | Y | | | | | | | Y |
| | 3.5. | | | | | | | | | | N | U | Y | Y | Y | U | U | U | Y | Y | Y | U | | | | | | | | | | Y | Y | Y | | | | | | | Y |
| 4. Quantitative (descriptive, e.g. incidence or prevalence study without a comparison group, survey, case series, case report) | 4.1. | | | | | | | | | | | | | | | | | | | | | | Y | N | Y | Y | Y | Y | Y | Y | | | | U | Y | Y | Y | Y | Y | Y | |
| | 4.2. | | | | | | | | | | | | | | | | | | | | | | Y | Y | Y | Y | Y | Y | N | U | | | | Y | U | Y | U | U | U | Y | |
| | 4.3. | | | | | | | | | | | | | | | | | | | | | | Y | Y | U | Y | Y | Y | Y | N | | | | Y | Y | Y | U | N | U | U | |
| | 4.4. | | | | | | | | | | | | | | | | | | | | | | / | Y | N | U | U | Y | U | U | | | | U | Y | U | U | / | N | N | |
| | 4.5. | | | | | | | | | | | | | | | | | | | | | | / | / | / | Y | / | Y | / | Y | | | | / | Y | / | / | / | / | / | |
| 5. Mixed methods (convergent design, sequential explanatory design, sequential exploratory design) | 5.1. | | | | | | | | | | | | | | | | | | | | | | | | | | | | | | | | | | | | | | | U | Y |
| | 5.2. | | | | | | | | | | | | | | | | | | | | | | | | | | | | | | | | | | | | | | | N | N |
| | 5.3. | | | | | | | | | | | | | | | | | | | | | | | | | | | | | | | | | | | | | | | Y | Y |
| | 5.4. | | | | | | | | | | | | | | | | | | | | | | | | | | | | | | | | | | | | | | | Y | N |
| | 5.5. | | | | | | | | | | | | | | | | | | | | | | | | | | | | | | | | | | | | | | | N | Y |

A description of each methodological quality criteria is presented in the "Mixed Methods Appraisal tool" (MMAT, 2018) Hong, Q.N., et al. Mixed methods appraisal tool (MMAT), version 2018. IC Canadian Intellectual Property Office, Industry Canada 2018; Available from http://mixedmethodsappraisaltoolpublic.pbworks.com/w/file/fetch/127916259/MMAT_2018_criteria-manual_2018-08-01_ENG.pdf.

**Responses: Y - Yes; N - No; U – Unclear (Can't tell); / - Not applicable for this study.**
**Categories of study design: QUAN – quantitative design, QUAL – qualitative design, MIX – mixed methods design; MULTI – multimethod design.**

Additional file 7. Assessment of methodological strengths and limitations.

**References** (studies assessed using the MMAT tool)

21. Van Ginderdeuren, E., et al., *Health system barriers to implementation of TB preventive strategies in South African primary care facilities.* PLoS One, 2019. **14**(2): p. e0212035.
22. Aisu, T., et al., *Preventive chemotherapy for HIV-associated tuberculosis in Uganda: an operational assessment at a voluntary counselling and testing centre.* AIDS 1995. **9**(3): p. 267-273.
23. Chan, A.K., et al., *The Lablite project: A cross-sectional mapping survey of decentralized HIV service provision in Malawi, Uganda and Zimbabwe.* BMC Health Services Research, 2014. **14**.
24. Faust, L., et al., *How are high burden countries implementing policies and tools for latent tuberculosis infection? A survey of current practices and barriers.* Health Sci Rep, 2020. **3**(2): p. e158.
25. Horwood, C., et al., *Prevention of mother to child transmission of HIV (PMTCT) programme in KwaZulu-Natal, South Africa: an evaluation of PMTCT implementation and integration into routine maternal, child and women's health services.* Trop Med Int Health, 2010. **15**(9): p. 992-9.
26. Kamuhabwa, A.A.R. and V. Manyanga, *Challenges facing effective implementation of cotrimoxazole prophylaxis in children born to HIV-infected mothers in the public health facilities.* Drug Healthcare and Patient Safety, 2015. **7**: p. 147-156.
27. Lai, J., et al., *Provider barriers to the uptake of isoniazid preventive therapy among people living with HIV in Ethiopia.* Int J Tuberc Lung Dis, 2019. **23**(3): p. 371-377.
28. Meribe, S.C., et al., *Sustaining tuberculosis preventive therapy scale-up through direct supportive supervision.* Public Health Action, 2020. **10**(2): p. 60-63.
29. Mwambete, K.D. and A.A. Kamuhabwa, *Resistance of commensal intestinal Escherichia coli and other enterics to co-trimoxazole and commonly used antibiotics in HIV/AIDS patients.* Clinical Microbiology: Open Access, 2013.
30. Durovni, B., et al., *The implementation of isoniazid preventive therapy in HIV clinics: the experience from the TB/HIV in Rio (THRio) study.* AIDS 2010. **24**(Suppl 5): p. S49.
31. Gust, D.A., et al., *Risk factors for non-adherence and loss to follow-up in a three-year clinical trial in Botswana.* PLoS One, 2011. **6**(4): p. e18435.
32. Rowe, K.A., et al., *Adherence to TB preventive therapy for HIV-positive patients in rural South Africa: implications for antiretroviral delivery in resource-poor settings?* Int J Tuberc Lung Dis, 2005. **9**(3): p. 263-9.
33. Huerga, H., et al., *Implementation and Operational Research: Feasibility of Using Tuberculin Skin Test Screening for Initiation of 36-Month Isoniazid Preventive Therapy in HIV-Infected Patients in Resource-Constrained Settings.* J Acquir Immune Defic Syndr, 2016. **71**(4): p. e89-95.
34. Kamuhabwa, A.A., R. Gordian, and R.F. Mutagonda, *Implementation of co-trimoxazole preventive therapy policy for malaria in HIV-infected pregnant women in the public health facilities in Tanzania.* Drug Healthc Patient Saf, 2016. **8**: p. 91-100.
35. Jarrett, B.A., et al., *Promoting Tuberculosis Preventive Therapy for People Living with HIV in South Africa: Interventions Hindered by Complicated Clinical Guidelines and Imbalanced Patient-Provider Dynamics.* Aids and Behavior, 2019. **24**(4): p. 1106-1117.
36. Luyirika, E., et al., *Scaling Up Paediatric HIV Care with an Integrated, Family-Centred Approach: An Observational Case Study from Uganda.* Plos One, 2013. **8**(8).
37. McRobie, E., et al., *HIV policy implementation in two health and demographic surveillance sites in Uganda: Findings from a national policy review, health facility surveys and key informant interviews.* Implementation Science, 2017. **12**(1).
38. Okot-Chono, R., et al., *Health system barriers affecting the implementation of collaborative TB-HIV services in Uganda.* The international journal of tuberculosis and lung disease, 2009. **13**(8): p. 955-961.
39. Catalani, C., et al., *A Clinical Decision Support System for Integrating Tuberculosis and HIV Care in Kenya: A Human-Centered Design Approach.* Plos One, 2014. **9**(8).
40. Reddy, M.M., et al., *To start or to complete? - Challenges in implementing tuberculosis preventive therapy among people living with HIV: a mixed-methods study from Karnataka, India.* Glob Health Action, 2020. **13**(1): p. 1704540.
