## Supplementary material for "Mixed methods systematic review and metasummary about barriers and facilitators for the implementation of cotrimoxazole and isoniazid – preventive therapies for people living with HIV": S8 Additional file. Metasummary tables.

**Table 1. Summary of all barrier themes that emerged for both preventive therapies, identified in forty studies included in this review.**

| Health system component<br>Emerging barrier themes | No. of articles | Supporting articles |
|---|---|---|
| Patient & community related barriers | | |
| Adverse reactions (side effects) | 18 | [1-18] |
| Lacking financial and organisational feasibility | 14 | [1, 2, 4, 6, 8, 10, 12, 13, 15, 19-23] |
| Knowledge gaps & misperceptions | 14 | [2, 4, 10-14, 17-22, 24] |
| Patients‘ lacking motivation | 12 | [2, 4, 6, 11-15, 18, 19, 21, 22] |
| Forgetfulness | 4 | [2, 6, 9, 23] |
| Patients' HIV denial, religion & competing medicinal approaches | 7 | [2, 3, 10, 13, 15, 16, 19] |
| Stigma & fear of rejection/ discrimination | 11 | [2, 3, 8, 10, 13, 15, 16, 19, 20, 22, 23] |
| Influence of relatives and friends | 8 | [2, 3, 8, 11-13, 15, 19] |
| *Socio-demographic, lifestyle and clinical factors | 10 | [1, 2, 7, 8, 10, 12, 16, 18, 25, 26] |
| *Concerns about the efficacy of CPT | 2 | [9, 11] |
| Health provider related barriers | | |
| Shortage of health providers | 12 | [1, 3, 7, 12, 14, 17, 18, 22, 24, 27-29] |
| Knowledge- and training gaps | 12 | [1, 3, 7, 12, 18, 19, 21, 22, 24, 28, 30, 31] |
| *Attitudes, beliefs & fear to induce INH resistance | 10 | [5, 7, 12, 14, 17-19, 21, 24, 28] |
| *Provider-patient communication | 3 | [6, 20, 24] |
| Clinical information related barriers | | |
| Inaccurate recording and lack of integration of health records | 9 | [3, 7, 13, 17, 22, 24, 28, 30, 32] |
| Ineffective monitoring, evaluation & surveillance | 4 | [7, 17, 22, 29] |
| Pharmaceutical management related barriers | | |
| Stock-outs/ Shortages of PT’s | 16 | [3, 4, 12, 14, 15, 17, 18, 20, 22-24, 26, 28, 29, 33, 34] |
| Lack of pharmaceutical personnel | 2 | [3, 29] |
| *Lack of written instructions | 1 | [3] |
| Service delivery related barriers | | |
| Sub-optimal service delivery | 14 | [1, 4, 12, 14, 15, 18-20, 22, 24, 27, 29, 30, 32] |
| Inadequate facility infrastructure | 3 | [4, 22, 29] |
| Poor integration of preventive therapy-related services | 10 | [1, 3, 7, 18, 20-22, 29, 30, 33] |
| Patient loss to follow-up | 7 | [2, 12, 13, 17, 25, 29, 30] |
| *Ruling out TB disease | 15 | [1, 5, 7, 12, 14, 17, 18, 20-22, 24, 28, 35-37] |
| *Investigation of TB suspects | 8 | [7, 14, 17, 21, 22, 28, 36, 37] |
| Health system financing related barriers | | |
| Health system funding issues | 4 | [4, 7, 29, 35] |
| Vertical funding | 3 | [7, 22, 29] |
| Leadership & governance related barriers | | |
| Issues regarding policies & guidelines | 8 | [4, 7, 17, 18, 21, 22, 24, 35] |
| Lacking leadership and coordination | 6 | [1, 7, 18, 20, 22, 30] |
| Lacking top-down policy support and management issues | 4 | [18, 20, 22, 24] |
| Inadequate planning | 2 | [7, 22] |
| Lacking engagement of stakeholders | 3 | [7, 18, 22] |

Total number of themes identified (N= 32), Intervention specific themes (n= 7). The asterisk (*) is used to highlight intervention-specific themes.
CPT - cotrimoxazole preventive therapy; IPT – isoniazid preventive therapy, PT - preventive therapy; INH – isoniazid..

S8 Additional file. Metasummary tables.

**Table 2. Summary of barrier themes that emerged for cotrimoxazole preventive therapy.**

| Health system component<br>Emerging barrier themes | No. of articles | Relevant articles |
|---|---|---|
| <u>Patient & community related barriers</u> | 7 | [3, 4, 7, 9, 11, 15, 22] |
| Adverse reactions (side effects) | 6 | [3, 4, 7, 9, 11, 15] |
| Lacking financial and organisational feasibility | 3 | [4, 15, 22] |
| Knowledge gaps & misperceptions | 2 | [4, 11] |
| Patients‘ lacking motivation | 4 | [4, 11, 15, 22] |
| Forgetfulness | 1 | [9] |
| Patients' HIV denial, religion & competing medicinal approaches | 2 | [3, 15] |
| Stigma & fear of rejection/ discrimination | 3 | [3, 15, 22] |
| Influence of relatives and friends | 3 | [3, 11, 15] |
| *Concerns about the efficacy of CPT | 2 | [9, 11] |
| <u>Health provider related barriers</u> | 4 | [3, 22, 30, 31] |
| Shortage of health providers | 1 | [3] |
| Knowledge and training gaps | 4 | [3, 22, 30, 31] |
| <u>Clinical information related barriers</u> | 5 | [3, 7, 22, 30, 32] |
| Inaccurate recording and lack of integration of health records | 5 | [3, 7, 22, 30, 32] |
| Ineffective monitoring, evaluation & surveillance | 2 | [7, 22] |
| <u>Pharmaceutical management related barriers</u> | 6 | [3, 4, 7, 15, 22, 33] |
| Stock-outs/ Shortages of PT’s | 6 | [3, 4, 7, 15, 22, 33] |
| Lack of pharmaceutical personnel | 1 | [3] |
| *Lack of written instructions | 1 | [3] |
| <u>Service delivery related barriers</u> | 8 | [3, 4, 7, 15, 22, 30, 32, 33] |
| Sub-optimal service delivery | 5 | [4, 15, 22, 30, 32] |
| Inadequate facility infrastructure | 2 | [4, 22] |
| Poor integration of preventive therapy-related services | 5 | [3, 7, 22, 30, 33] |
| Patient loss to follow-up | 1 | [30] |
| <u>Health system financing related barriers</u> | 3 | [4, 7, 22] |
| Health system funding issues | 2 | [7, 22] |
| Vertical funding | 2 | [4, 7] |
| <u>Leadership & governance related barriers</u> | 3 | [4, 22, 30] |
| Issues regarding policies & guidelines | 2 | [4, 22] |
| Lacking leadership & coordination | 2 | [22, 30] |
| Lacking top-down policy support & management issues | 1 | [22] |
| Inadequate planning | 1 | [22] |
| Lacking engagement of stakeholders | 1 | [22] |

Barrier themes presented in this table emerged from eleven peer-reviewed articles identified with barriers to CPT.
Total number of themes identified (N= 27), Intervention specific themes (n= 2). The asterisk (*) is used to highlight intervention-specific themes.
CPT - cotrimoxazole preventive therapy; PT - preventive therapy.

S8 Additional file. Metasummary tables.

**Table 3. Summary of barrier themes that emerged for isoniazid preventive therapy.**

| Health system component<br>Emerging barrier themes | No. of articles | Relevant articles |
|---|---|---|
| Patient & community related barriers | 21 | [1, 2, 5-8, 10, 12-14, 16-26] |
| Adverse reactions (side effects) | 13 | [1, 2, 5-8, 10, 12-14, 16-18] |
| Lacking financial and organisational feasibility | 12 | [1, 2, 6, 8, 10, 12, 13, 19-23] |
| Knowledge gaps & misperceptions | 12 | [2, 10, 12-14, 17-22, 24] |
| Patients‘ lacking motivation | 9 | [2, 6, 11-14, 18, 19, 21] |
| Forgetfulness | 3 | [2, 6, 23] |
| Patients' HIV denial, religion & competing medicinal approaches | 5 | [2, 10, 13, 16, 19] |
| Stigma & fear of rejection/ discrimination | 9 | [2, 8, 10, 13, 16, 19, 20, 22, 23] |
| Influence of relatives and friends | 5 | [2, 8, 12, 13, 19] |
| *Socio-demographic, lifestyle and clinical factors | 10 | [1, 2, 7, 8, 10, 12, 16, 18, 25, 26] |
| Health provider related barriers | 16 | [1, 5-7, 12, 14, 17-22, 24, 27-29] |
| Shortage of health providers | 11 | [1, 7, 12, 14, 17, 18, 22, 24, 27-29] |
| Knowledge and training gaps | 9 | [1, 7, 12, 18, 19, 21, 22, 24, 28] |
| *Attitudes, beliefs & fear to induce INH resistance | 10 | [5, 7, 12, 14, 17-19, 21, 24, 28] |
| *Provider-patient communication | 3 | [6, 20, 24] |
| Clinical information related barriers | 7 | [7, 13, 17, 22, 24, 28, 29] |
| Inaccurate recording and lack of integration of health records | 6 | [7, 13, 17, 22, 24, 28] |
| Ineffective monitoring, evaluation & surveillance | 4 | [7, 17, 22, 29] |
| Pharmaceutical management related barriers | 10 | [12, 14, 17, 18, 20, 23, 24, 28, 29, 34] |
| Stock-outs/ Shortages of PT’s | 10 | [12, 14, 17, 18, 20, 23, 24, 28, 29, 34] |
| Lack of pharmaceutical personnel | 1 | [29] |
| Service delivery related barriers | 21 | [1, 2, 5, 7, 12-14, 17-22, 24, 25, 27-29, 35-37] |
| Sub-optimal service delivery | 10 | [1, 12, 14, 18-20, 22, 24, 27, 29] |
| Inadequate facility infrastructure | 1 | [29] |
| Poor integration of preventive therapy-related services | 7 | [1, 7, 18, 20-22, 29] |
| Patient loss to follow-up | 6 | [2, 12, 13, 17, 25, 29] |
| *Ruling out TB disease | 15 | [1, 5, 7, 12, 14, 17, 18, 20-22, 24, 28, 35-37] |
| *Investigation of TB suspects | 8 | [7, 14, 17, 21, 22, 28, 36, 37] |
| Health system financing related barriers | 4 | [7, 22, 29, 35] |
| Health system funding issues | 3 | [7, 29, 35] |
| Vertical funding | 3 | [7, 22, 29] |
| Leadership & governance related barriers | 9 | [1, 7, 17, 18, 20-22, 24, 35] |
| Issues regarding policies & guidelines | 7 | [7, 17, 18, 21, 22, 24, 35] |
| Lacking leadership & coordination | 5 | [1, 7, 18, 20, 22] |
| Lacking top-down policy support & management issues | 4 | [18, 20, 22, 24] |
| Inadequate planning | 2 | [7, 22] |
| Lacking engagement of stakeholders | 3 | [7, 18, 22] |

Barrier themes presented in this table emerged from twenty-eight peer-reviewed articles identified with barriers to IPT.
Total number of themes identified (N= 30), Intervention specific themes (n= 5). The asterisk (*) is used to highlight intervention-specific themes.
IPT - isoniazid preventive therapy; INH - isoniazid.

S8 Additional file. Metasummary tables.

**Table 4. Relative magnitude of health system components identified as the source of barriers to cotrimoxazole preventive therapy.**

| Health system component from which barriers arose \ Peer-reviewed articles reporting barriers to CPT (N= 11) | Chan (2014) | Horwood (2010) | Kamuhabwa (2015) | Kamuhabwa(2016) | Lougwagie (2012) | Mugomeri (2018)* | Mwambete (2013) | Naikoba (2017) | Okot-Chono (2009)* | Okwera (2015) | Sibanda (2015) | Frequency | Effect size [%] |
|---|---|---|---|---|---|---|---|---|---|---|---|---|---|
| **Patient & community** | | | X | X | | X | X | | X | X | X | 7/11 | 64 |
| Health providers | | X | | X | | | | X | X | | | 4/11 | 36 |
| Clinical information | | X | | X | X | X | | | X | | | 5/11 | 45 |
| **Pharmaceutical management** | X | | X | X | | X | | | X | | X | 6/11 | **55** |
| **Service delivery** | X | X | X | X | X | X | | | X | | X | 8/11 | **73** |
| Health system financing | | | X | | | X | | | X | | | 3/11 | 27 |
| Leadership & governance | | X | X | | | | | | X | | | 3/11 | 27 |

*Two studies reported barriers to both preventive therapies (Okot-Chono (2009), Mugomeri (2018)). CPT - cotrimoxazole preventive therapy. Out of the 14 peer-reviewed articles identified for CPT, three articles (Luyirika, (2013), Chang (2015), Ansa (2014)) only addressed facilitators for CPT, and where therefore excluded from metasummary.

**Table 5. Relative magnitude of health system components identified as the source of barriers to isoniazid preventive therapy.**

| Health system component from which barriers arose \ Peer-reviewed articles reporting barriers to IPT (N= 28) | Adepoju (2020) | Aisu (1995) | Catalani (2014) | Durovni (2010) | Faust (2020) | Gust (2011) | Huerga (2016) | Jacobson (2017) | Jarrett (2019) | Khan (2014) | Lai (2019) | Lester (2010) | Little (2018) | McRobie (2017) | Meribe (2020) | Mindachew (2011) | Mugomeri (2018)* | Mugomeri (2019) | Munseri (2008) | Ngamvithayapong (1997) | Okot-Chono (2009)* | Reddy (2020) | Rowe (2005) | Selehelo (2019) | Szakacs (2006) | Tram (2020) | Van Ginderdeuren (2019) | Wambiya (2018) | Frequency | Effect size [%] |
|---|---|---|---|---|---|---|---|---|---|---|---|---|---|---|---|---|---|---|---|---|---|---|---|---|---|---|---|---|---|---|
| **Patient & community** | | X | | X | | X | | X | X | | X | X | X | | | X | X | X | X | X | X | X | X | X | X | X | X | X | 21/28 | **75** |
| **Health providers** | X | X | X | X | | | | X | X | | X | X | | X | | X | X | | | | X | X | | X | | | X | X | 16/28 | **57** |
| Clinical information | | | X | | | | | | X | | | | | X | | | X | | | | X | | X | | | | X | | 7/28 | 33 |
| Pharmaceutical management | | | X | | | | | X | X | | | | | X | X | | | | | | | X | | X | X | | X | X | 10/28 | 36 |
| Service delivery | X | X | X | X | X | X | X | X | X | X | X | X | X | X | | | X | | | | X | X | X | X | | | X | X | 21/28 | **75** |
| Health system financing | | | | | X | | | | | | | | | X | | | X | | | | X | | | | | | | | 4/28 | 4 |
| Leadership & governance | | | | X | X | | | X | X | | | X | | | | | X | | | | X | | | | | | X | X | 9/28 | 32 |

All twenty-eight articles identified for IPT were included for metasummary. *Two studies reported barriers to both preventive therapies (Okot-Chono (2009), Mugomeri (2018)). IPT - isoniazid preventive therapy.

S8 Additional file. Metasummary tables.
